# Artificial Intelligence for Predicting Treatment Adherence in Opioid Use Disorder: A Scoping Review

**DOI:** 10.1101/2025.07.10.25331331

**Authors:** Aghna Wasim, Ali Abud, Samir Malick, Nazeefa Arifina Nashrah, Veronika Lošanová, Hetvi Raimugia, Marisha Karim, Minh Van Khanh Le, Venusha Baskarathasan, Charlotte Gibson, Siba Alkhatib, Silvia S. Martins

## Abstract

**Background and Aim:** Opioid use disorder (OUD) is a chronic condition in which an individual engages in the persistent use of opioids that causes significant distress and negatively impacts their societal functioning. Treatment for OUD involves pharmacological therapies such as methadone, buprenorphine, and naltrexone, typically used in combination with behavioral interventions such as counselling and cognitive behavioural therapy. However, non-adherence to OUD treatment is high, potentially leading to negative outcomes like relapse and increased risk of overdose. Therefore, identifying patients at risk of treatment nonadherence is essential to ensure that OUD is adequately managed. Models utilizing AI and ML techniques have emerged as promising candidates to achieve risk stratification in this patient population. We conducted a scoping review to capture and systematically map existing literature on AI and ML applications predicting adherence to treatment in individuals with OUD.

**Methods:** Ovid MEDLINE, Embase, PsycINFO, Web of Science, Scopus, CINAHL, IEEE Xplore, and ACM Digital Library were searched to identify and obtain peer-reviewed empirical research articles published from inception to October 7, 2024. Twenty-two studies were selected to be included in the review.

**Results:** All studies that matched our search criteria were published after 2018 and predominantly conducted in the United States. Random forest models were frequently identified as the top performer although significant variability in algorithms, evaluation metrics, and key predictors was noted in the literature.

**Conclusion:** The need for future research to cover more geographical locations, diversify patient populations, focus on a standardized group of models and outcomes, and utilize larger samples was highlighted.

## Introduction

Opioid use disorder (OUD) is a chronic condition characterized by a prolonged, problematic pattern of opioid usage leading to significant distress or impairment in everyday life [1]. OUD affects more than 16 million people globally and claims over 120,000 lives per year [1]. According to the 2023 U.S. National Survey on Drug Use and Health, 8.6 million people aged 12 years or older in the United States had misused prescription opioids in the preceding year [2]. It is difficult to accurately determine OUD prevalence as national surveys often exclude high-risk populations such as homeless, hospitalized, or incarcerated individuals [3]. Nevertheless, recent estimates have indicated stability or even a small decline in the prevalence of OUD in the United States [3]. Despite a decrease in prescription opioid prescribing in the US over the past decade, over 6 million people in the country continue to have OUD, heroin use has substantially increased, and synthetic opioid-related mortality constitutes a major cause of death [3, 4]. Furthermore, individuals with untreated OUD often experience challenges in their physical, emotional, and social functioning, leading to a decline in their quality of life [5, 6]. Therefore, it is important that OUD is managed effectively.

OUD management is typically pharmacological and involves the routine administration of methadone, buprenorphine, or naltrexone as the gold standard [7–10]. Pharmacological treatment for OUD reduces symptoms of opioid withdrawal, decreases cravings, attenuates risk of relapse, and lowers the likelihood of overdose mortality while improving psychosocial functioning [7, 10]. OUD may also be managed using behavioural interventions, such as counselling, motivational interviewing, cognitive behavioural therapy, acceptance and commitment therapy, relational psychotherapy, and contingency management [8, 11]. These interventions are often used in conjunction with medication and are aimed at addressing the psychosocial elements of OUD, increasing treatment compliance, minimizing drug relapse, and managing comorbid psychiatric disorders [8, 11].

The effectiveness of treatments for OUD is limited due to relatively high rates of nonadherence [12]. Nonadherence among individuals with OUD is associated with poor outcomes, including an increased risk of relapse and overdose as well as greater healthcare costs [12–14]. Reducing rates of nonadherence to treatment for OUD is thus of prime importance. This requires an accurate assessment of the likelihood of treatment adherence and a subsequent identification of individuals that are at risk, allowing for timely mitigation of any challenges or barriers that might be keeping them from continuing their treatment. Models that leverage machine learning (ML) and artificial intelligence (AI) can be used to assist in predicting adherence to OUD treatment. ML algorithms are a branch of AI that uses big data as input to identify patterns and compute predictions for outcomes of interest [15–18]. The benefit of ML approaches lies in the ability of models to learn or train on input data to continually improve their predictive performance [16–18].

Although several studies have explored the use of ML and AI in predicting adherence to treatment for OUD, attempts to synthesize and capture the landscape of research on the topic have not been made. This scoping review was conducted to provide an overview of different types of ML and AI applications used to predict OUD treatment adherence and evaluate their effectiveness and accuracy to identify models with the best predictive performance.

## Methods

### I. Approach and framework

Scoping reviews are a type of evidence synthesis that are considered the methodology of choice for comprehensively capturing the landscape of research on a topic, highlighting any gaps in knowledge [19]. This methodology was consistent with the objective of synthesizing extant literature on predicting OUD treatment adherence using ML and AI algorithms.

This scoping review was conducted in accordance with the Arksey and O’Malley (2005) framework which consists of 6 stages, including 1) developing a research question; 2) conducting a literature search; 3) screening studies against an inclusion and exclusion criteria; 4) extracting data from included articles; 5) analyzing abstracted data; and 6) a stakeholder consultation. The final step of the framework is considered optional [20] and thus, was not performed as a part of this review. The structure and content of this scoping review was guided by the preferred reporting items for systematic reviews and meta-analyses extension for scoping reviews (PRISMA-ScR) [21]. The protocol for this review was registered on Open Science Framework (OSF) [22].

### II. Search strategy, study selection, and screening

A comprehensive search strategy was developed using keywords such as artificial intelligence, opioid, treatment adherence, and compliance that would capture articles on the use of ML and AI models in predicting treatment adherence for OUD. The search was run across Ovid MEDLINE, Embase, PsycINFO, Web of Science, Scopus, CINAHL, IEEE Xplore, and ACM Digital Library from inception to October 7, 2024. The full search strategy is present in the supplementary material.

Articles obtained from the search results were uploaded onto Covidence (Veritas Health Innovation, Melbourne, Australia; available at www.covidence.org). The title and abstracts of the retrieved studies were reviewed to determine their eligibility against the inclusion criteria. Studies that were included after title and abstract screening were further assessed through a full-text review. Articles were screened independently by two reviewers at both screening stages and disagreements were resolved by a third reviewer. Peer-reviewed, primary and secondary empirical studies on ML and AI applications to predict treatment adherence in individuals with OUD were deemed eligible. Relevant conference papers were included only if they were from IEEE or ACM. There were no restrictions on study setting, date of publication, or language. Articles were excluded if they did not include predictive ML and AI models or assess outcomes of interest. Moreover, studies with non-human participants were considered ineligible. Non-research articles (e.g., erratums, commentaries, book chapters, letters to the editor, and protocols), thesis dissertations, and reviews were also eliminated.

### III. Data extraction and analysis

Data extraction was completed on Covidence using a standardized template populated independently by a pair of researchers for each study. The abstracted information was reviewed for accuracy by a third researcher and conflicts were resolved through consensus. The following information was collected: 1) title of the article; 2) primary author; 3) publication date; 4) aims of the study; 5) country where the study was conducted; 6) study design; 7) location or site; 8) sample size; 9) sociodemographic characteristics (e.g., age, gender, race or ethnicity, medical history or comorbidities); 10) description of outcomes; 11) how outcomes were measured (*e.g*., surveys, health data records, questionnaires); 12) length of data collection; 13) description of treatment intervention; 14) ML and AI models discussed; 15) source of input data (*e.g.*, ECG, electronic health records); 16) predictive or modelling variables; 17) size of training and testing dataset; 18) performance metrics (*e.g*., 5-fold cross validation, random sampling); 19) best model and its evaluation metrics (*e.g*., accuracy, precision, recall, sensitivity, specificity, positive predictive value); 20) key findings of the study; and 21) its limitations. The extracted data was analyzed and reported using a narrative synthesis approach.

## Results

### Search results

The database search yielded 2800 hits. Of these 2729 articles were removed at the title and abstract screening stage with another 49 articles excluded following full-text review. A total of 22 articles were deemed eligible and included in this scoping review (Figure 1).

**Figure 1:**
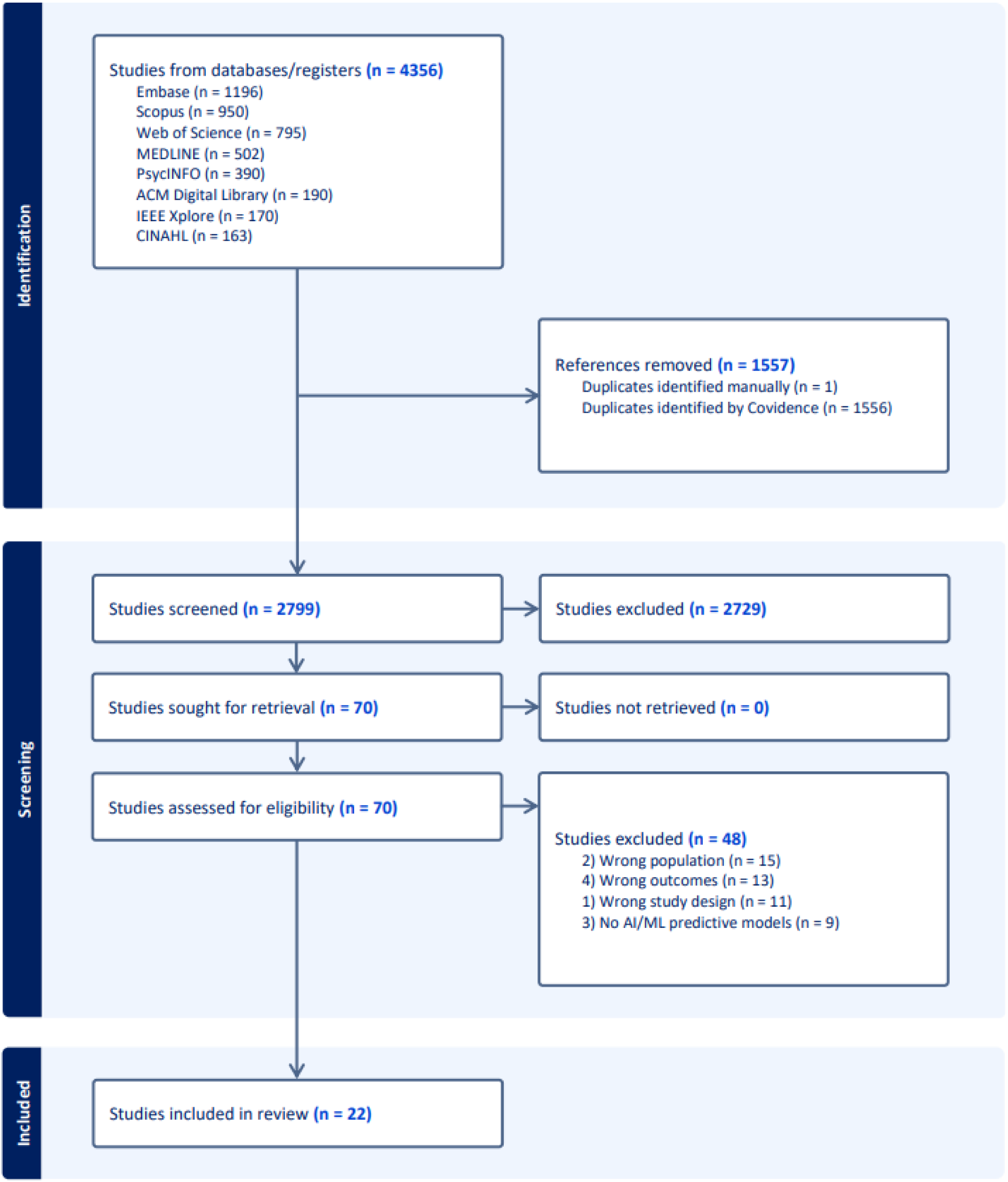
Preferred Reporting Items for Scoping reviews and Meta-Analysis (PRISMA) flow chart for the study

### Results of individual sources of evidence

The study characteristics and main findings of each of the 22 included articles are summarized in Tables 1-2.

**Table 1.**
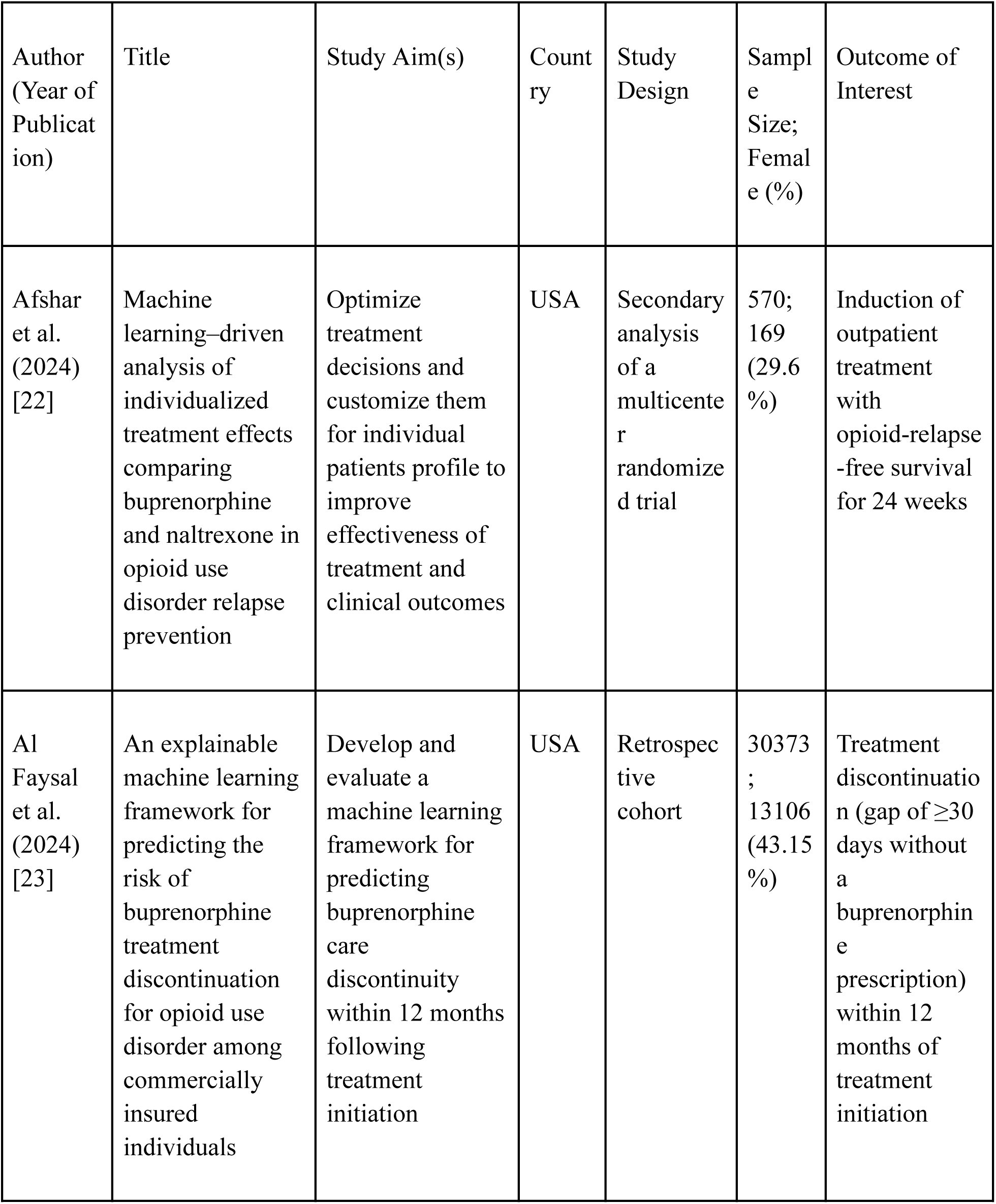
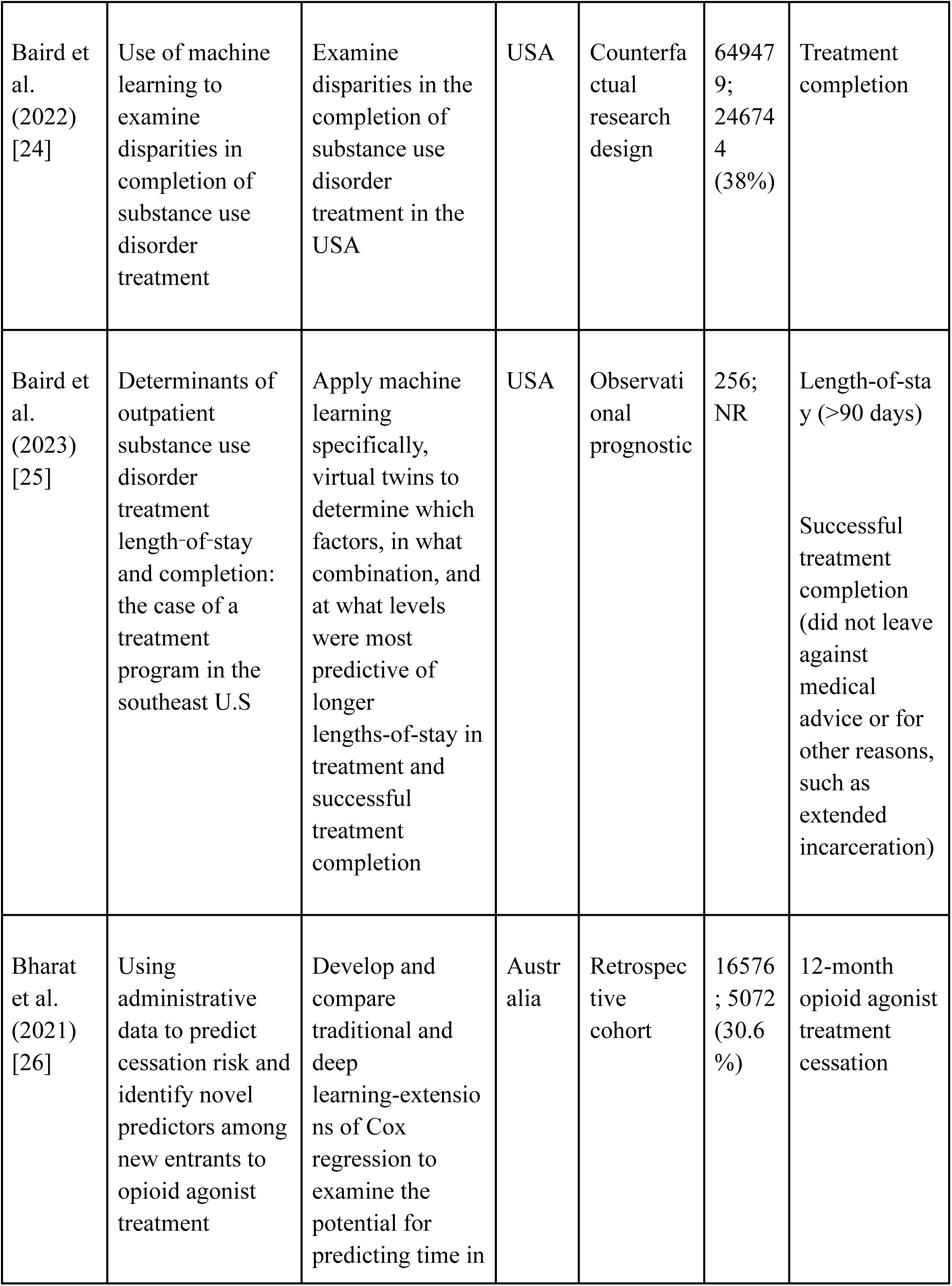
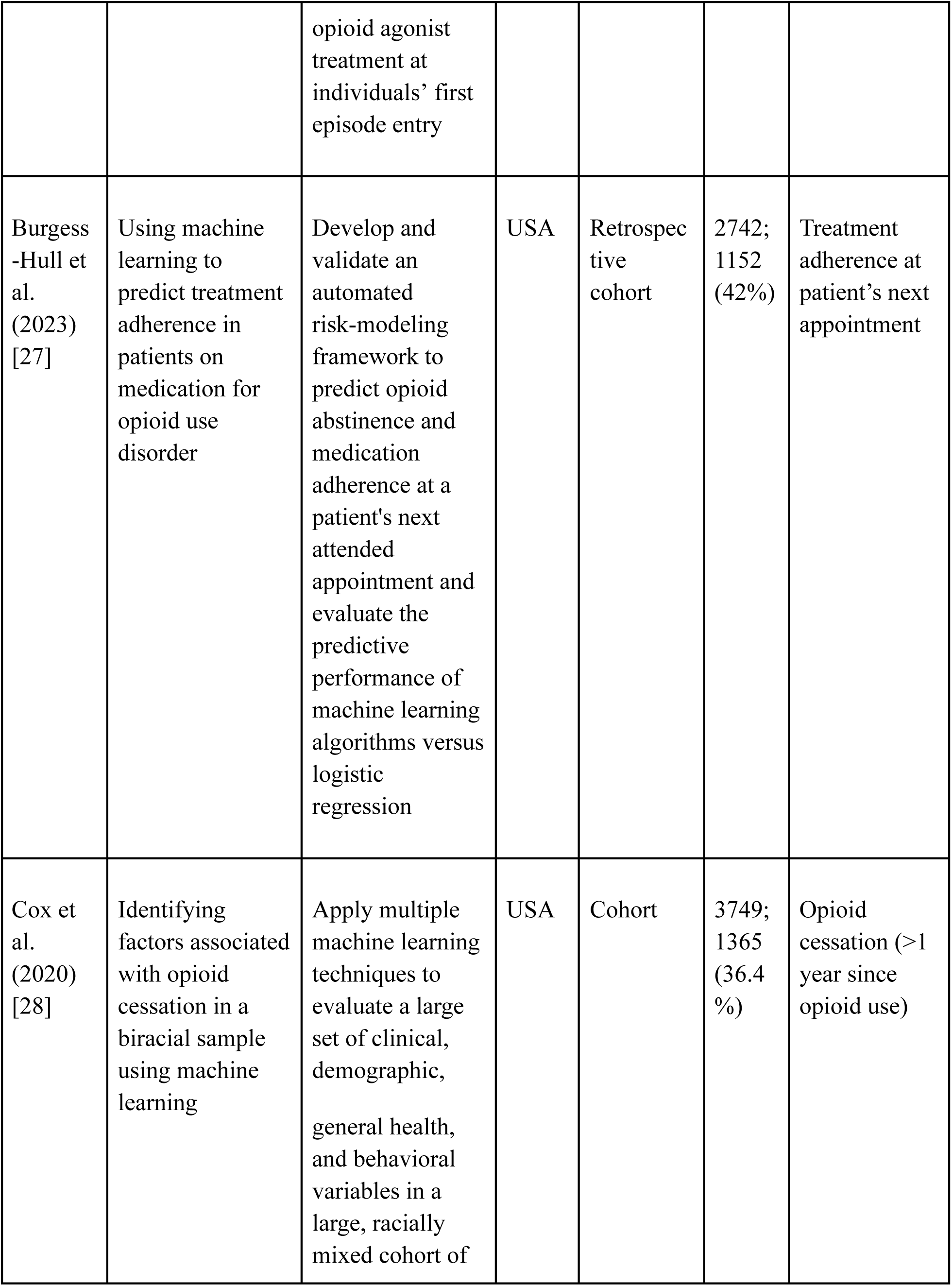
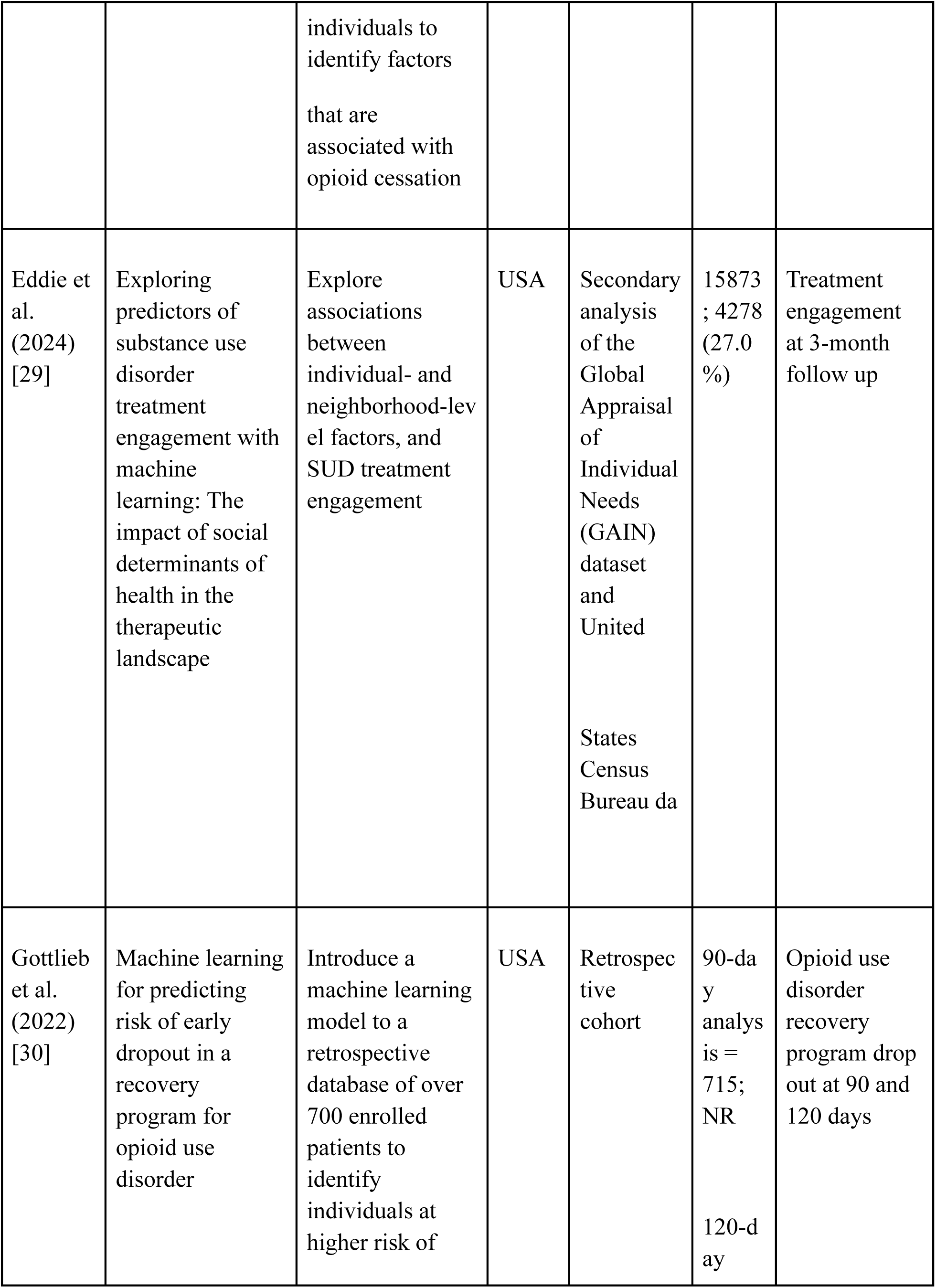
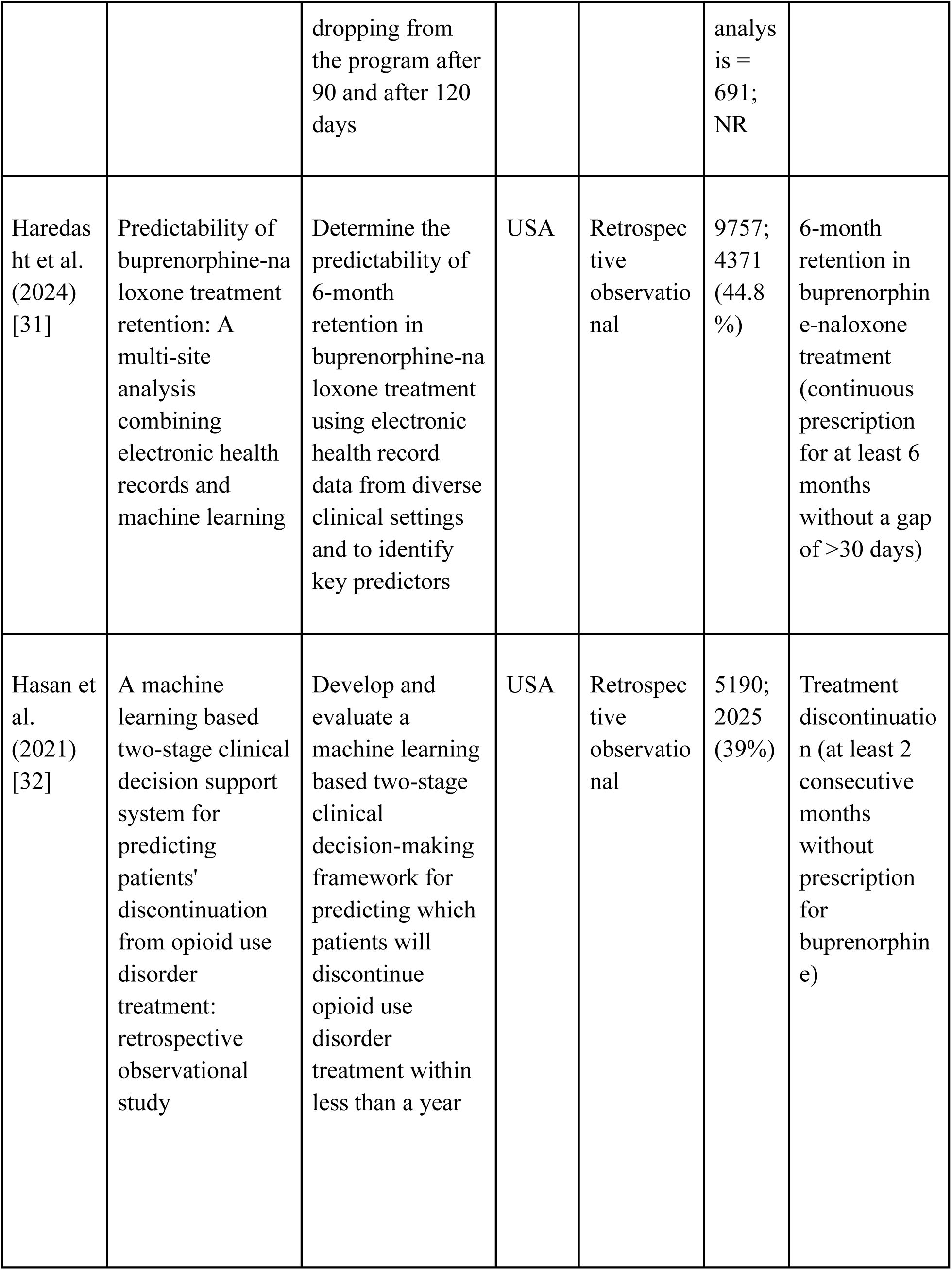
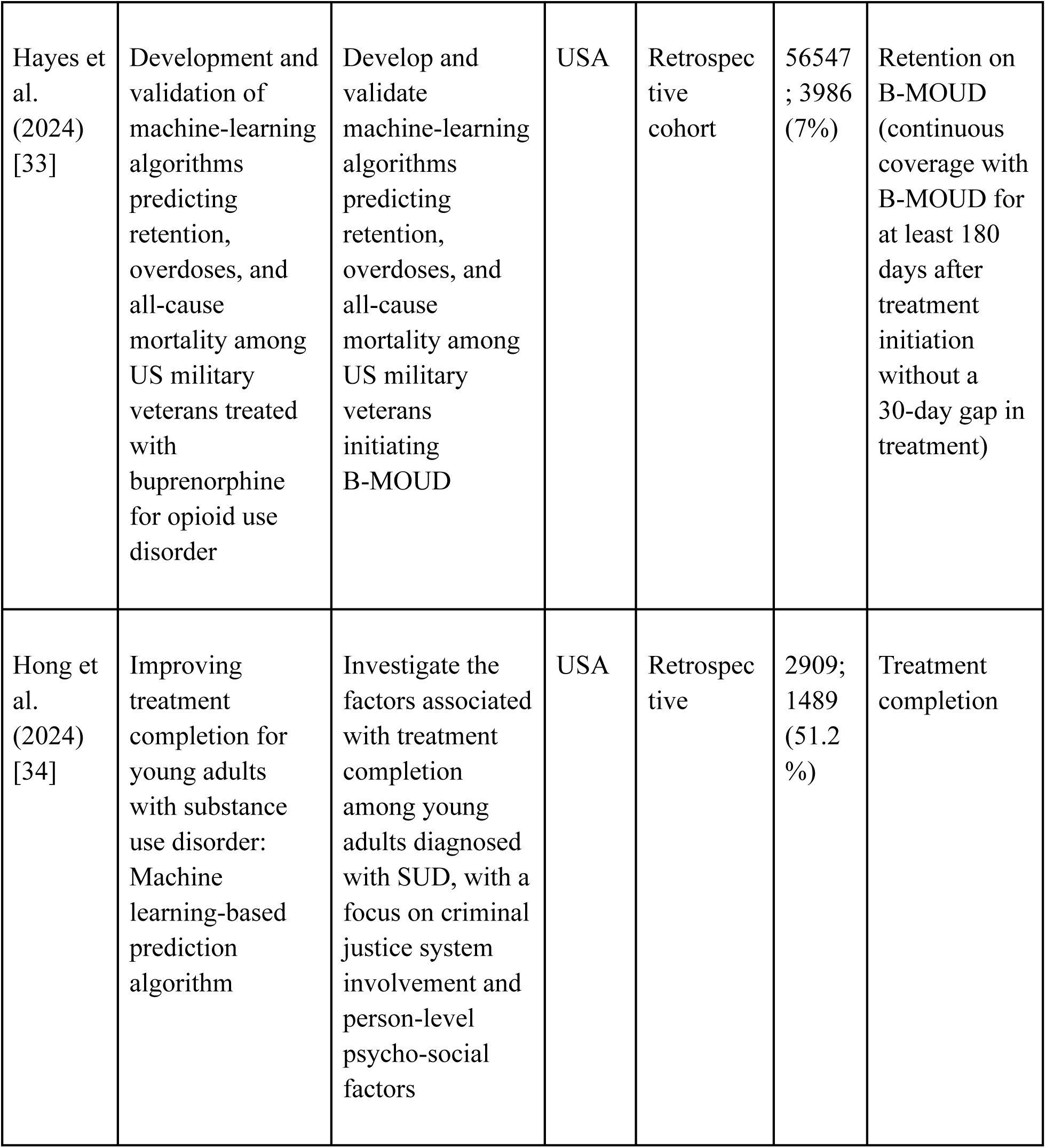
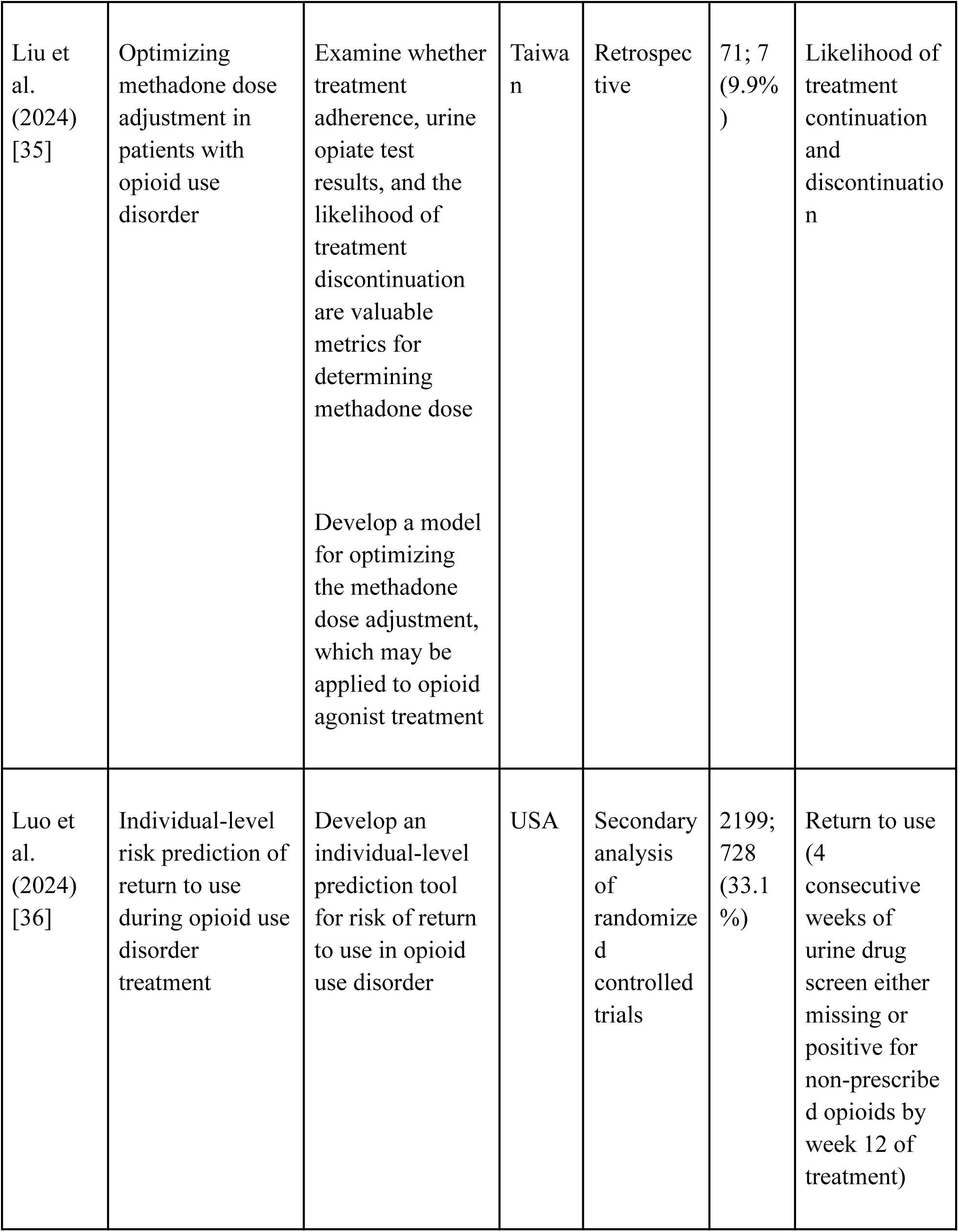
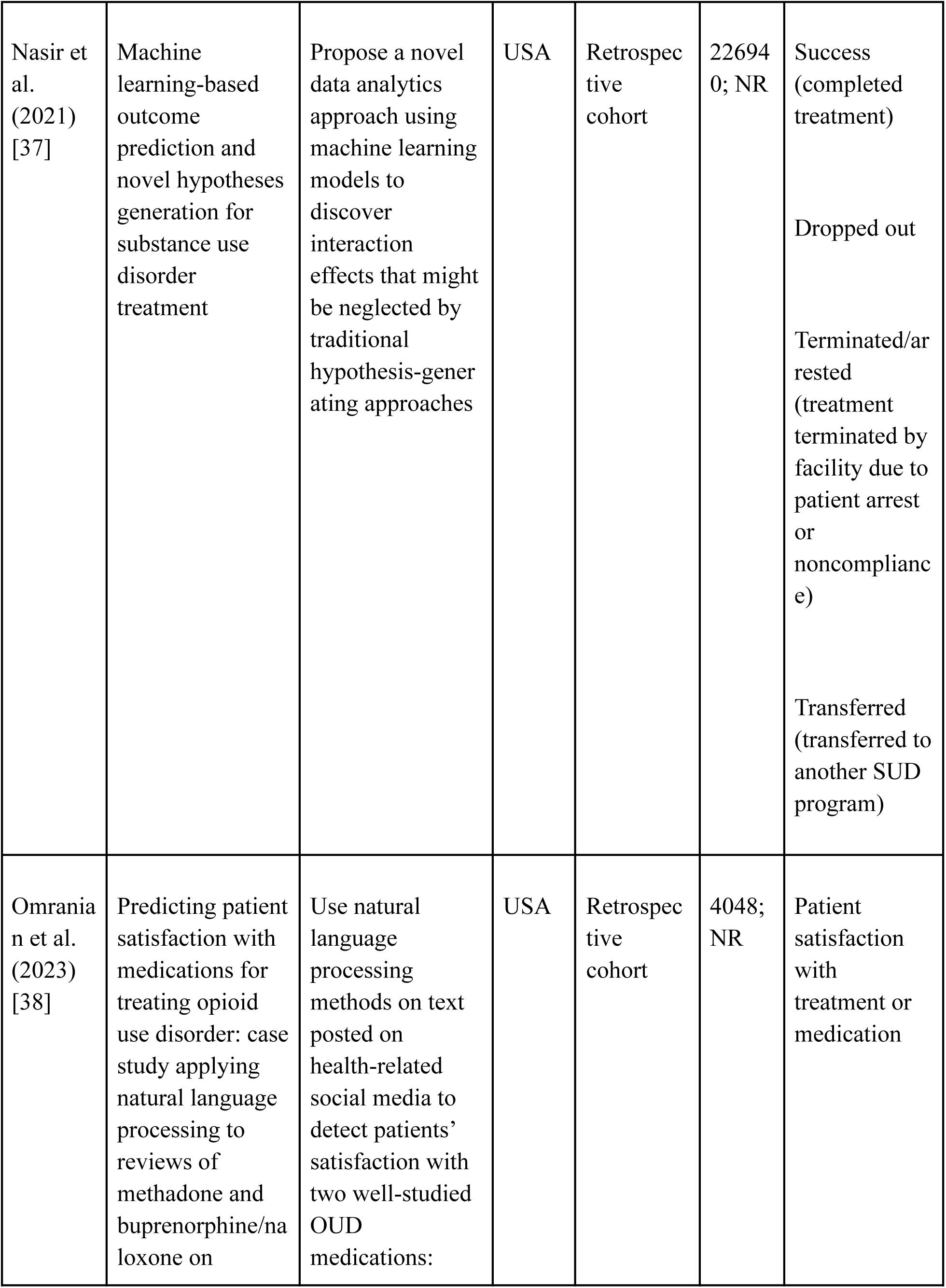
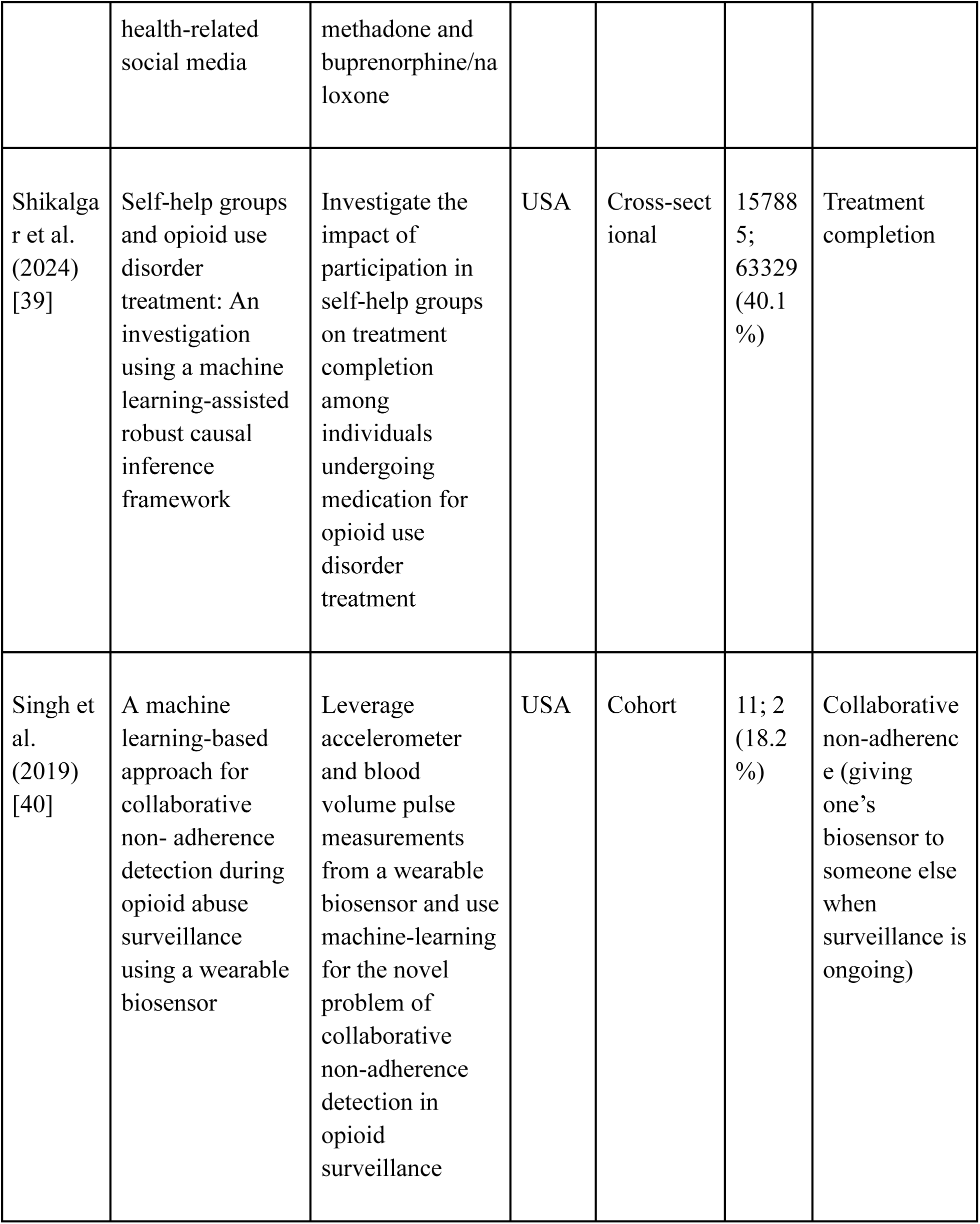
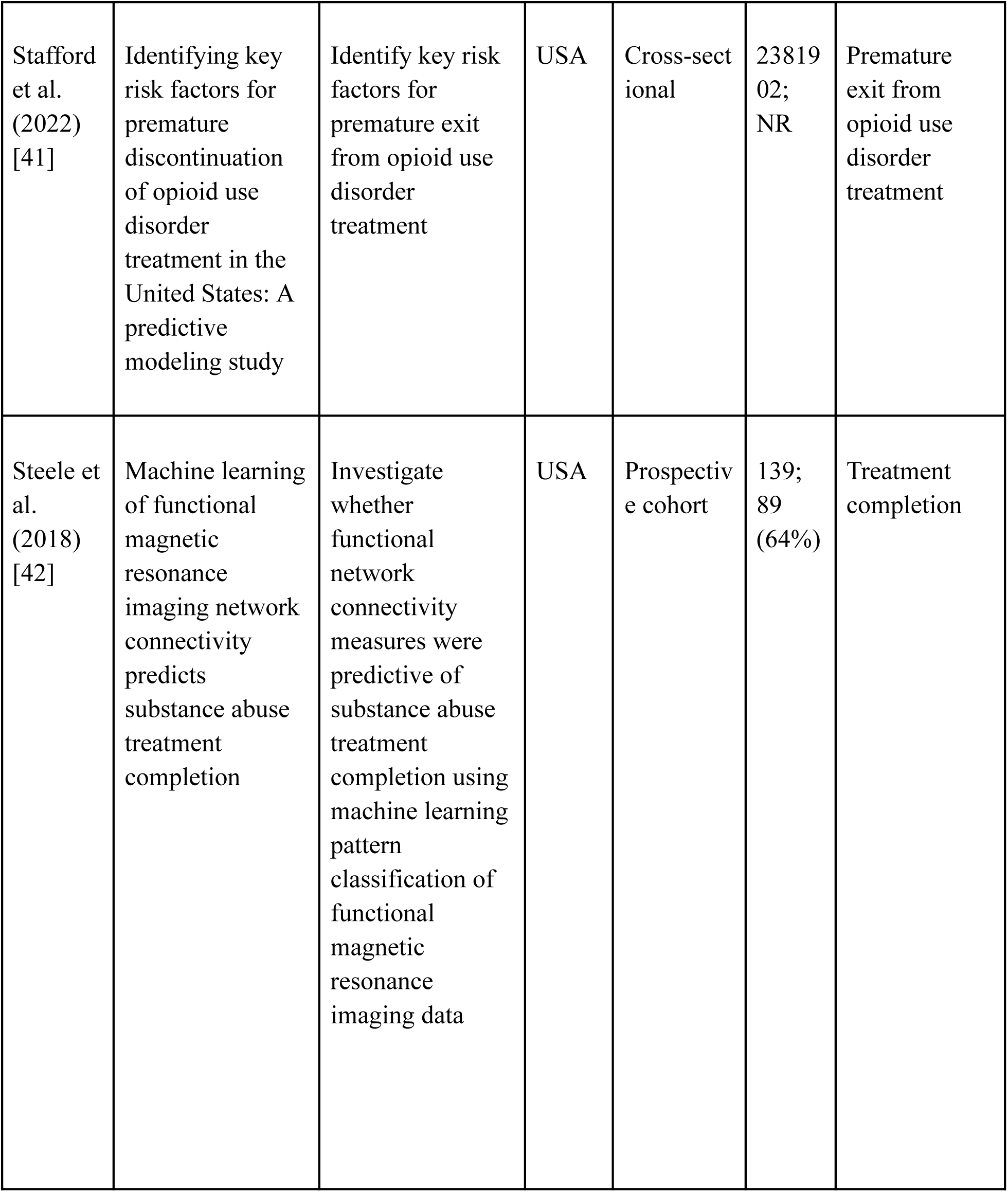
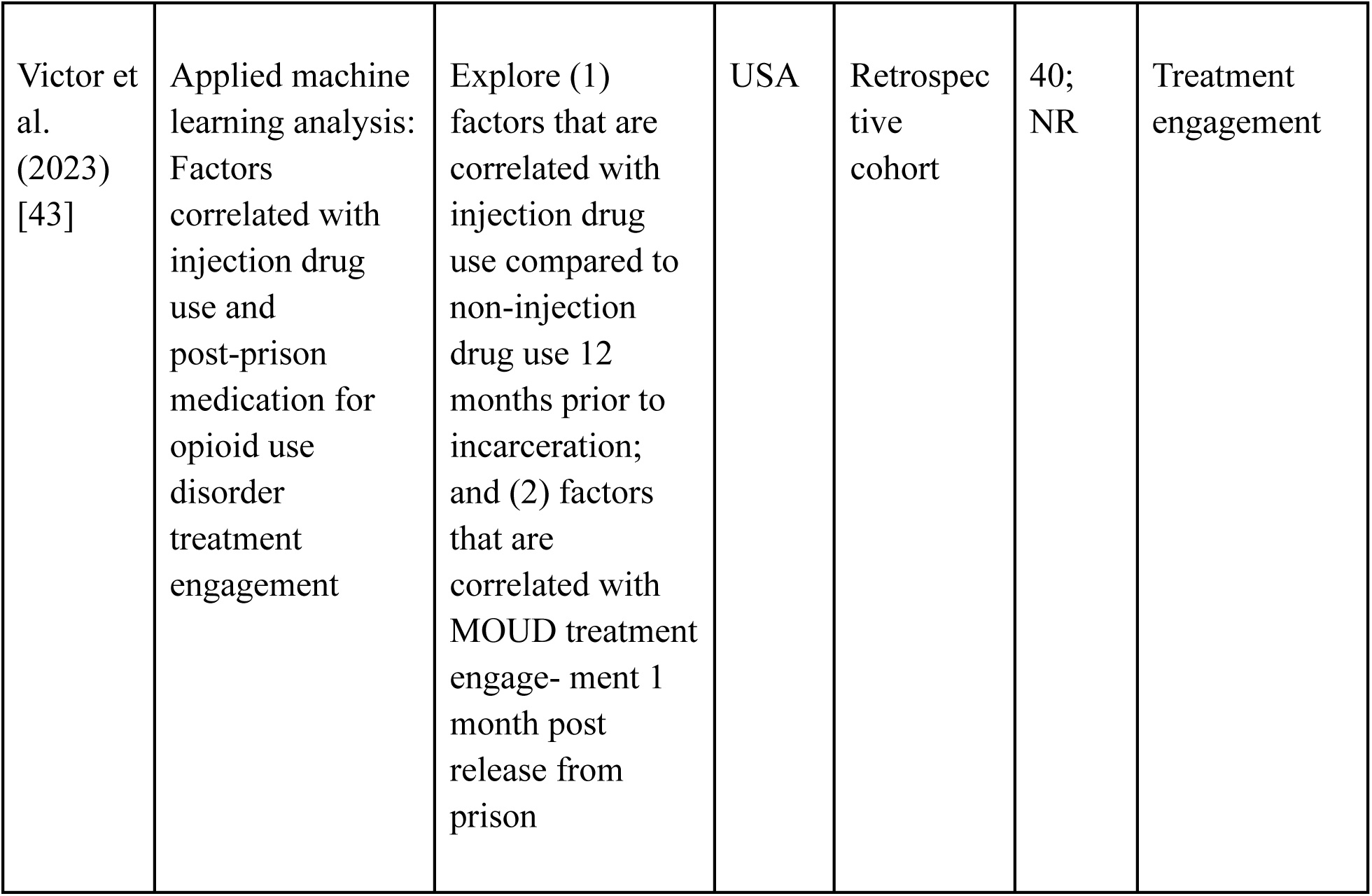
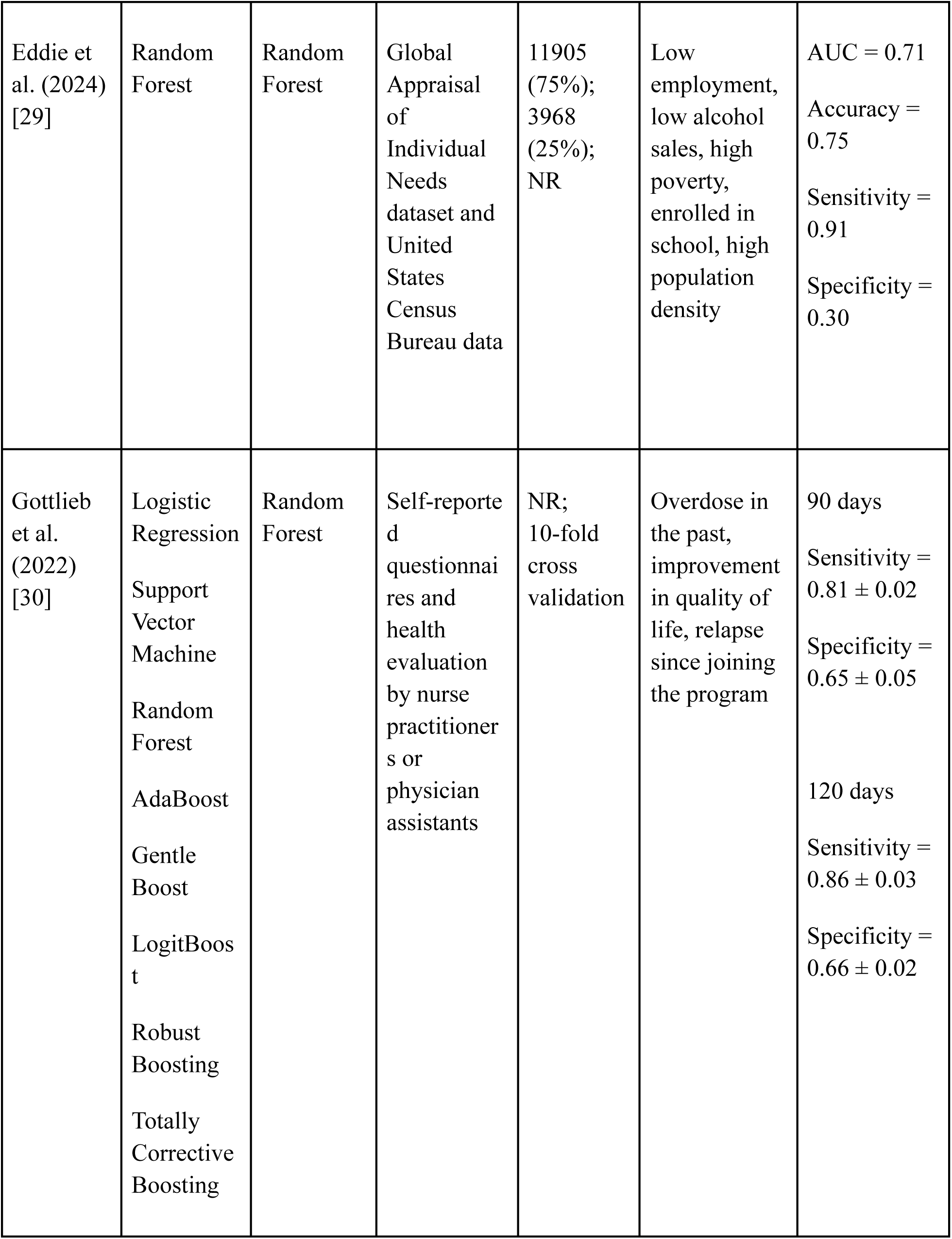
Summary of study characteristics.

**Table 2.**
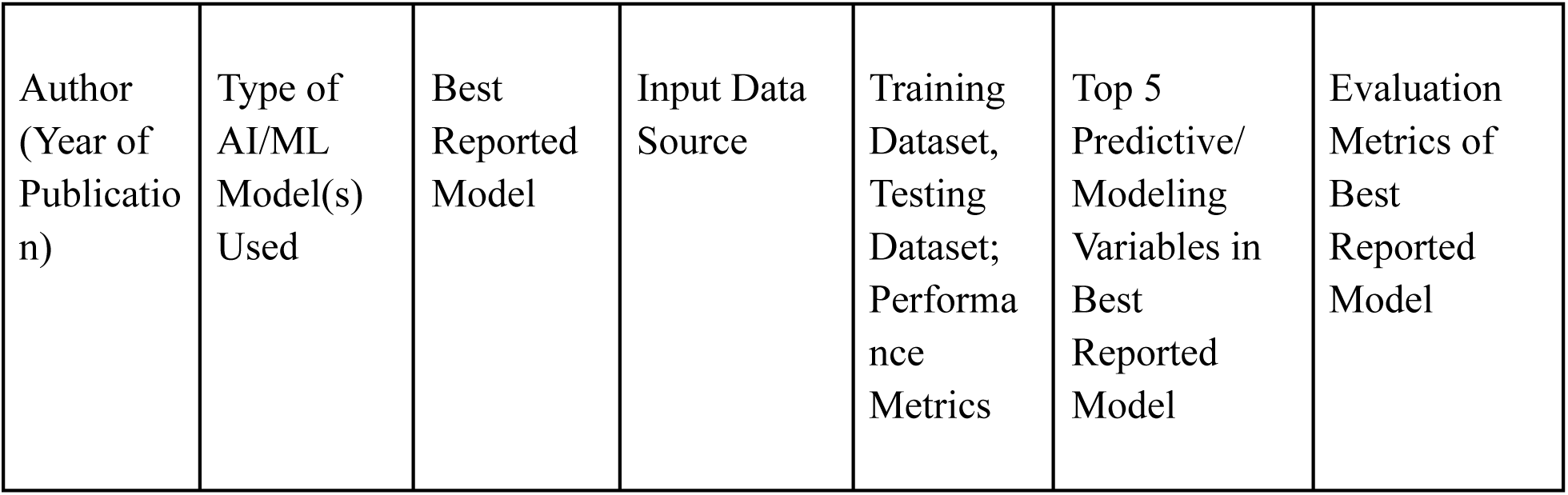
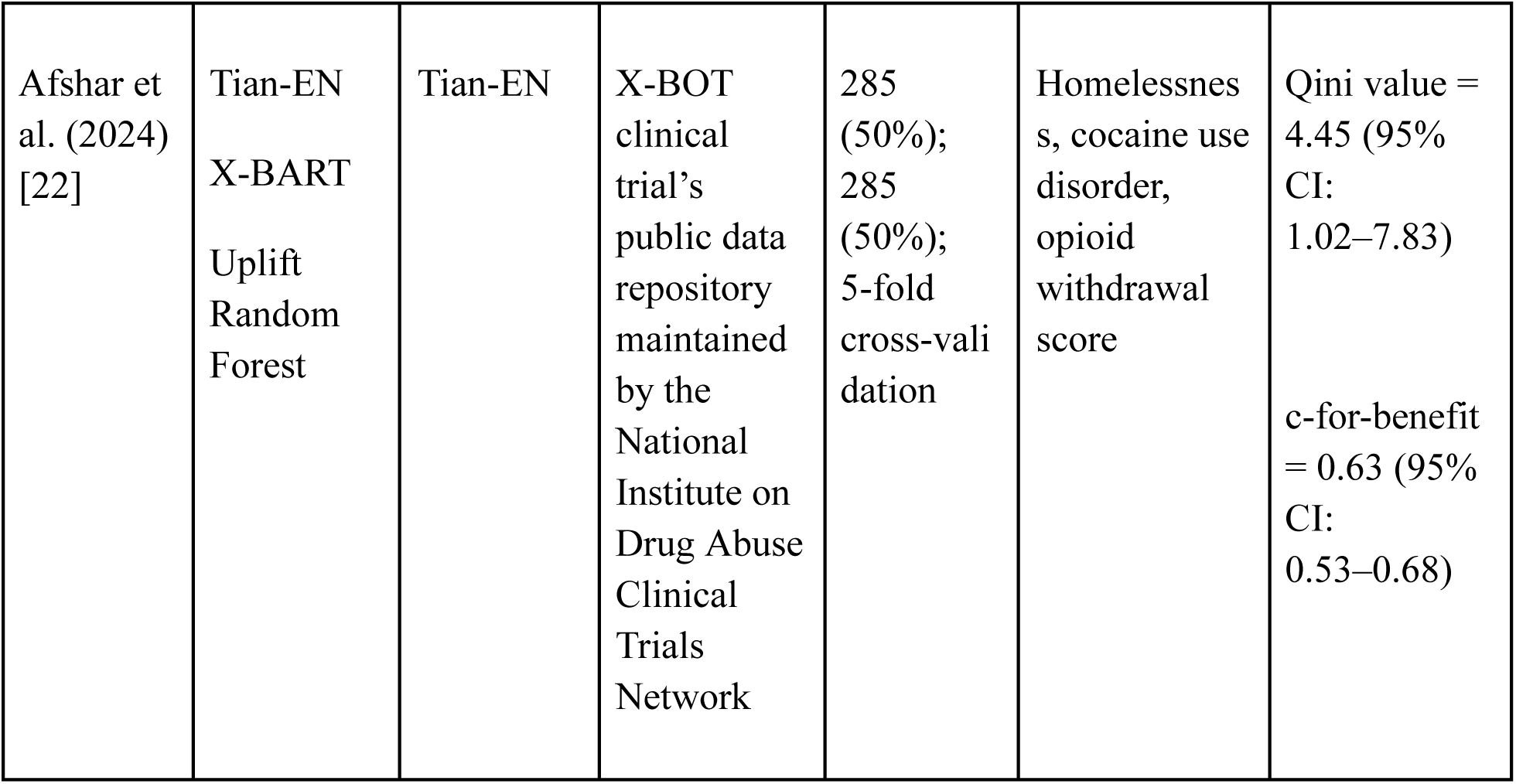
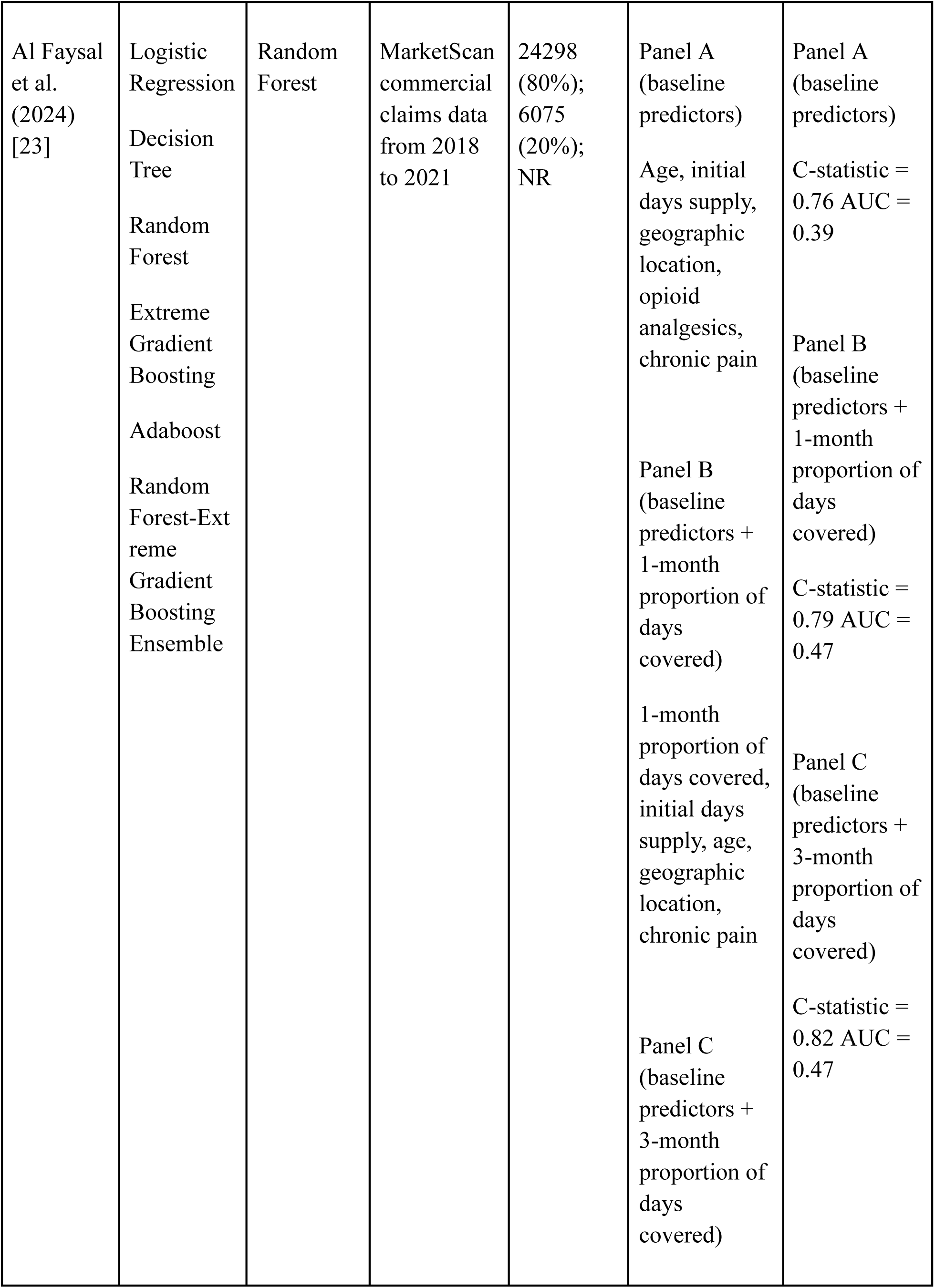
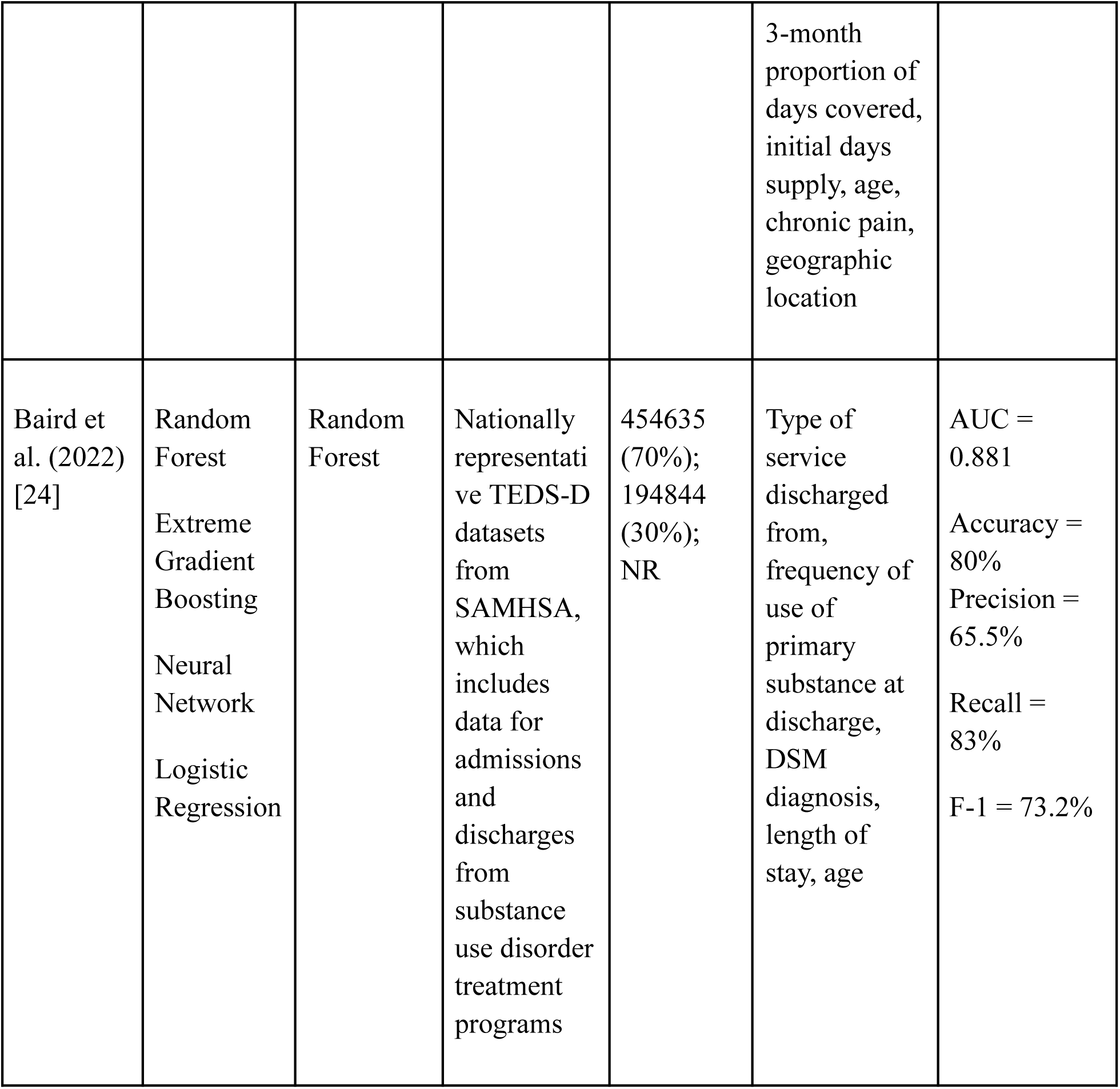
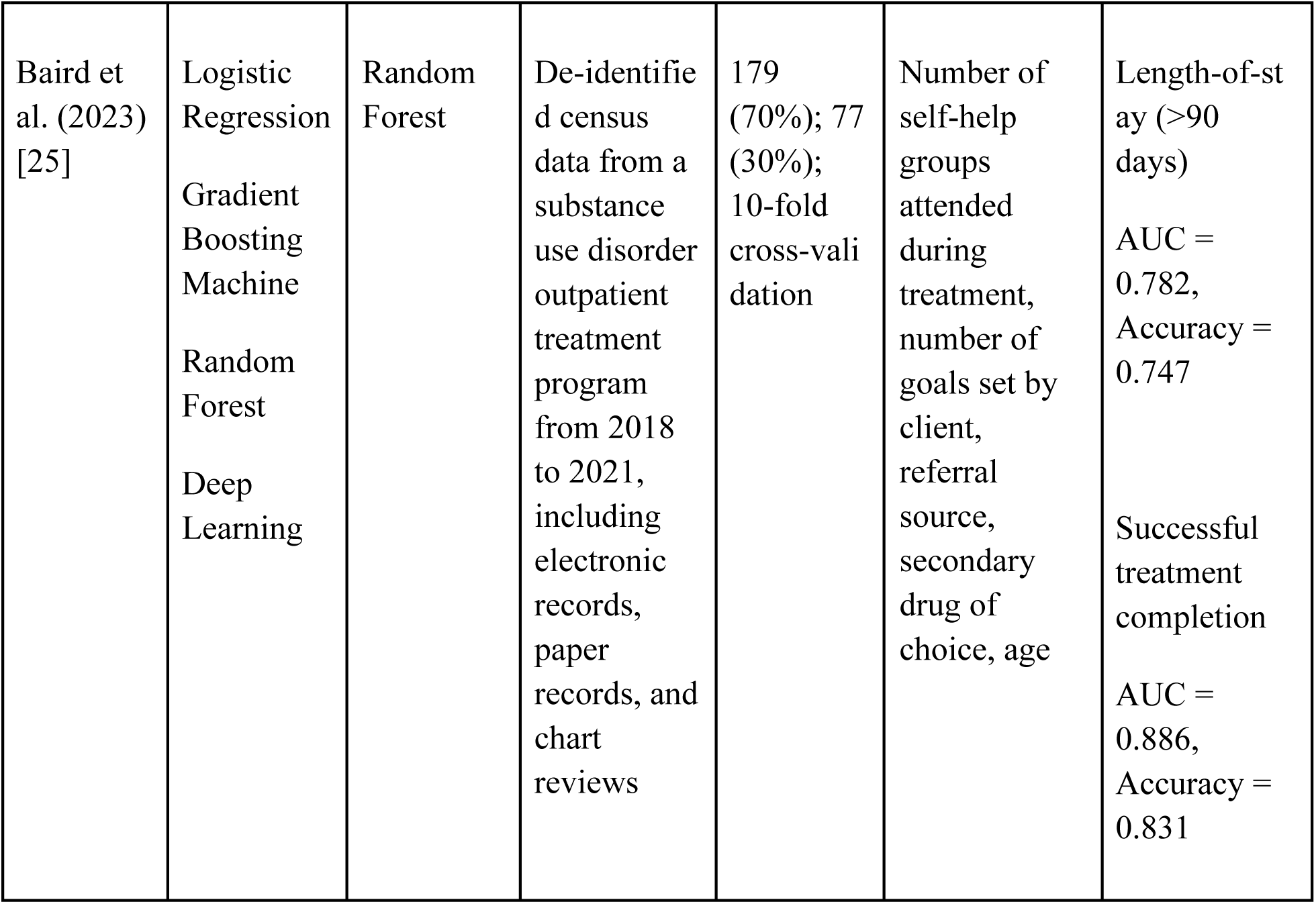
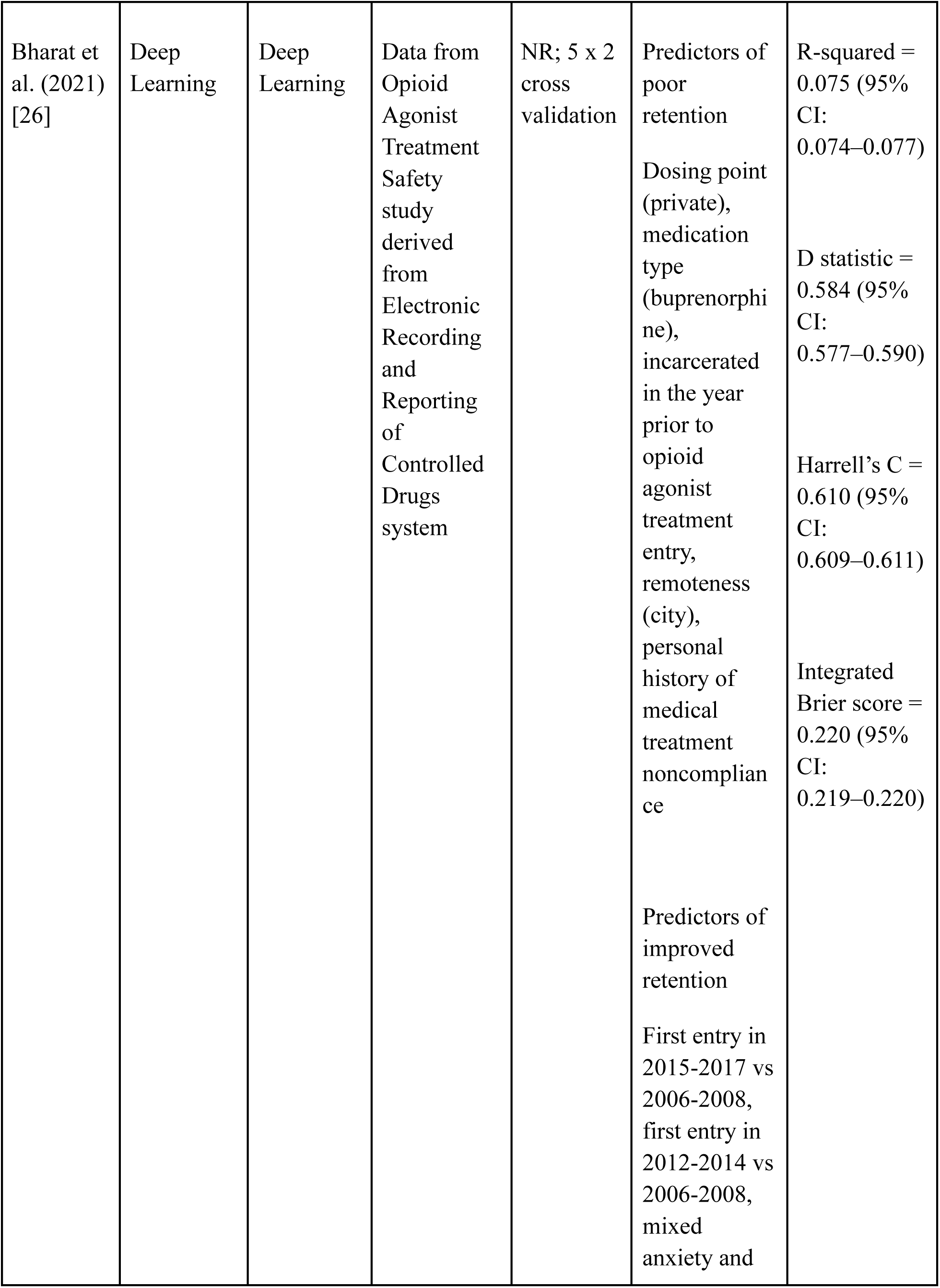
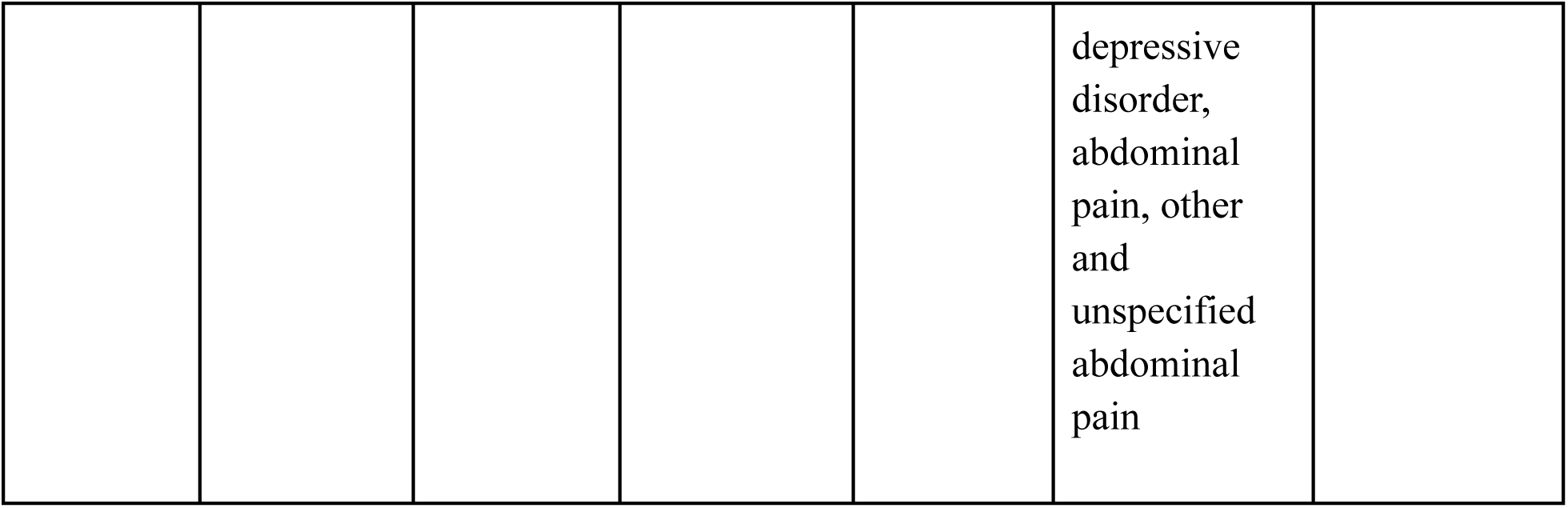
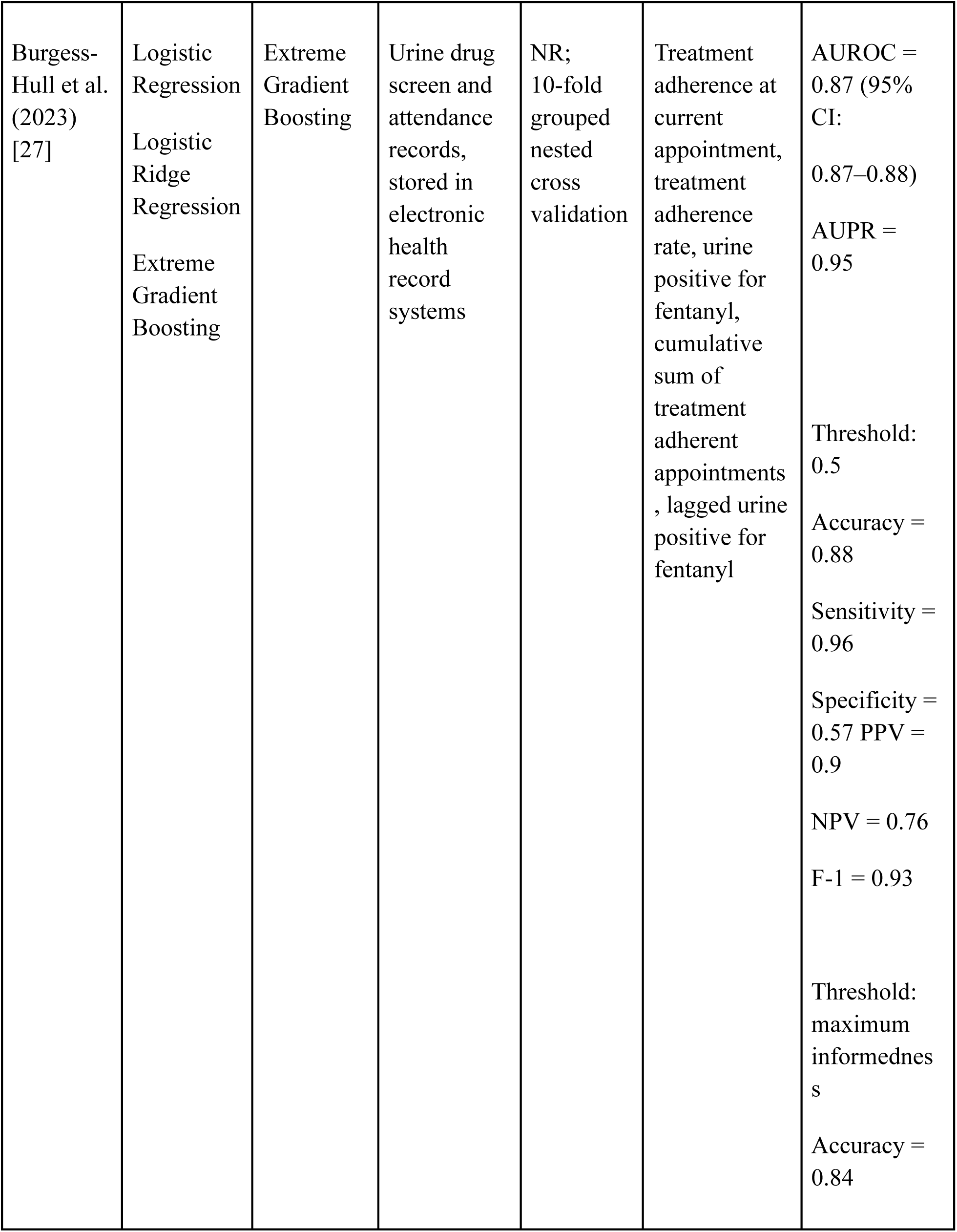
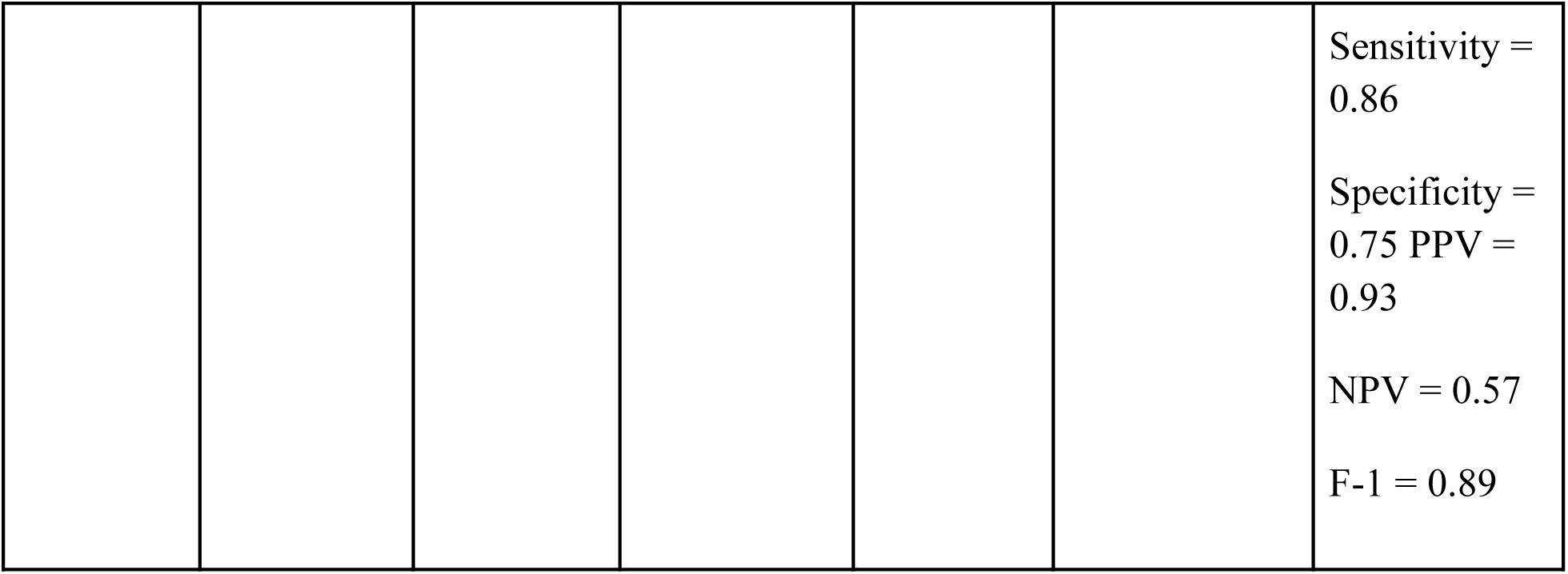
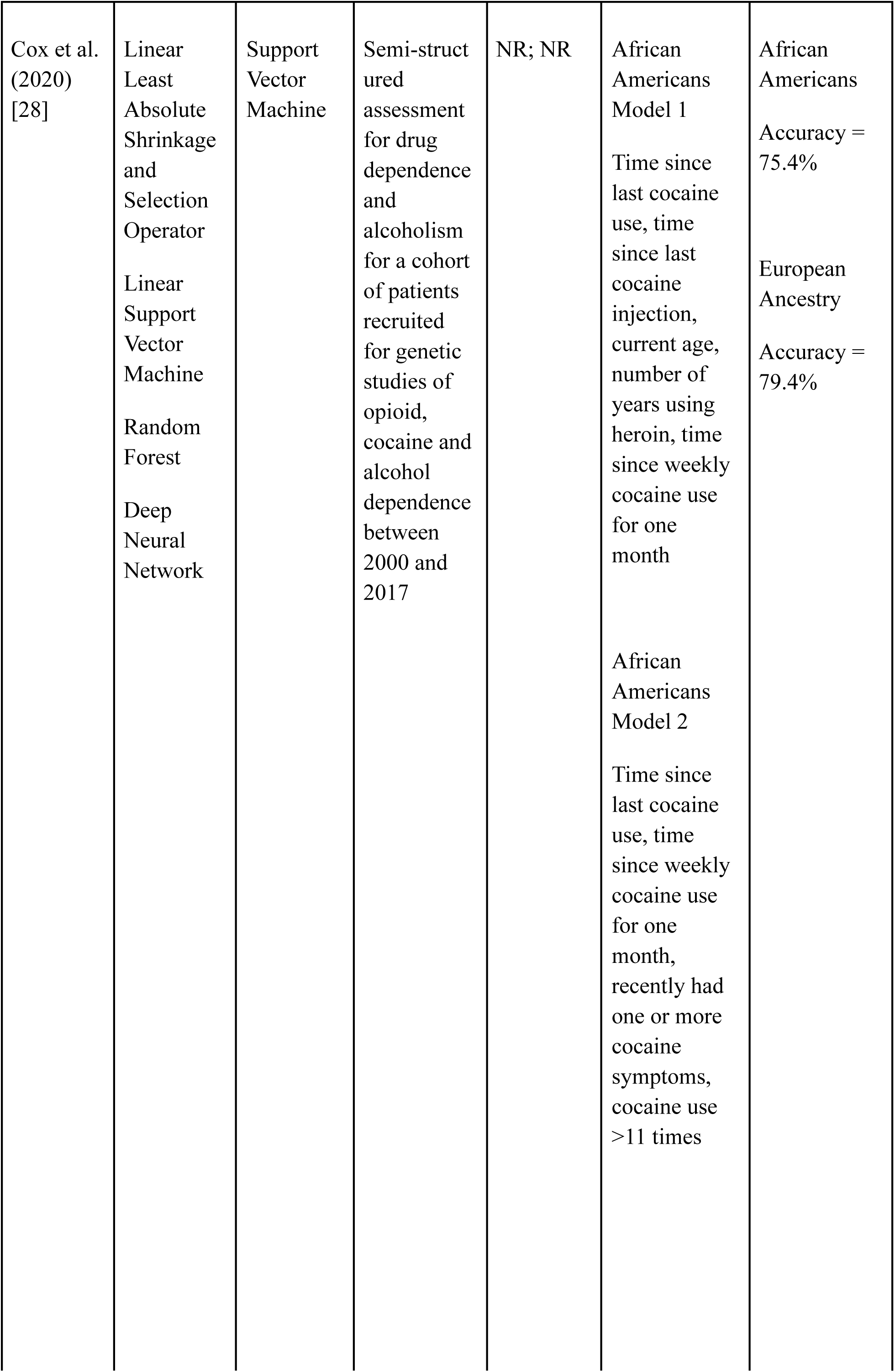
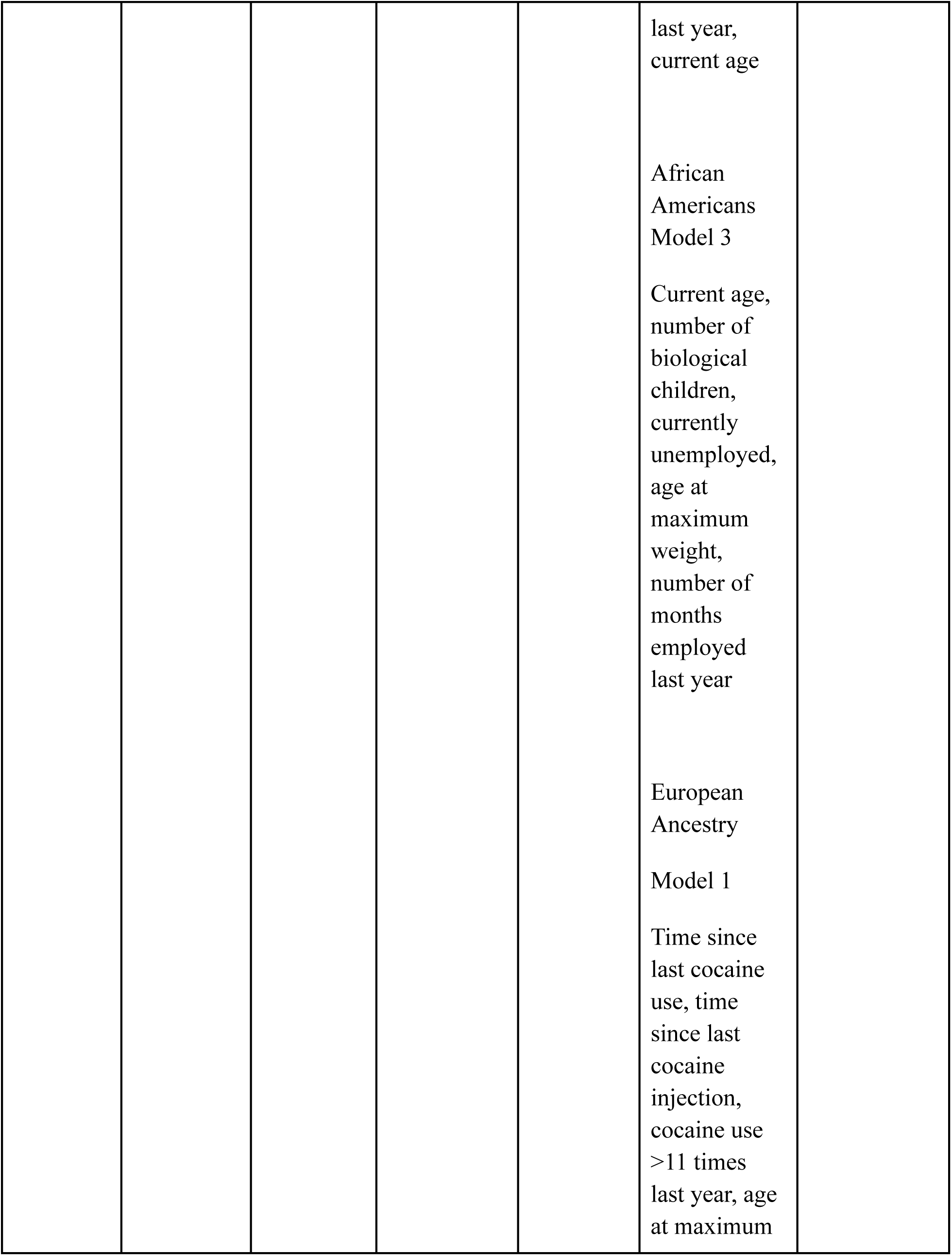
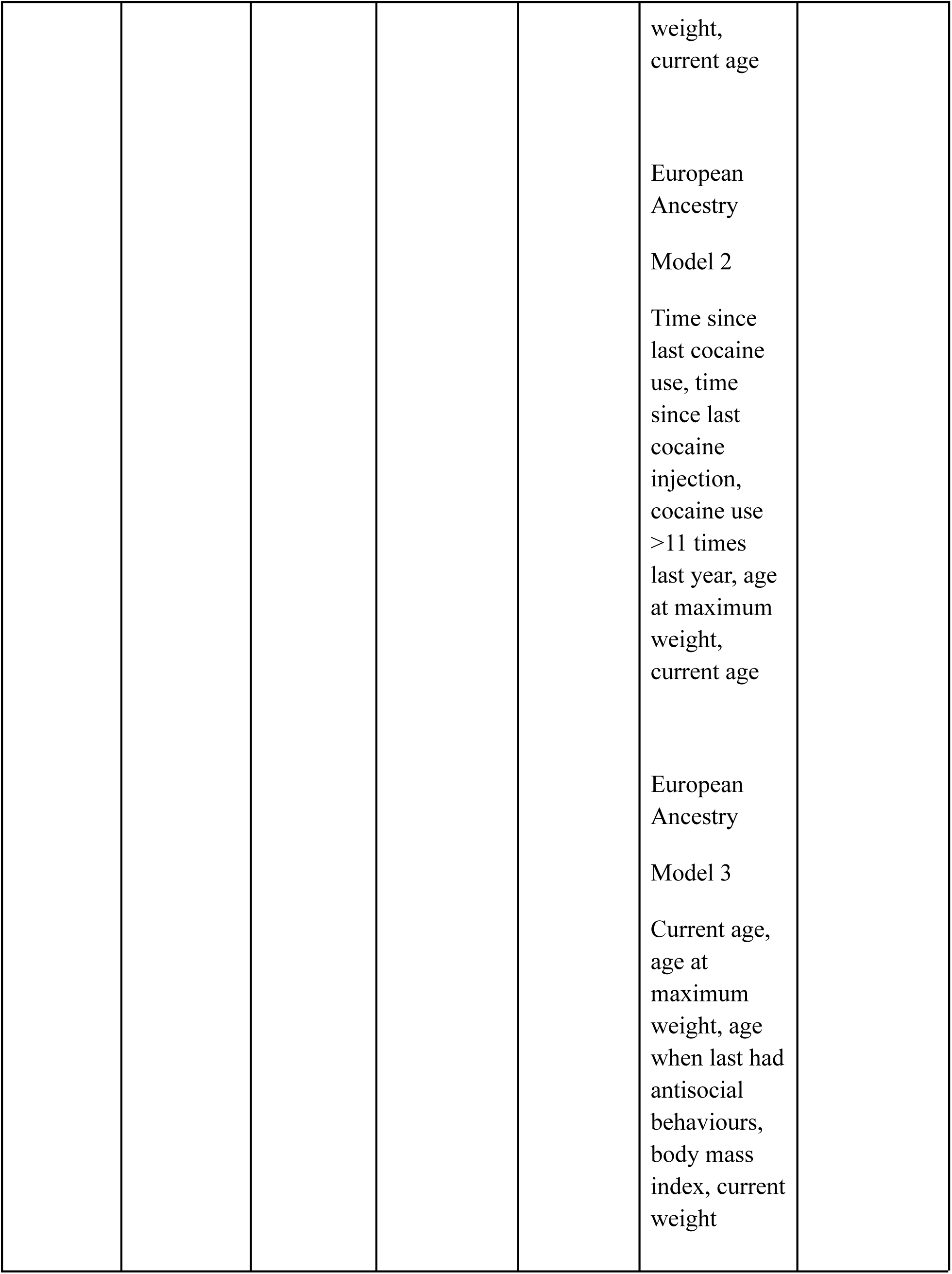
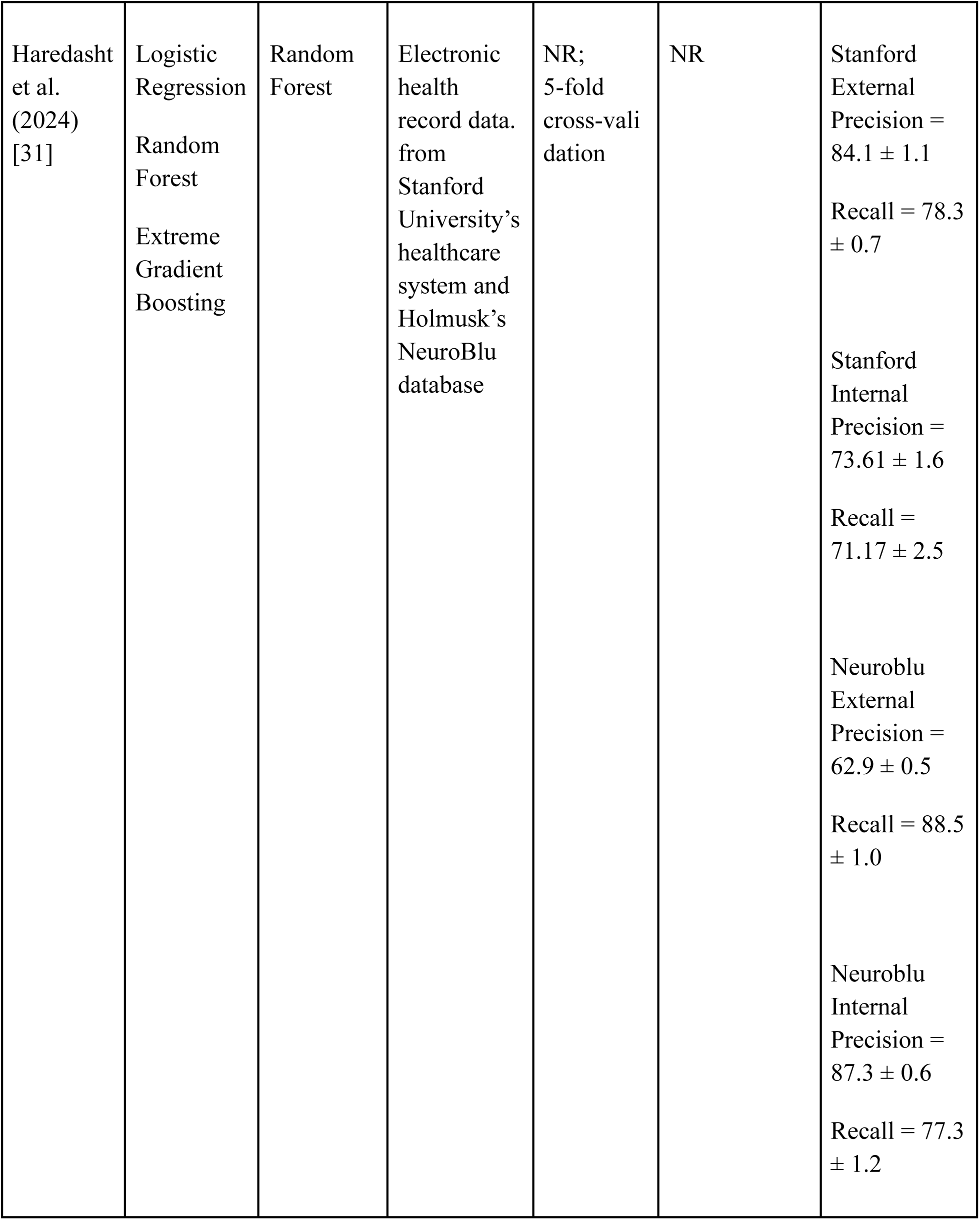
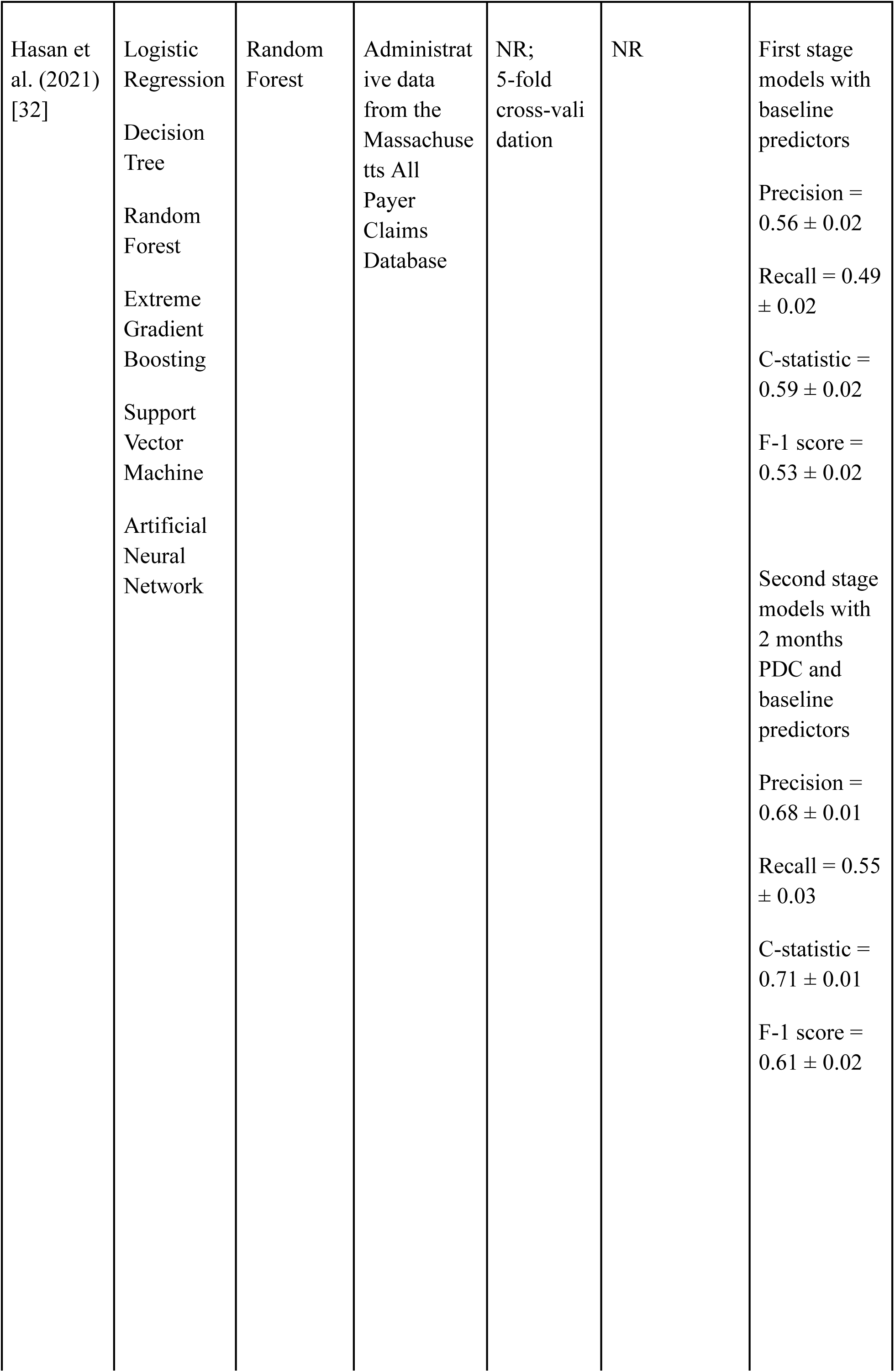
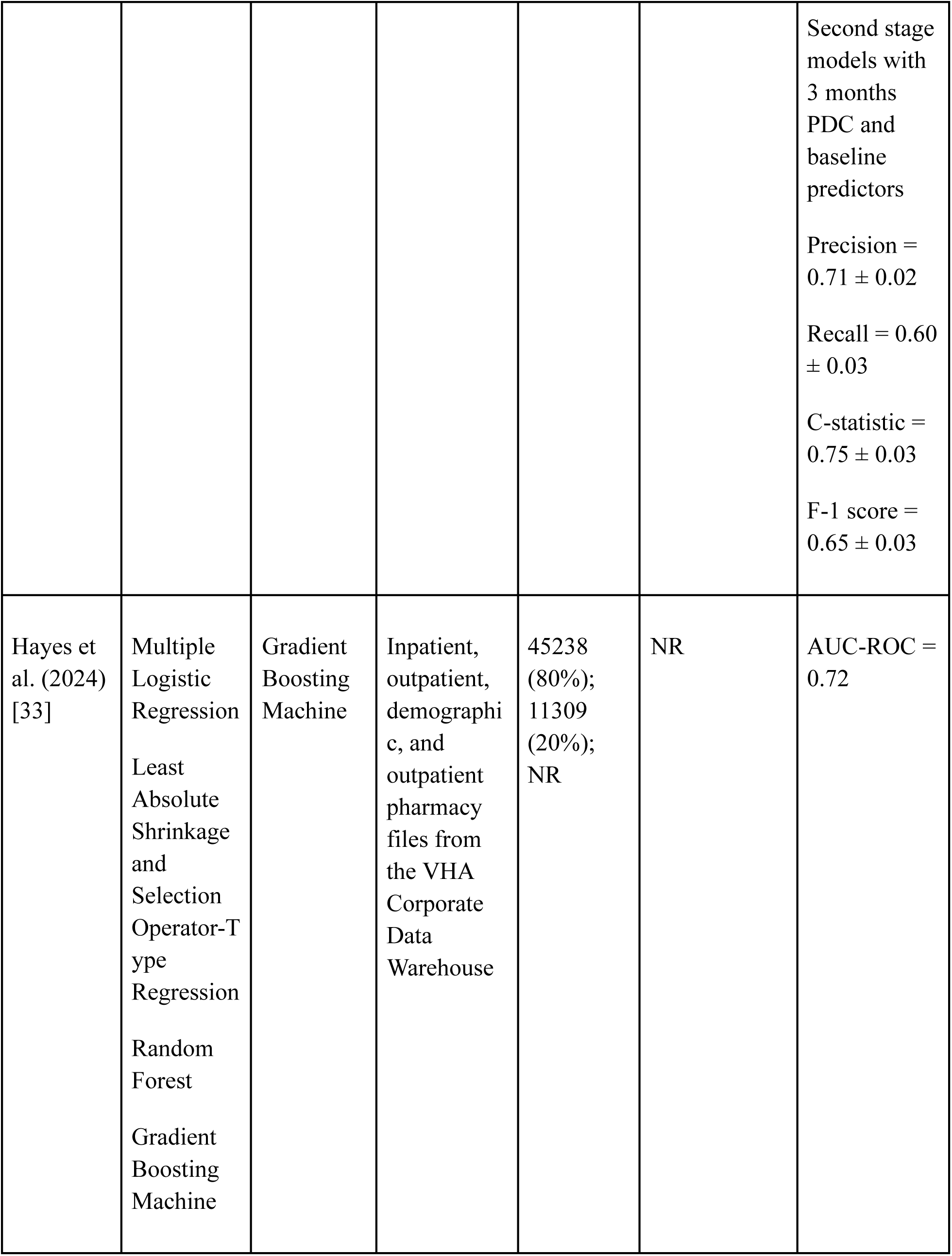
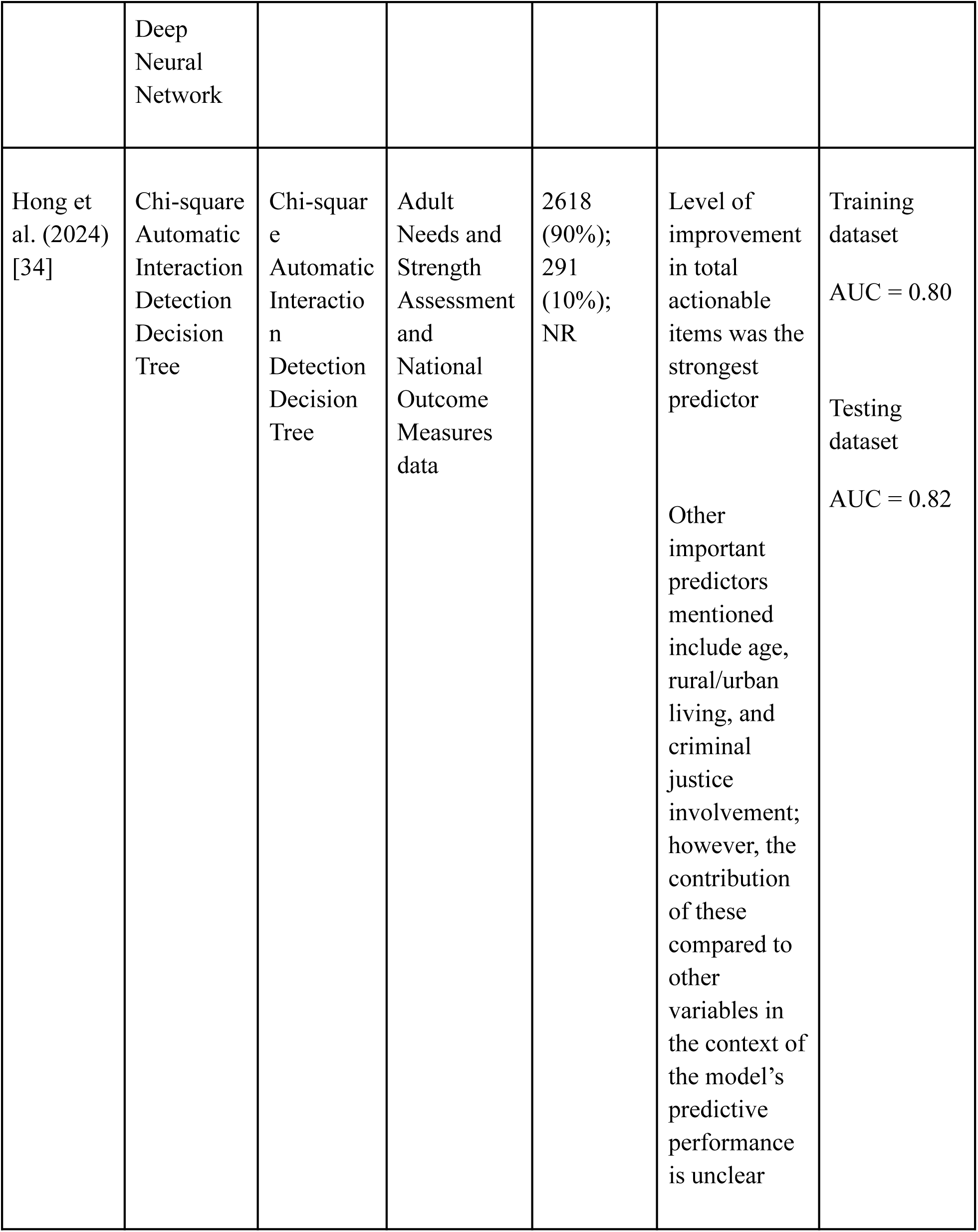
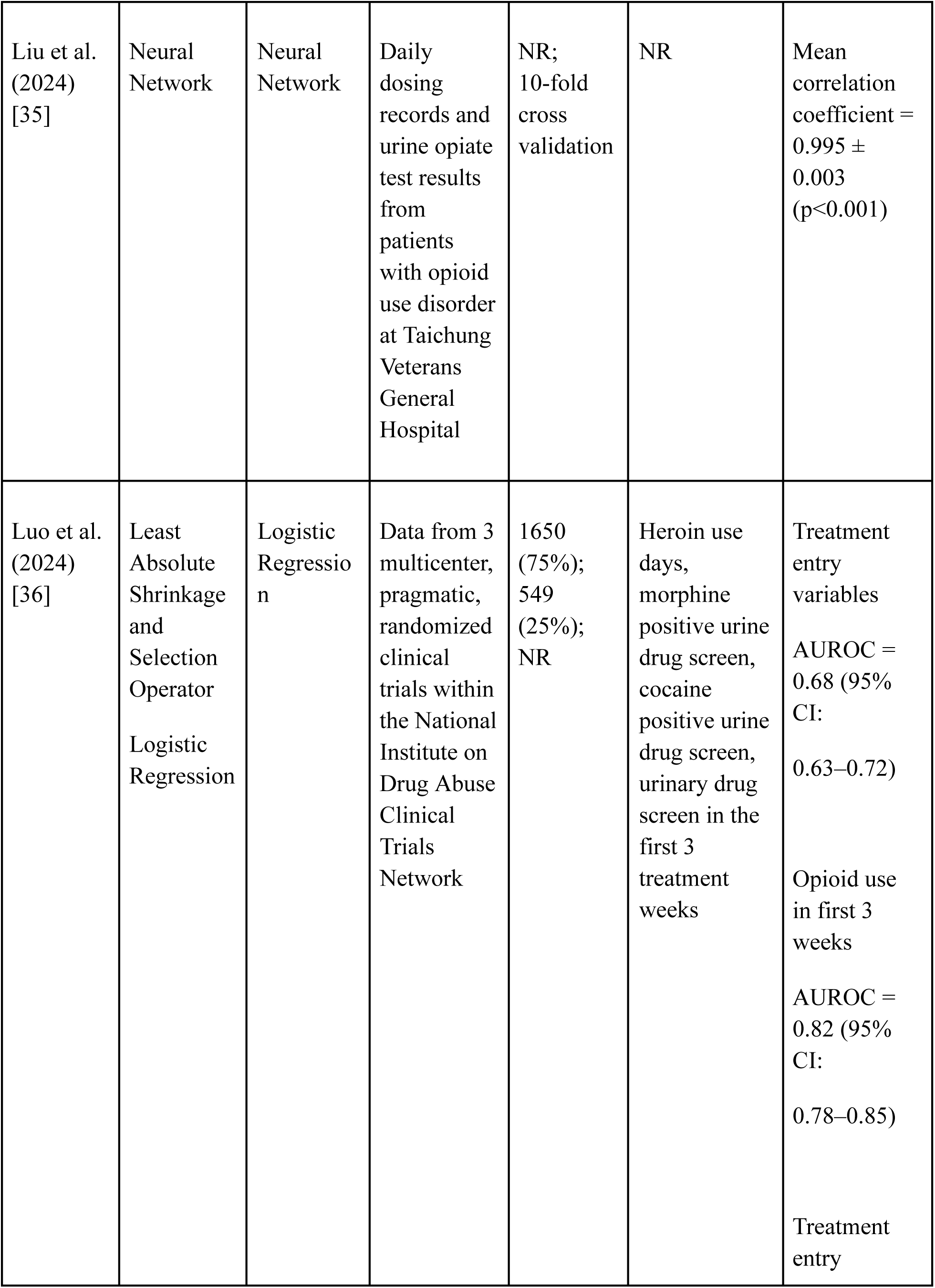
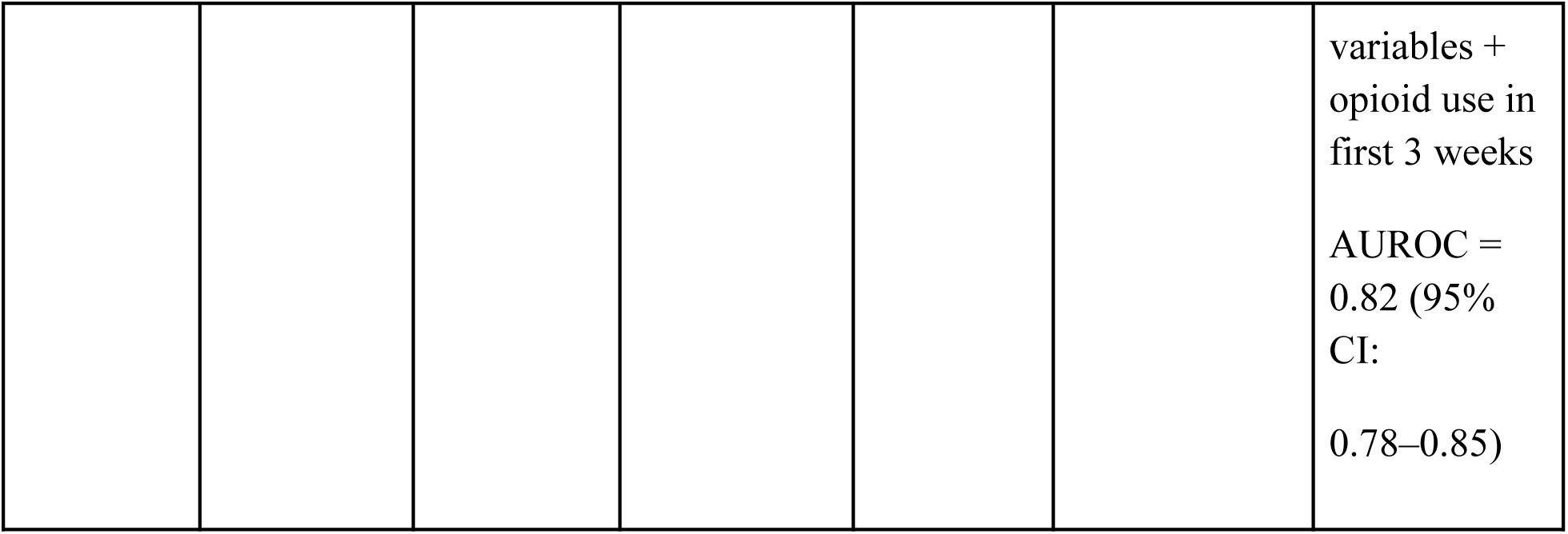
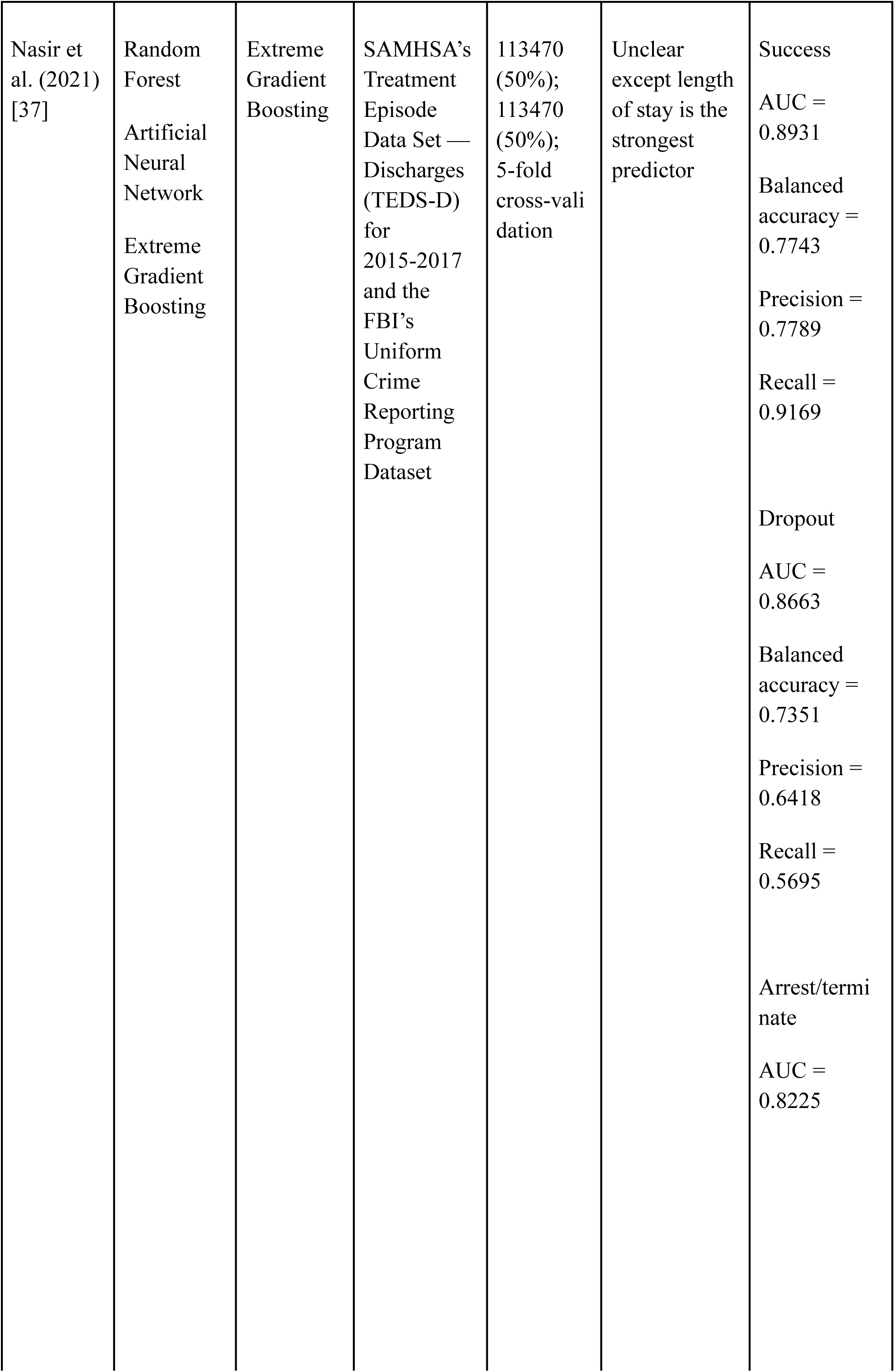
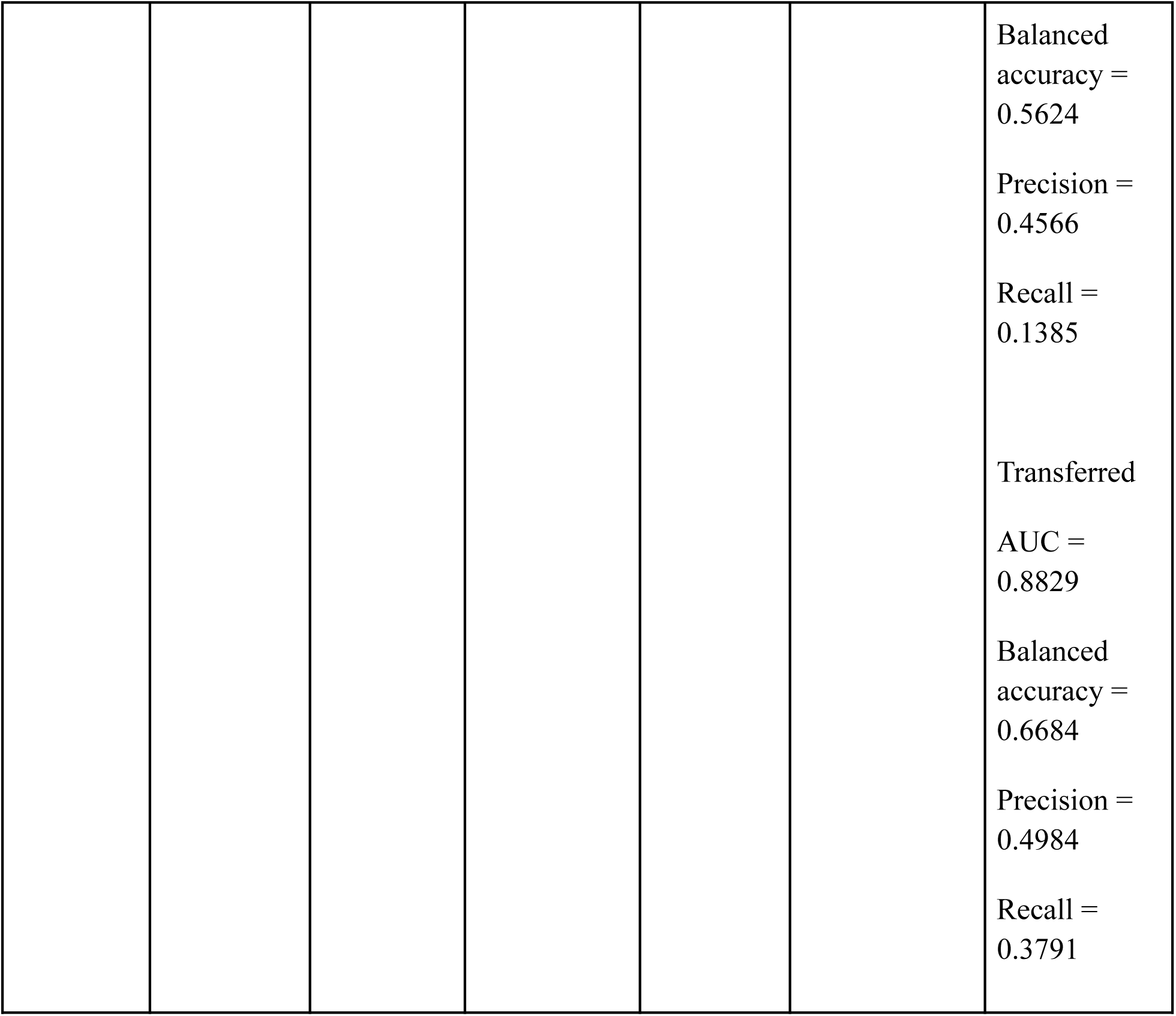
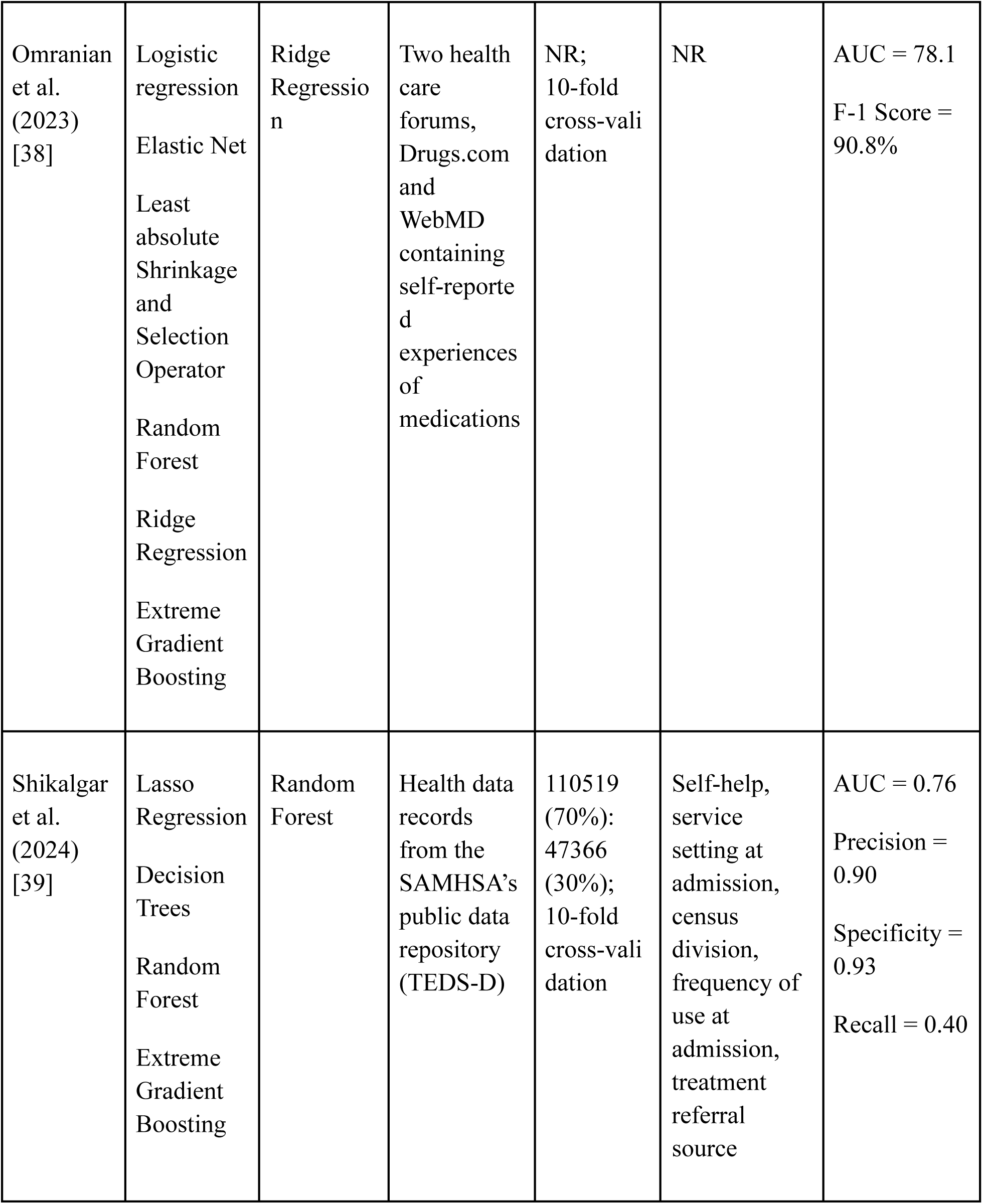
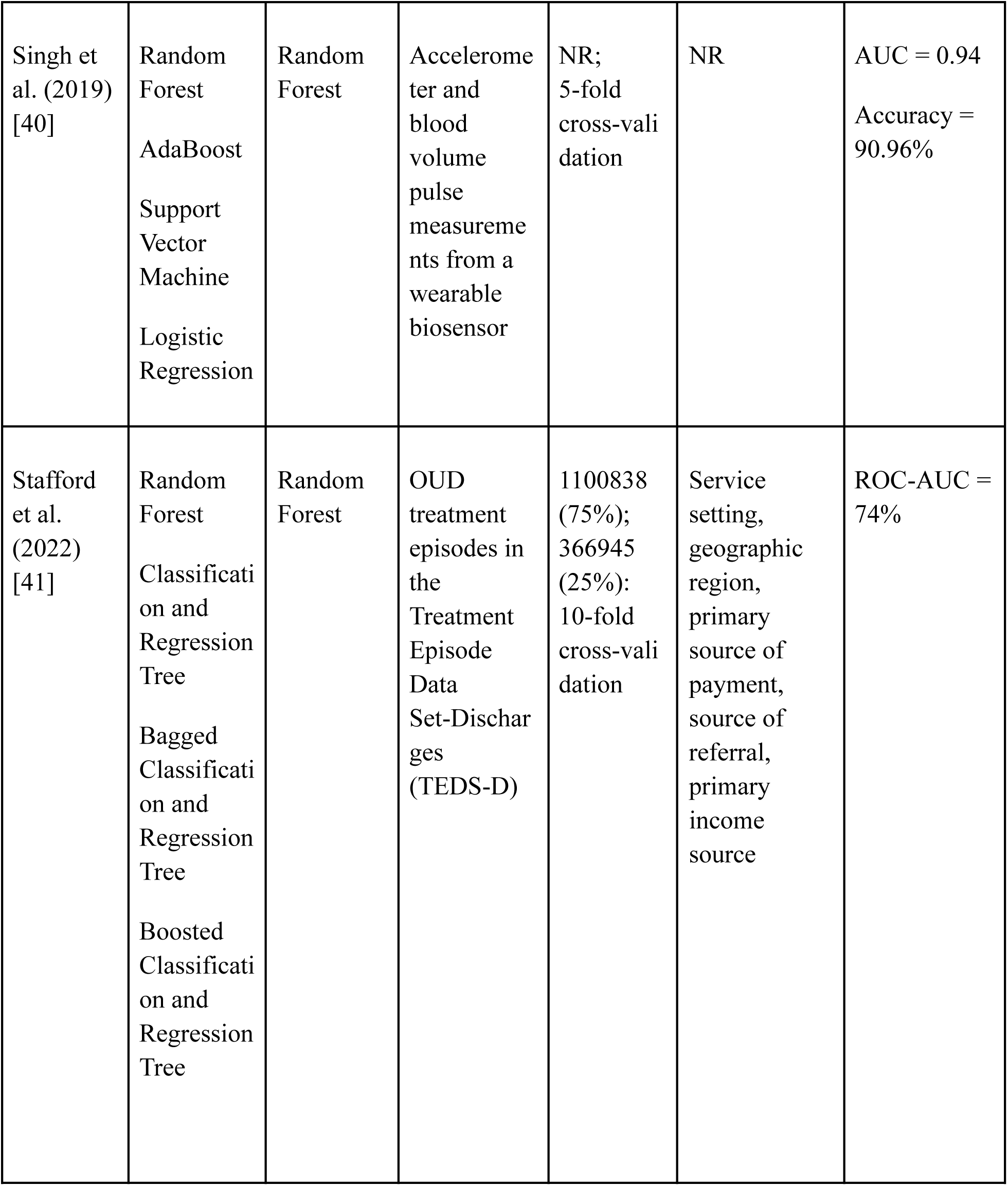
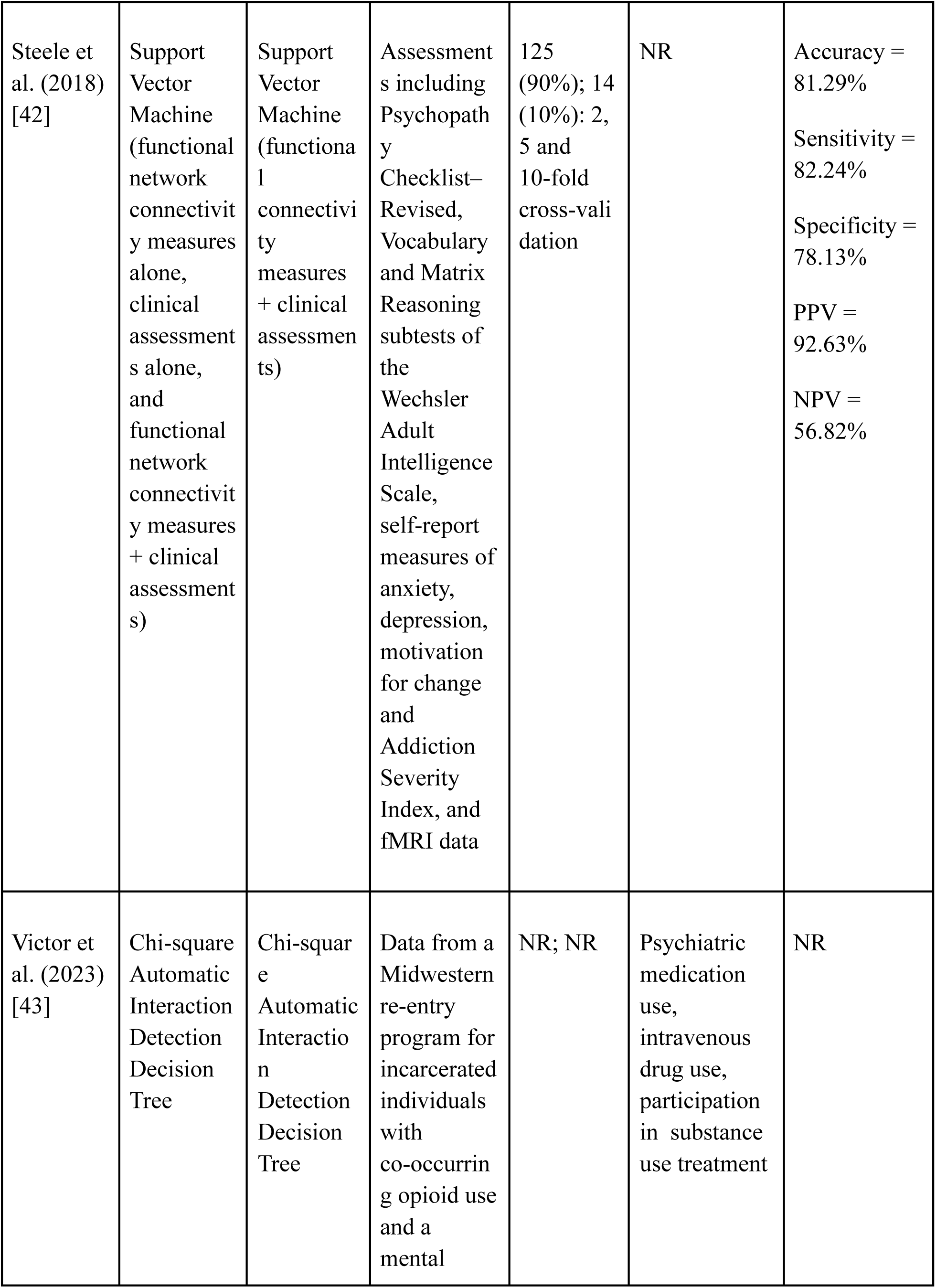
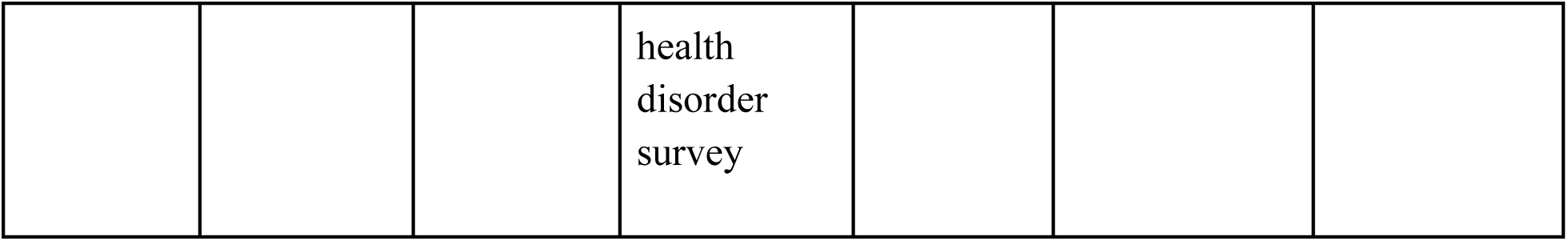
Summary of AI/ML models utilized.

### Synthesis of findings by year

We identified an increase in the number of studies assessing the application of ML and AI models to predict treatment adherence among individuals with OUD over time (Table 3). The smallest number of studies that matched our search criteria were published from 2018 to 2020, each of which had only one (4.5%) relevant publication [29, 41, 43]. Three (13.7%) studies were published in 2021 [27, 33, 38], another three (13.7%) in 2022 [25, 31, 42], and four (18.2%) in 2023 [26, 28, 39, 44]. A majority of the included articles, amounting to nine (40.9%) studies, were published in 2024 [23, 24, 30, 32, 34–37, 40].

**Table 3.**
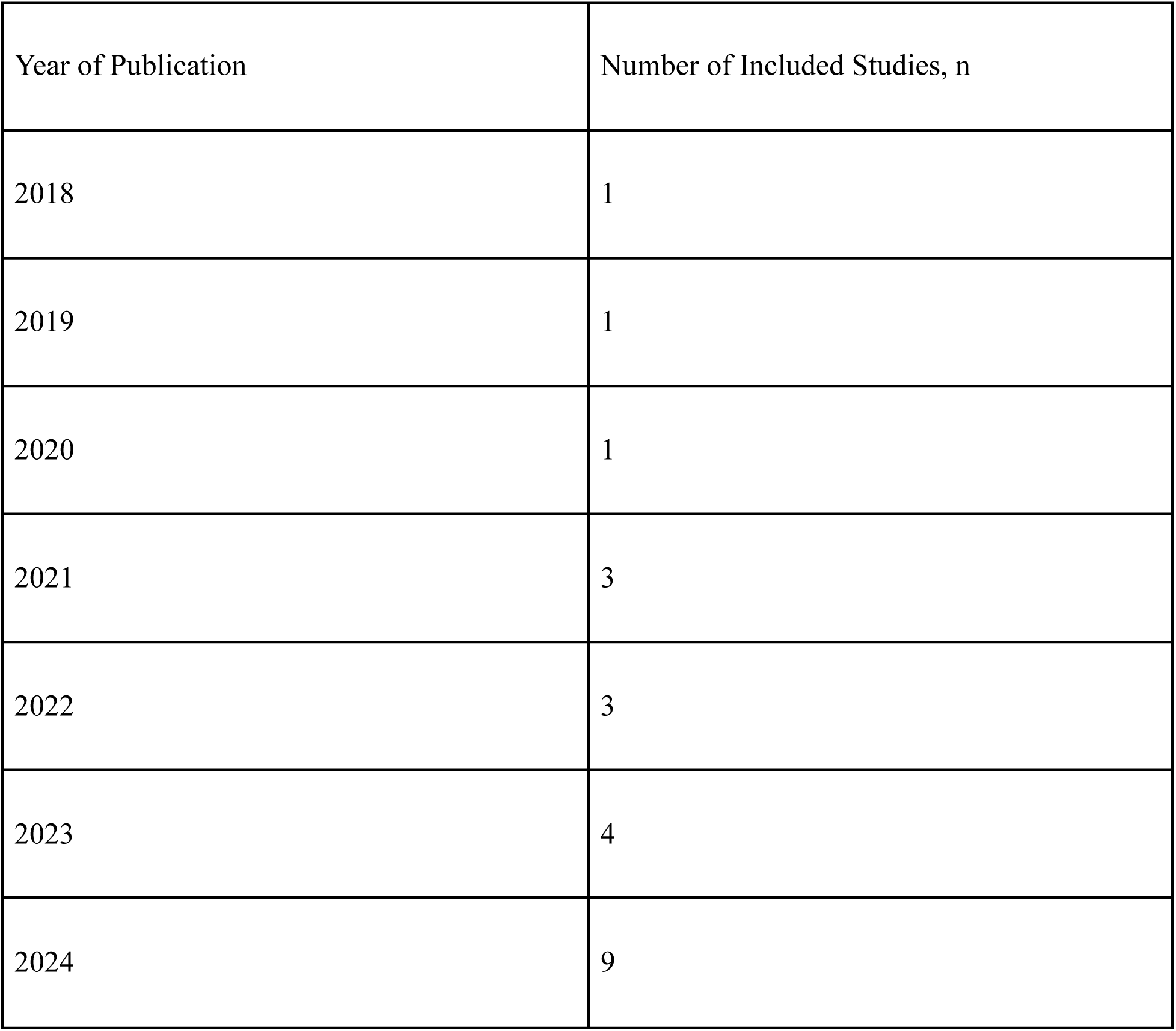
Number of included studies per year.

### Synthesis of findings by country

An analysis of included articles by country showed that most studies were conducted in the United States (Table 4). Numerically, 20 (91%) out of the 22 studies in this scoping review were US-based [23–26, 28–35, 37–44]. Only two of the included studies were based outside of the United States. Of these remaining studies, one (4.5%) was conducted in Australia [27] and one (4.5%) in Taiwan [36].

**Table 4.** Number of included studies per country.

| County of Publication | Number of Included Studies, n |
| --- | --- |
| Australia | 1 |
| Taiwan | 1 |
| USA | 20 |

### Synthesis of findings by adherence outcomes

Induction in OUD treatment [23], opioid cessation [29], relapse [37], and patient satisfaction with OUD treatment [39] were the least frequently explored outcomes and were assessed in only one (4.5%) study each. These were followed by non-adherence with treatment, which was the examined outcome in two (9.1%) articles [38, 41]. Three (13.7%) studies used AI and ML algorithms to predict treatment engagement and adherence [28, 30, 44], while retention in OUD treatment programs constituted the outcome of predictive models in four (18.2%) articles [26, 32, 34, 36]. The second most frequently examined outcome was treatment completion. This was assessed in six (27.3%) of the 22 studies in this scoping review [25, 26, 35, 38, 40, 43]. Treatment discontinuation, cessation, or drop out was explored in seven (31.8%) articles, making it the most frequently measured outcome in literature on the application of AI and ML algorithms to predict adherence to treatment and management among patients with OUD [24, 27, 31, 33, 36, 38, 42]. The number of included studies by outcome are presented in Table 5.

**Table 5.**
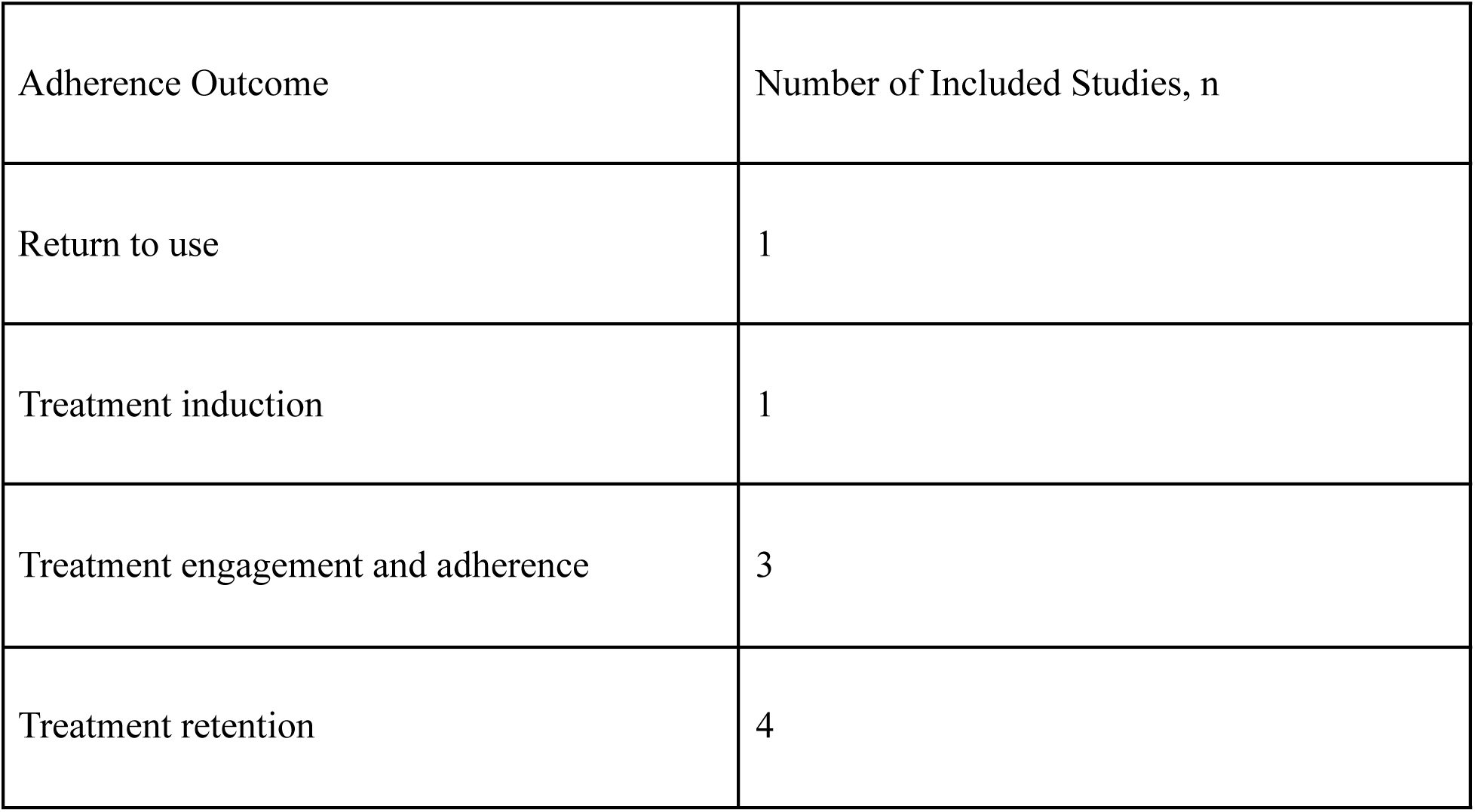
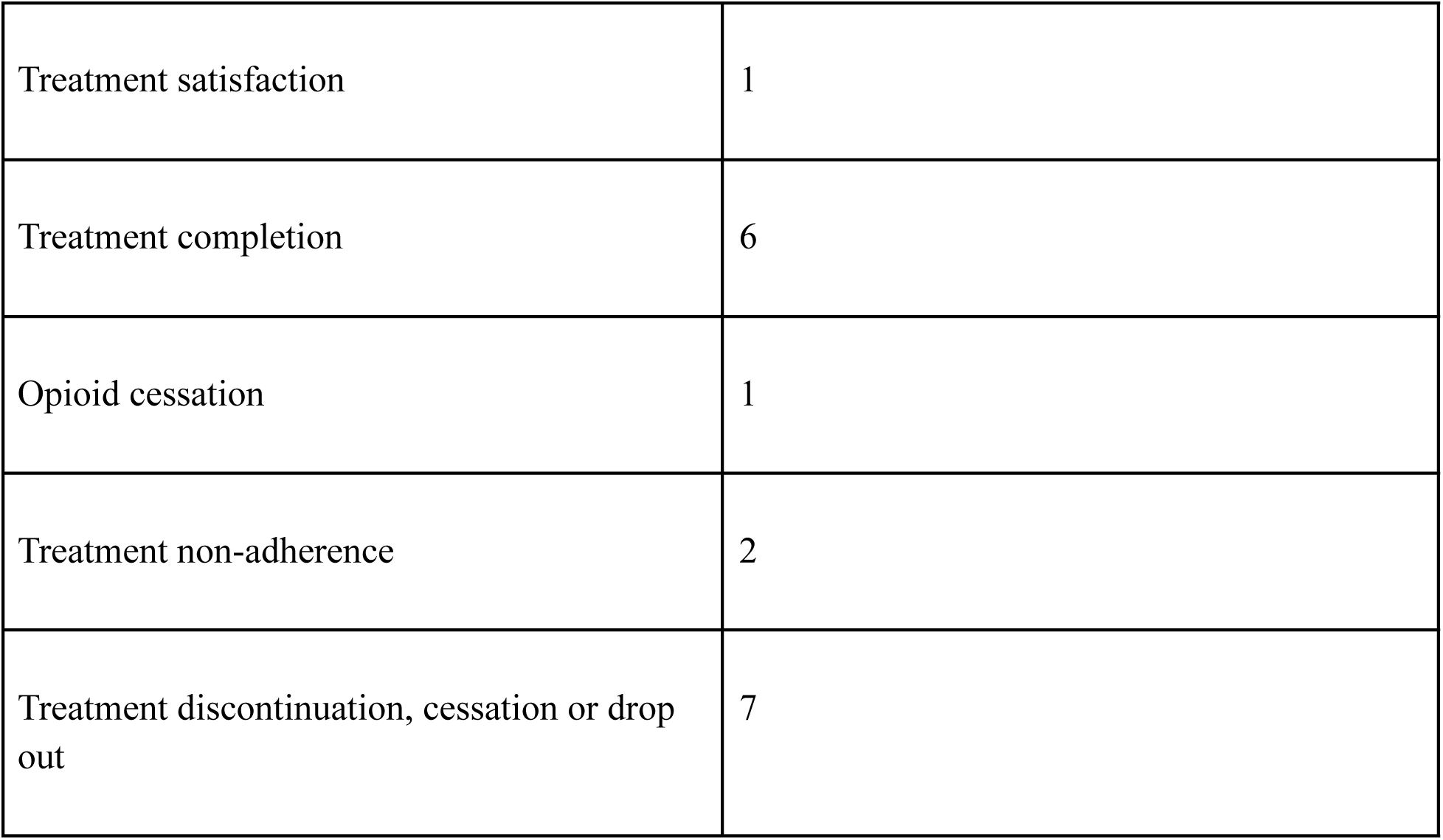
Number of included studies per treatment adherence outcome.

### Synthesis of findings by best reported AI and ML models

Random forest models were identified as the best performing AI and ML algorithms for predicting treatment adherence outcomes among individuals with OUD in ten (45.5%) studies, constituting a majority of the included articles [24–26, 30–33, 40–42]. Chi-square automatic interaction detection decision tree [35, 44], extreme gradient boosting [28, 38], and support vector machine [29, 43] were deemed the most effective predictive models in two (9.1%) studies each. Of the remaining six articles, one (4.5%) study each identified the following as top performing AL and ML models: deep learning [27], gradient boosting machine [34], logistic regression [37], neural network [36], ridge regression [39], and Tian-EN [23]. A list of AI and ML models and the corresponding number of studies that report them as having the highest predictive power is displayed in Table 6.

**Table 6.** Number of included studies per best reported model.

| Best Reported Model | Number of Included Studies, n |
| --- | --- |
| Chi-square Automatic Interaction Detection<br>Decision Tree | 2 |
| Deep Learning | 1 |
| Extreme Gradient Boosting | 2 |
| Gradient Boosting Machine | 1 |
| Logistic Regression | 1 |
| Neural Network | 1 |
| Random Forest | 10 |
| Ridge Regression | 1 |
| Support Vector Machine | 2 |
| Tian-EN | 1 |

## Discussion

This scoping review was conducted to provide an overview of the state of evidence on the use of AI and ML models to predict outcomes related to treatment adherence among patients with OUD. The findings from each of the 22 included articles were synthesized individually and reported along with information on the studied AI and ML algorithms, their predictive performance, key modelling variables, and the adherence outcomes measured. Moreover, studies were collectively examined to identify patterns in the context of publication dates, geographic distribution, treatment adherence outcomes, and top AI and ML models.

The included studies were all published between 2018 and 2024 [23–44], indicating that predicting adherence to treatment among patients with OUD using AI and ML algorithms is a fairly recent topic within the research literature. Relevant articles were found to be low in number during the first three years [29, 41, 44]. The trend changed in 2021 with an increasing number of studies being published each year [23–28, 30–40, 42, 44], potentially representing a rising interest in the topic.

Geographically, evidence on the application of AI and ML tools to forecast treatment adherence in an OUD patient population comes from three countries: Australia, Taiwan, and the United States, all of which are considered developed economies [23–44]. Only one (4.5%) of the included studies was based in Taiwan (i.e., the east) [36], while an overwhelming majority (21 studies, 95.4%) of the relevant research was concentrated in western countries [23–35, 37–44]. This illustrates a lack of representation of both developing and eastern countries in pertinent literature, in turn limiting the generalizability of findings to these regions. Moreover, a major proportion (20 articles, 91%) of the studies in this scoping review were conducted in the United States [23–26, 28–35, 37–44]. It is possible that the findings in these articles regarding top models and predictors do not extend to a non-American context, impacting the generalizability of these studies and ultimately the results of this scoping review.

Although random forest models took an obvious lead in the proportion of articles identifying them as the best algorithm [24–26, 30–33, 40–42], there was substantial heterogeneity across studies regarding which AI or ML technique was most suitable for the analyses. However, candidate AI models and applications assessed in the literature were similar. Random forest algorithms were an example of one such model and were evaluated in 14 (63.6%) articles [24–26, 29–34, 38–42]. Another example included models employing logistic regression which were examined in 11 (50%) studies [24–26, 28, 31–34, 37, 39, 41]. Similarly, a comparative analysis of the predictive ability of extreme gradient boosting was performed as a part of eight (36.4%) studies [24, 25, 28, 32, 33, 38–40]. While many algorithms were tested in several of the included publications, some candidate models appeared much less frequently. For example, the performance of artificial neural networks [33, 38] and chi-square automatic interaction detection decision trees [35, 44] were investigated in only two (9.1%) studies. Similarly, models like Gentle Boost and LogitBoost were explored in a single (4.5%) study. These observations point towards a need for additional evidence, preferably with a more standardized set of candidate models, to provide deeper insight into which models excel in their ability to forecast treatment adherence in individuals with OUD relative to other algorithms.

Heterogeneity in the data was also evident in the evaluation metrics used to gauge the performance of AI and ML models. Performance was assessed across a major (nine studies, 40.9%) proportion of the included articles with AUC [24–26, 30, 35, 38–41]. Accuracy was utilized as a metric in seven (31.8%) studies [25, 26, 28, 29, 30, 41, 43]. Precision [25, 32, 33, 38, 40], recall [25, 32, 33, 38, 40], and specificity [28, 30, 31, 40, 43] were employed as performance metrics in five (22.7%) publications. This was followed by AUROC [28, 34, 37, 42], F-1 score [25, 28, 33, 39], and sensitivity [28, 30, 31, 43] were calculated in four (18.2%) of the 22 articles. Other measures of evaluation included c-statistics [24, 27, 33], positive and negative predictive values [28, 43], qini-value [23], c-for-benefit [23], d-statistics [27], r-squared value [27], integrated Brier score [27], AUPR [28], mean correlation coefficient [36], and balanced accuracy [38]. The heterogeneity seen in performance metrics limits cross-study comparisons, particularly for measures that are used in few studies, making it difficult to draw meaningful conclusions.

Several of the five most important modelling features were common to at least two studies. This included age, employment levels, geographic location, length of stay, service setting, source of referral, and involvement with the criminal justice system [24–27, 29, 30, 35, 38, 40, 42]. This overlap among modelling variables suggests that AI and ML models from these studies might potentially be applicable to different samples or populations of patients while retaining their predictive performance. However, substantial research is required into the external validity of the AI and ML models before their external validity can be conclusively established. It is also worth noting that top predictive variables were not reported in seven (31.8%) of the 22 articles [32–34, 36, 39, 41, 43], hindering attempts at testing for replicability by researchers outside of the original group of authors.

Moreover, the findings of this scoping review must be interpreted in light of some additional limitations. First, there was a significant degree of variation in the outcomes used to assess treatment adherence. Second, the adherence-related outcomes in the included studies are measured over a duration of one year or less. As such, the best reported algorithms identified in this review may not be as effective in predicting longer-term outcomes, potentially affecting the generalizability of the results. Third, some studies tested their AI and ML models on a small sample. The results from these studies had low statistical power. Fourth, despite the comprehensiveness of the search strategy, there remains a possibility of the exclusion of potentially eligible studies that may not have been indexed in the selected databases. Finally, there is a possibility that factors relevant for AI and ML algorithms might evolve over time, resulting in changes to the optimal set of predictive variables and models considered most suitable for predicting adherence outcomes in an OUD patient population. This implies that the scoping review may need to be updated in a few years to ensure that the findings are reflective of changes in the relevant literature.

## Conclusion

OUD is a significant public health and societal concern, often associated with poor treatment adherence. This scoping review synthesized literature on leveraging AI and ML to predict treatment adherence and the lack thereof in patients with OUD. An interest surrounding the topic was on the rise since 2018 with most of the research coming from the United States and random forest algorithms taking the lead as the model of choice in terms of predictive performance. Moreover, there was considerable heterogeneity in the top models, performance metrics, and modelling variables, highlighting the need for more standardized models, outcomes, and predictive variables in future research to facilitate cross-study comparisons.

## Author Contributions

Aghna Wasim, Ali Abud, and Samir Malick: investigation, methodology, conceptualization, screening, data extraction, writing (original draft), and writing (review & editing); Nazeefa Arifina Nashrah, Veronika Lošanová, Hetvi Raimugia, Marisha Karim, Minh Van Khanh Le, Venusha Baskarathasan, Charlotte Gibson, Siba Alkhatib: screening, data extraction, writing (review & editing); Silvia S. Martins: critical revision of the manuscript, writing (review & editing). All authors have read and approved the manuscript for submission.

## Data Availability

All data utilized in this study are available in the published article or the supplementary material.

## Acknowledgements

None.

## Declaration of Interests

The authors declare that they have no competing interests.

## Funding

None.

## Supporting Information

Supplementary Material 1: Complete List of Predictive or Modeling Variables for Each Study and Search Strategy

Supplementary Material 2: PRISMA-ScR Checklist

---

**S1.** Complete List of Predictive or Modeling Variables for Each Study.

S1. Complete List of Predictive or Modeling Variables for Each Study
| Author (Year of Publication) | Predictive/Modeling Variables |
| --- | --- |
| Afshar et al. (2024) | Age at randomization, gender, ethnicity, education, looking for work, late randomization, high severity addiction, heroin as the major drug of abuse, any heroin use, heroin use route, cannabis use (30 days), cocaine use (30 days), opiate use onset, opioid withdrawal score, nicotine use onset, primary drug cost per day, living with an alcoholic, living with a drug user, homeless, on parole, smoking, chronic pain for 6 months, moderate or extreme pain, psychiatric diagnosis, moderate or extreme anxiety/depression, Hamilton depression score, alcohol use disorder, amphetamine use disorder, benzodiazepine use disorder, cannabis use disorder, cocaine use disorder previous methadone/ buprenorphine treatment success, strong medication preference, no medication preference, preference for SUB-BUP, no preference for SUB-BUP, no medication preference, preference for ER-NTX, no preference for ER-NTX |
| Al Faysal et al. (2024) | Age, sex, geographic region, residence, insurance plan type, relationship to policyholder, geographic location, chronic pain condition, HIV/AIDS condition, Hepatitis C condition, depressive disorder, anxiety, post-traumatic stress disorder, bipolar disorder, schizophrenia, non-opioid drug use disorder, alcohol use disorder, opioid use disorder severity level, cannabis use disorder severity level, Charlson Comorbidity Index, number of days in first buprenorphine subscription, use of antidepressants, use of antipsychotics, use of mood stabilizers, use of benzodiazepines, use of non-benzo, use of stimulants, use of opioid analgesics, number of inpatient visits, number of emergency department visits, number of outpatient visits, proportion of days covered measure 1-month of treatment initiation, proportion of days covered measure 3-month of treatment initiation |
| Baird et al. (2022) | Age, DSM diagnosis, length of stay, primary substance used, secondary substance used, referral source, frequency of use of primary substance at admission, frequency of use of primary substance at discharge, employment status at discharge, type of service the patient is discharged from |
| Baird et al. (2023) | Age, highest grade completed, number of self help groups attended 30 days prior to treatment, number of self help groups attended during treatment, number of goals set by client, referral source, drug of choice, secondary drug of choice, number of previous treatment episodes, linked to supplemental nutrition assistance program after intake |
| Bharat et al. (2021) | Age at treatment initiation, gender, Aboriginal and/or Torres Strait Islander, remoteness, incarcerated prior to opioid agonist treatment entry, number of convictions prior to opioid agonist treatment entry, medication type, year of first opioid agonist treatment entry, dosing point, tenure of prescriber's opioid agonist prescribing, prescriber working in multiple locations, history of noncompliance with medical treatment and regimen, outpatient treatment for DUDs, schizophrenia, homelessness, prior treatment for harmful opioid use, tobacco dependence, alcohol use disorder, occupant of vehicle in an accident, mixed anxiety and depressive disorder, abdominal pain, other and unspecified abdominal pain, treatment for opioid dependence, treatment for opioid withdrawal, personal history of tobacco use disorder, current tobacco use, 4-aminophenol derivatives |
| Burgess-Hull et al. (2023) | BlueCross as primary insurance type, Medicaid as primary insurance type, Medicare as primary insurance type, possible toxicology adulteration, urine positive for fentanyl, urine positive for non-fentanyl opioid, urine positive for cocaine, urine positive for benzodiazepine, urine positive for norbuprenorphine, urine positive for amphetamine, cumulative sum (cocaine, benzodiazepine, amphetamine, gabapentin), treatment adherence rate, attendance rate, treatment adherence rate x number of appointments attended (interaction), treatment adherent appointments, number of appointments attended, lagged urine positive for fentanyl, lagged urine positive for non-fentanyl opioids, lagged urine positive for norbuprenorphine, lagged urine positive for cocaine, lagged urine positive for norbuprenorphine, treatment adherence at current appointment, treatment adherence at last appointment |
| Cox et al.<br>(2020) | **For African Americans**<br>Current age, sex, time since first opioid treatment, older age at first opioid treatment, number of years using heroin, depressed after reducing cocaine use, time since last injected cocaine, time since last used cocaine, time since last stayed high in cocaine, used cocaine <11 times within year of interview, treated in outpatient program for cocaine use, time since first cocaine craving, never injected cocaine, often used marijuana more than intended to, mixed alcohol and drugs > 3 times in 12 months, time since last had alcohol symptoms lasting >1 month, smoked less frequently after waking up, older age at first cigarette, 2 marijuana symptoms lasting a month, HIV positive, previous health always better than current health<br>**For Individuals with European Ancestry**<br>Current age, household income, body mass index, time since last cocaine use, older age at first heavy opioid use number of years using heroin, time since last cocaine injection, time since last had alcohol symptoms that last >1 month, >20 outpatient visits for drug/psychiatric problems in the last year, time since opioid treatment initiation, used cocaine >11 times in last year, time since first used opioid 1/week for >1 month, have injected cocaine, older age at first heavy cocaine use, marijuana interfered with work/home, time since starting opioid self-help group, used tobacco but not addicted, disclosed problems with cocaine usage to professional, stopped using stimulants for >3 months, drinking resulting in objections or problems with family and work, <3 ASP criteria in 12-month period, often failed to pay debts, suspended or expelled from school, time since last suicidal ideation, age last had antisocial behaviour, no fear or most disturbing/traumatizing event, avoided scene that reminded of traumatic event, OCD behaviours when depressed, feeling distracted, unsafely raced cars, depression always started with drug problems, number of depression symptoms, outstanding emotional problem |
| Eddie et al.<br>(2024) | Age, sex, race/ethnicity, employment status, substance use severity, prior substance abuse treatment, treatment motivation index, treatment resistance index, victimization, substance use age of onset >15, current withdrawal symptoms, dual diagnosis, depression symptoms, emotional problem scale, ADHD, conduct disorder, post-traumatic stress disorder, suicide problems, criminal justice involvement, living risk index, social risk index, homelessness experience, lack of peers in recovery, general crime scale, self-efficacy scale, population density, public assistance, income inequality, health care, count of liquor places near treatment center |
| Gottlieb et al.<br>(2022) | Gender, quality of life improvement, have relapsed since joining the program, have used prescription oxycodone, past legal problems, substance use history of benzodiazepines, veteran status, first time taking medications for opioid use disorder, housing status, prior use of barbiturates, substance use history of hallucinogens, substance use history of non-prescription opiates, substance use history of cocaine, prior use of non-prescription suboxone, substance use history of alcohol, substance use history of marijuana, substance use history of heroin, taken medications for opioid use disorder from inpatient services, prior use of cocaine, prior use of nicotine or tobacco, prior arrest for drug charges, religiosity, prior arrest for parole or probation violations, ever taken medications for opioid use disorder in outpatient services |
| Haredasht et al.<br>(2024) | Diagnosis of opioid dependence, diagnosis of limb swelling, polyethylene glycol-based laxative, oxycodone extended release, acetaminophen, age at BUP-NAL initiation, 12-lead ECG, diagnosis of opioid withdrawal, diphenhydramine, docusate, aspirin, lidocaine low dose buprenorphine, IV fluids, IV thiamine |
| Hasan et al.<br>(2021) | Age, income, gender, patient location (urban/rural), insurance type, provider speciality, addiction treatment specialist, pain specialist, early treatment adherence, PDC in first 2 months, PDC in first 3 months |
| Hayes et al.<br>(2024) | Age, gender, length of time for Veterans Health Administration (VHA) enrollment, religion, race, ethnicity, marital status, VHA priority status, state of residence, US census division of residence, rural-urban commuting area code, miles from closest VHA facility, minutes of drive-time to the closest VHA facility, employment status, VHA disability status, VHA service-connected disability eligibility status, VHA pension eligibility status, smoking status, next of kin, days' supply of initial buprenorphine, dose of initial buprenorphine, days covered of initial buprenorphine, inpatient or outpatient initiation of buprenorphine, days between OUD diagnosis and buprenorphine initiation, year of buprenorphine initiation, month of buprenorphine initiation, methadone orders between OUD diagnosis and buprenorphine initiation, days' supply of methadone between OUD diagnosis and buprenorphine initiation, naltrexone orders between OUD diagnosis and buprenorphine initiation, days' supply of naltrexone between OUD diagnosis and buprenorphine initiation, number of previous buprenorphine episodes, number of previous failed buprenorphine episodes, days covered from previous buprenorphine episodes, total buprenorphine dose from previous buprenorphine episodes, average buprenorphine dose from previous buprenorphine episodes, time since last buprenorphine episode, binary indicator for whether the difference between buprenorphine initiation and start of data is 365 days, fractional value for number of days between buprenorphine initiation and fiscal year 2006, number of days between buprenorphine initiation and start of fiscal year 2006, number of emergency room visits, number of primary care/medicine visits, number of mental health visits, number of pain clinic visits, whether or not brief addiction monitor (BAM) or brief addiction monitor-revised (BAM-R) has been administered, number of BAM or BAM-R administrations, days between buprenorphine initiation and BAM administration, individual constructs within BAM measured over the past 30 days, cannabis use disorder, cocaine use disorder, tobacco use disorder, alcohol use disorder, non-opioid drug use disorder, endocarditis, Hepatitis B, Hepatitis C, HIV, post-traumatic stress disorder, opioid overdose, non-opioid overdose, any mental health disorder, depression, anxiety, bipolar disorder, psychosis, suicide or self-inflicted injury, elixhauser, comorbidity count, opioid prescription duration of action, opioid prescription schedule, average morphine milligram equivalents, total opioid days covered, number of medication fills received, number of unique VHA categories of medications received, number of naloxone prescription received, days covered of antidepressants, days covered of benzodiazepines, days covered of skeletal muscle relaxants, drugs covered of non-opioid analgesics, days covered of hypnotics, Social Vulnerability Index (SVI) composite score, SVI household composition & disability score, SVI housing type & transportation score, SVI socioeconomic score, homelessness, justice-involvement, provider speciality, provide creditals, VHA or non-VHA |
|  | prescriber, trainee, number of buprenorphine written by prescriber, number of methadone prescriptions written by prescriber, number of naltrexone prescriptions written by prescriber, number of patients prescriber wrote buprenorphine for, number of patients prescriber wrote methadone for, number of patients prescriber wrote naltrexone for, percent of patients retained on buprenorphine in the year by prescriber, whether veteran received buprenorphine from closest VHA facility, facility type, whether facility had an inpatient detoxification program, whether facility had a methadone clinic, whether facility had a substance abuse disorder residential treatment program, flag for VHA station |
| Hong et al. (2024) | Gender, race/ethnicity, types of substances used, opioid use, age of first use for the primary substance, Adult Needs and Strength Assessment strengths, total actionable items, rural/urban residence, living arrangement, employment, education, any justice system involvement |
| Liu et al. (2024) | HIV positivity, referral by the criminal justice system, 90-day urine opiate test results, last methadone dosage taken, willingness to receive treatment, 7-day treatment adherence, 30-day treatment adherence |
| Luo et al. (2024) | Heroin use days, morphine-positive urine drug screen, cocaine-positive urine drug screen, heroin injection in the past 30 days |
| Nasir et al. (2021) | Age, individual as the referral source, drug use care provider as referral source, health care provider as referral source, school as referral source, employer as referral source, community referral, education, primary substance frequency at admission, secondary substance frequency at admission, homeless, dependent living arrangements, primary substance frequency, length of stay, treatment locality population, treatment locality total crime cases, treatment locality disorderly conduct cases, change in self-help attendance, short-term rehab services, change in arrests, planned medication-assisted opioid therapy, treatment locality vagrancy cases, full-time employment status, substance use type (alcohol only), CBSA 35300, Services (short term rehab) CBSA 31700, CBSA 17200, CBSA 31700, CBSA 45860 |
| Omranian et al.<br>(2023) | Symptoms (suffocate, breathing problem, dehydration, vomit, confusion, restlessness, disoriented, muscle weakness, mood changes, depressing), drugs (hydroxyzine, luvox, temazepam, vitamins, miralax, fioricet, Narcan, magnesium citrate, Adderall, Ambien), illness (adrenal crisis, delirium, reflex sympathetic dystrophy, cyst, hydrocephalus, chronic headaches, sciatica, fibromyalgia, herniated discs, degenerative joint disease), duration of treatment, oral sensation, side effects, insurance, doctor visit |
| Shikalgar et al.<br>(2024) | Age at first use, age at admission,gender, ethnicity, education, employment status, marital status, service setting at admission, census division, year of discharge from substance use treatment, treatment referral source, frequency of drug use at admission, primary substance route of use, second substance use at admission, injecting substance as route of administration, no prior treatment episodes, cocaine use, heroin use, marijuana use, alcohol use, benzodiazepine use, methamphetamine use, hallucinogen use, arrests 30 days prior to discharge, comorbid psychological problems, homeless, veteran status |
| Singh et al.<br>(2019) | **Features taken from accelerometer**<br>Mean, medica, skewness, variance, standard deviation, mean crossing rate, maximum value, minimum value, mean derivatives, interquartile range, zero crossing rate, kurtosis<br>**Features taken from blood volume pulse**<br>Number of peaks, mean of peaks, standard deviation of peaks, root mean square, distance from maximum to minimum of peaks, mean peak-to-peak distance, standard deviation of peak-to-peak distance, power spectral density of signal |
| Stafford et al.<br>(2022) | Age, sex race, ethnicity, marital status, education, employment status, primary income source, service setting, geographic region, primary source of payment, referral source, planned use of medication for OUD, maximum frequency of use, days waited to enter treatment, prior treatment episodes, number of substances used, age of first substance use, infection use, heroin use, stimulant use, alcohol use, marijuana use, hallucinogen use, sedative use, inhalant use, tranquilizer use, arrests in 30 days prior to treatment, living situation, veteran status, diagnosed psychological problem |
| Steele et al.<br>(2018) | Age, sex, IQ, years of drug use, psychopathy, anxiety, depressive symptomatology, functional network connectivity between networks incorporating the anterior cingulate cortex; striatum; and insula |
| Victor et al.<br>2023 | Psychiatric medication use, intravenous drug use, participation in substance use treatment |

**S2.**
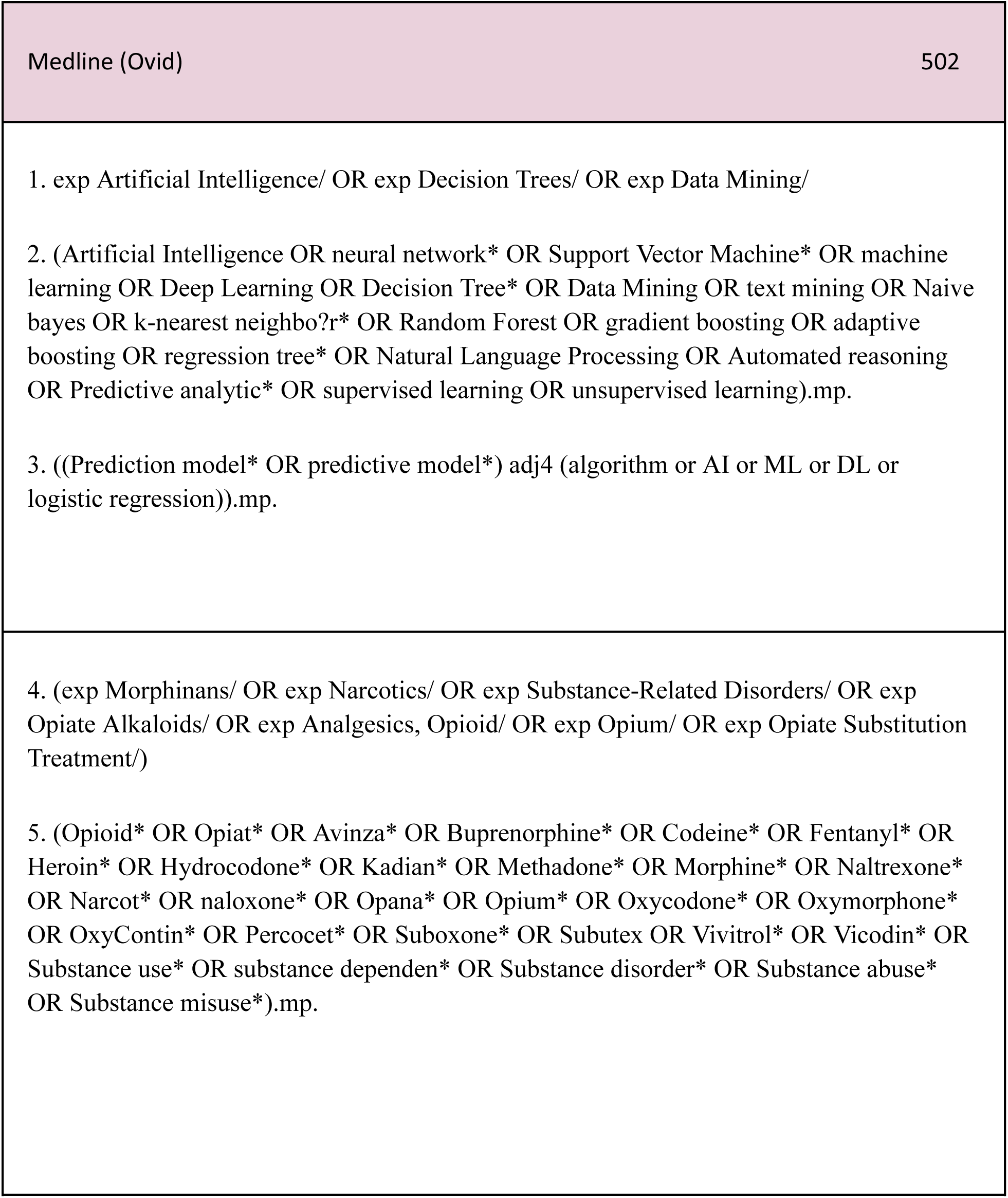
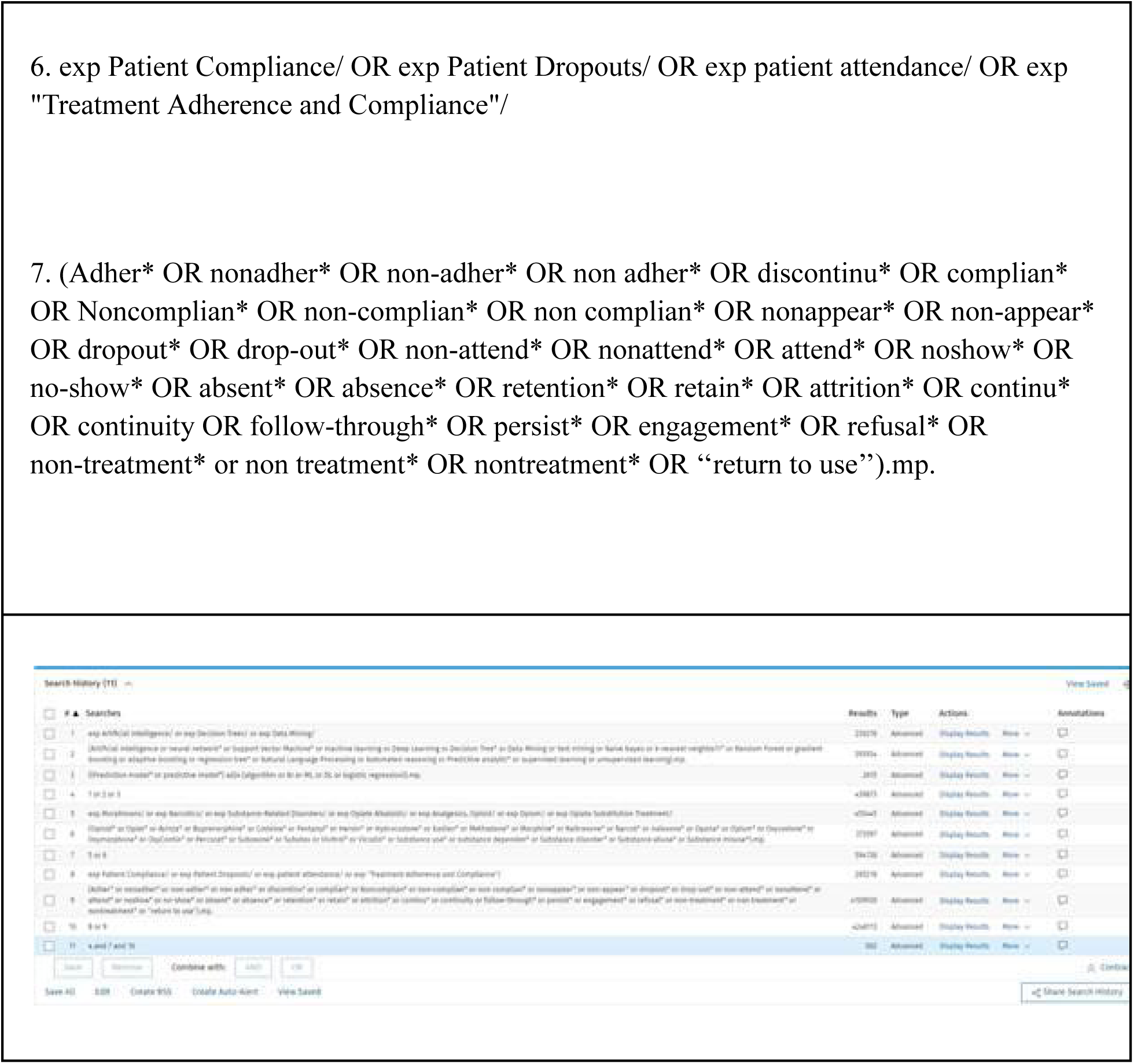
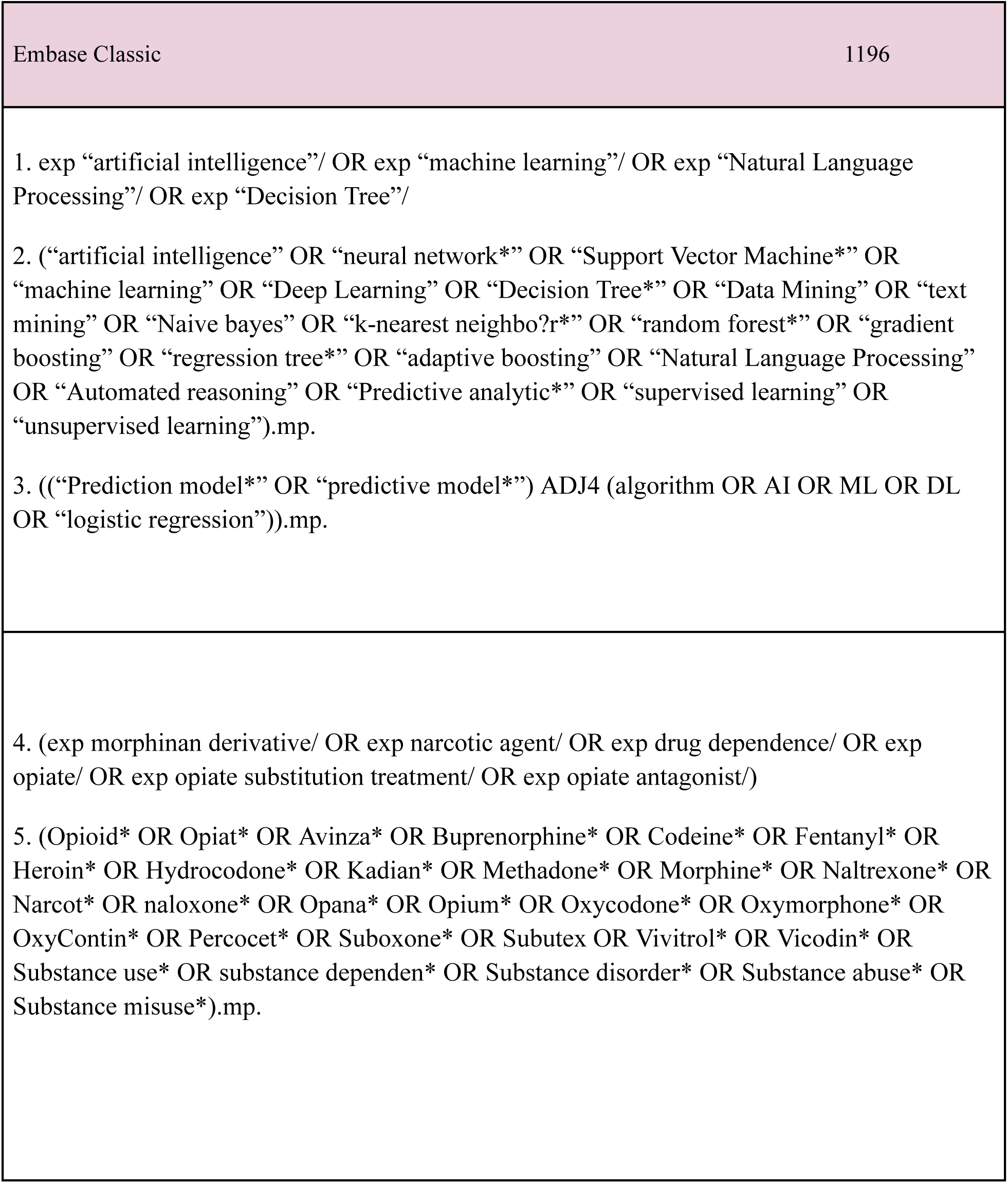
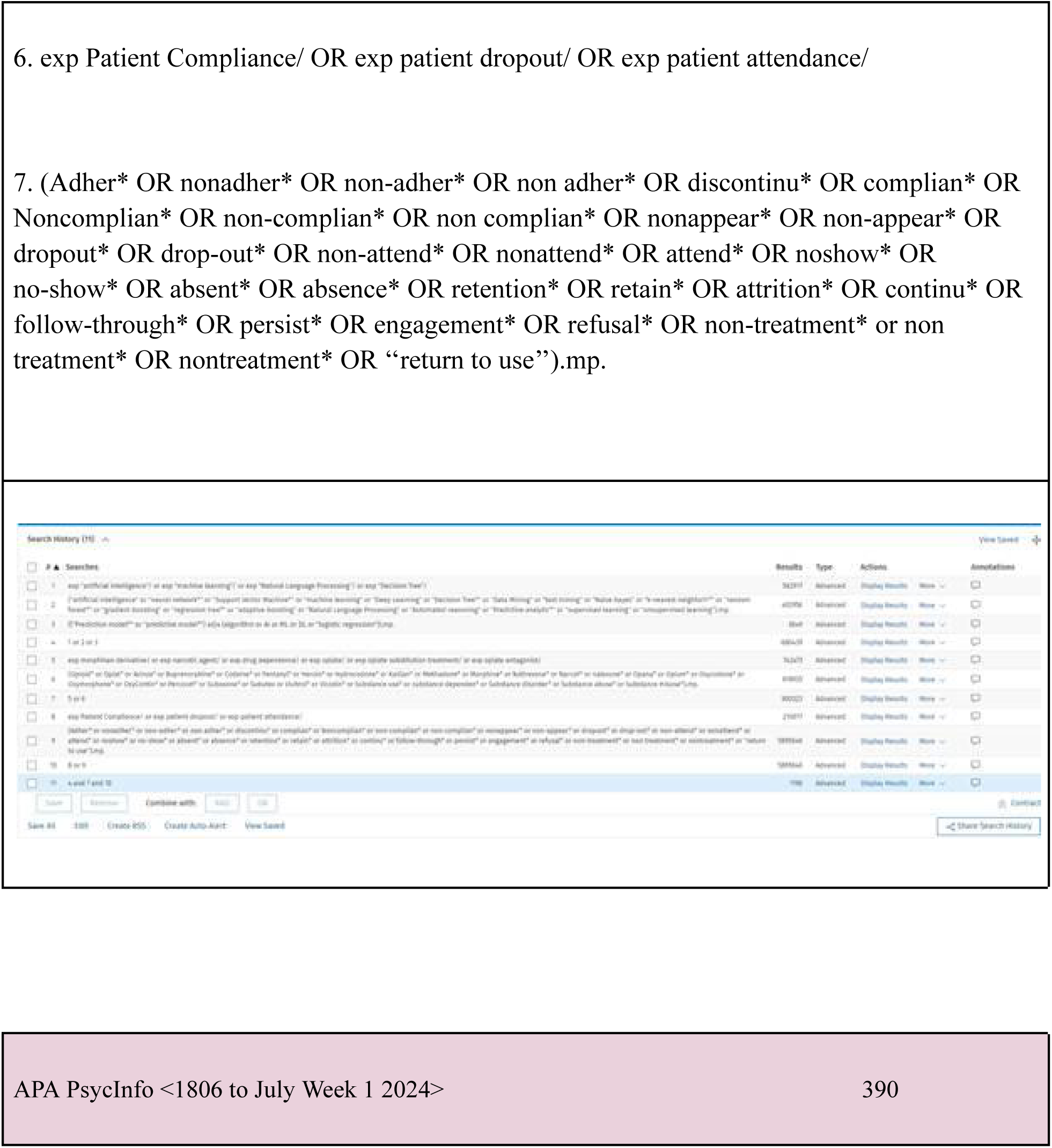
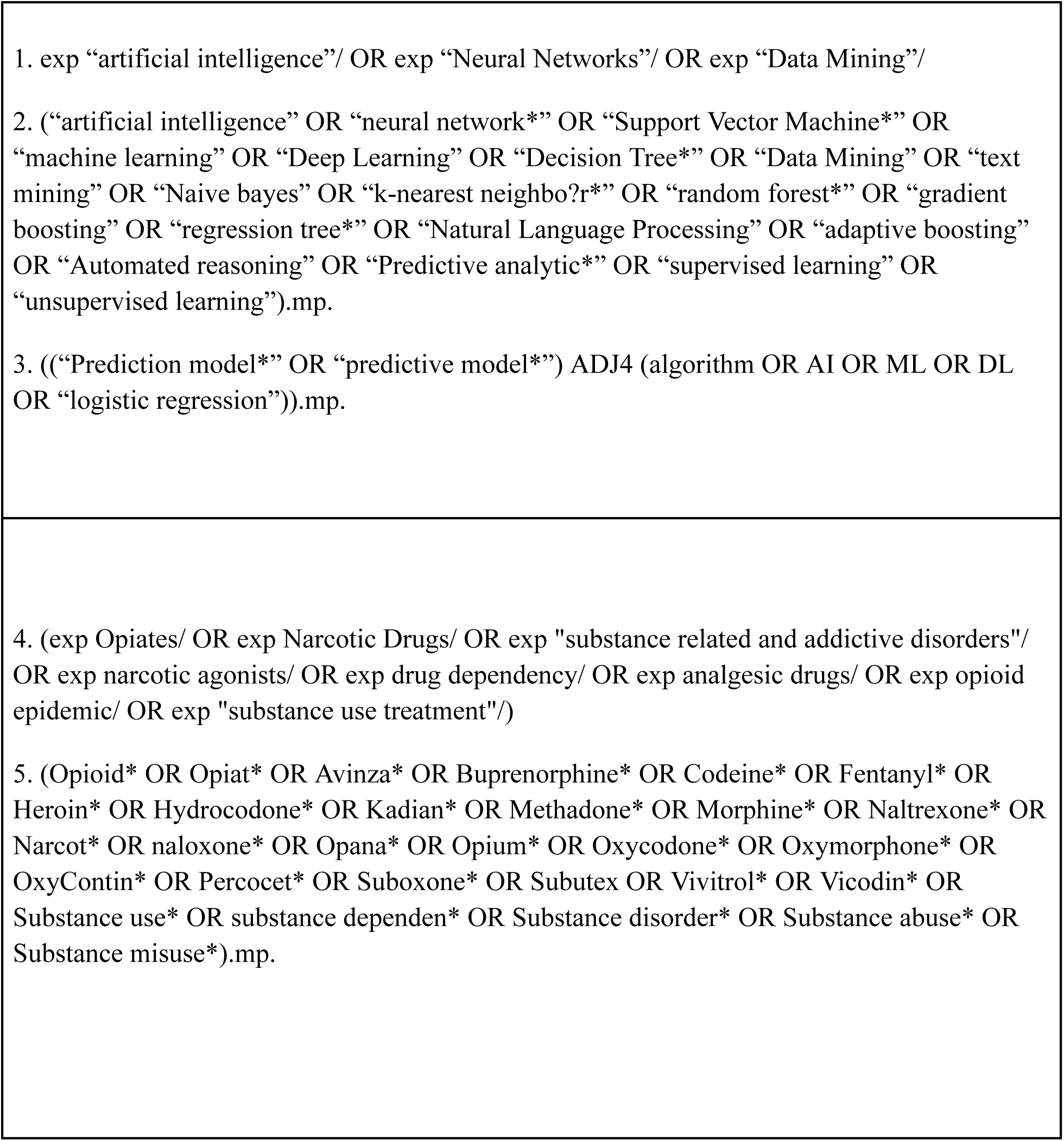
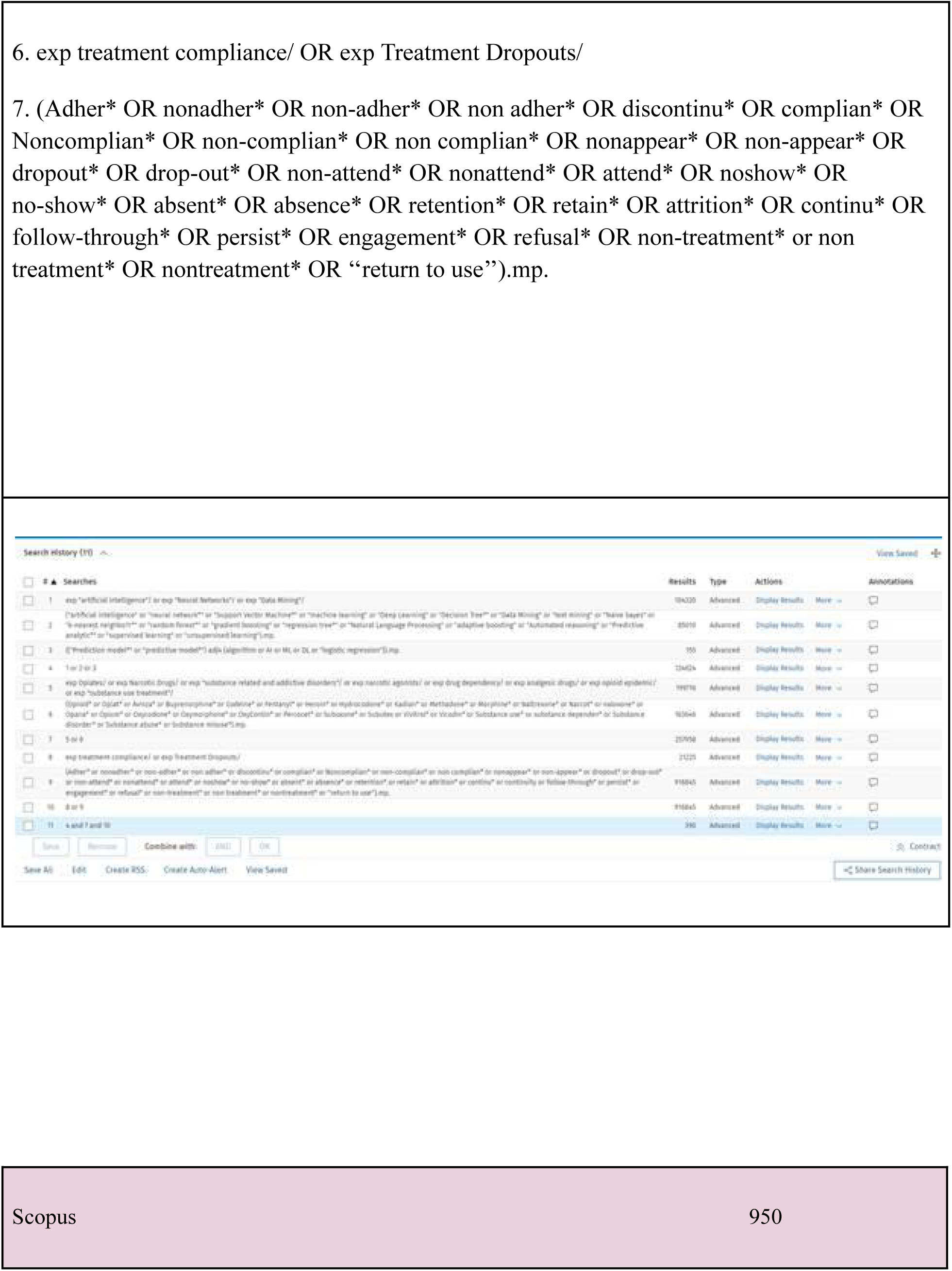
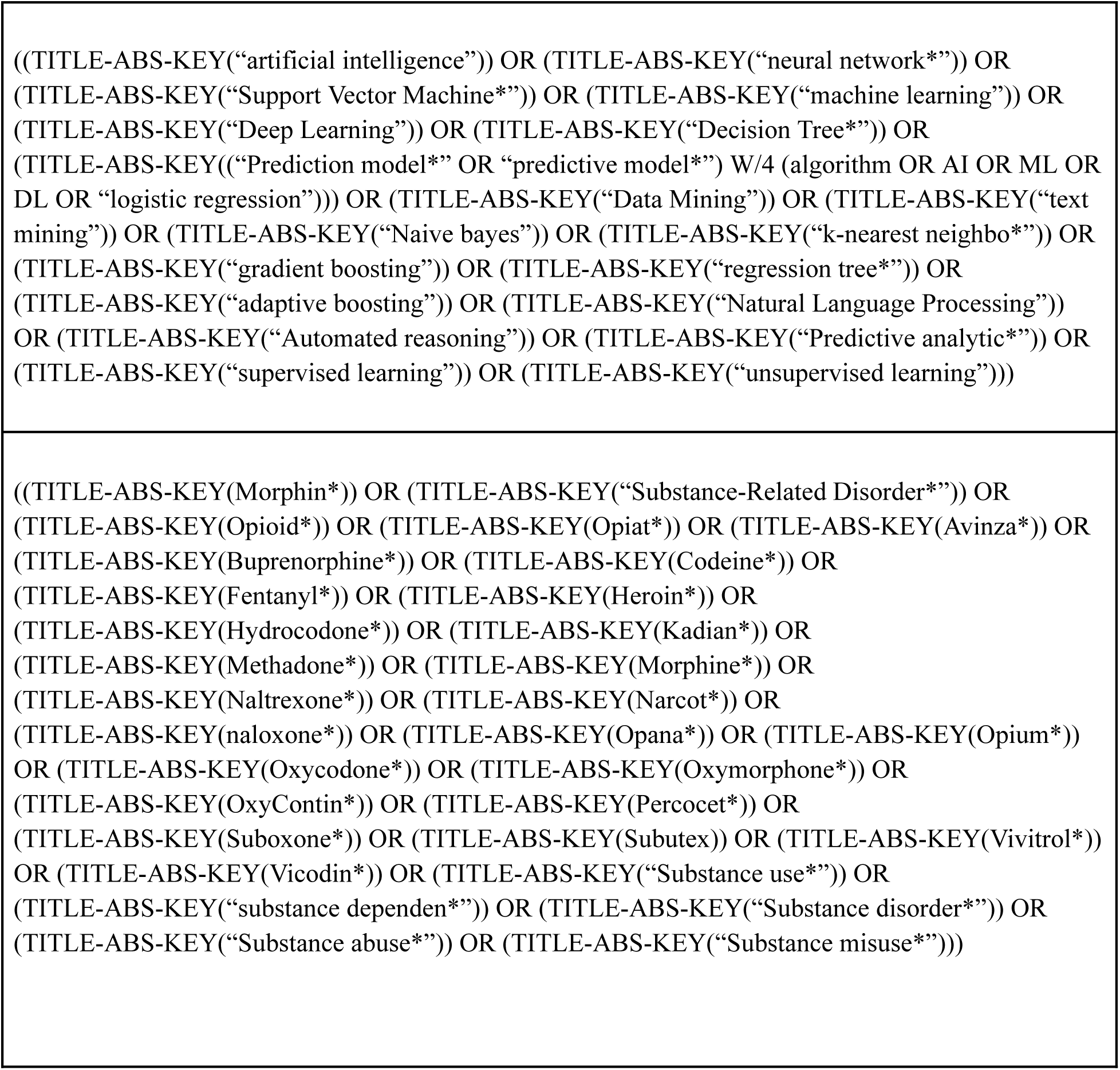
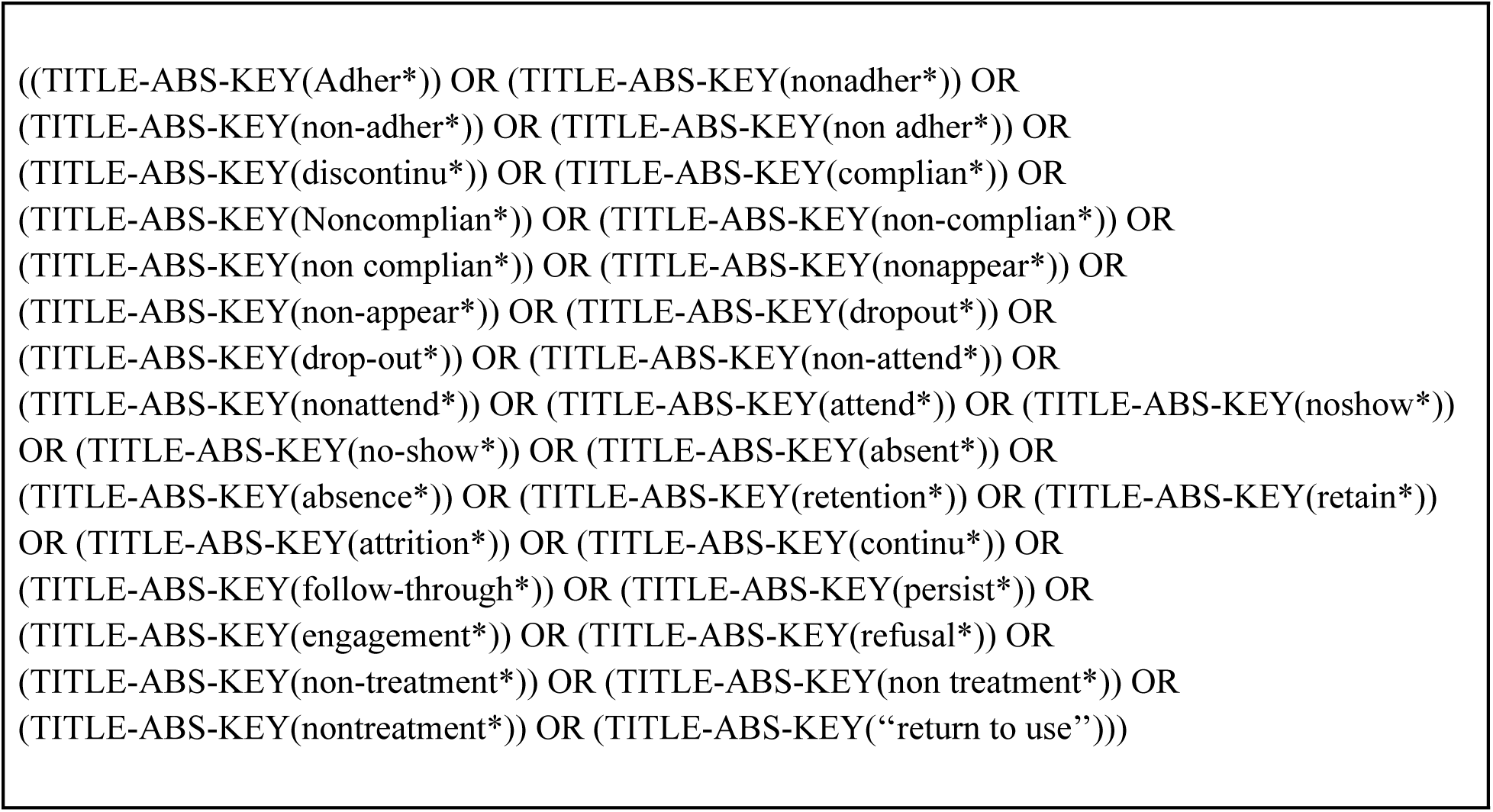
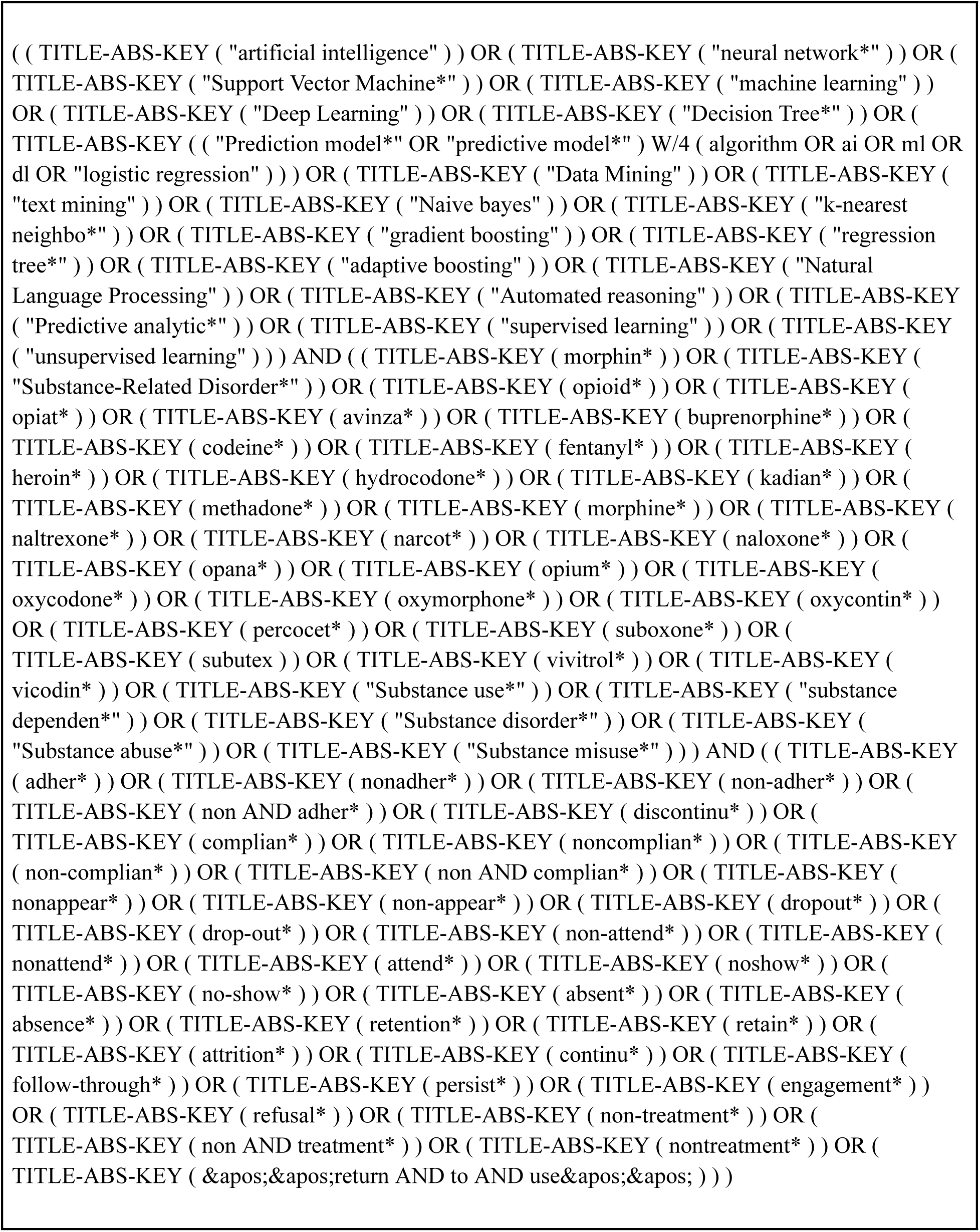
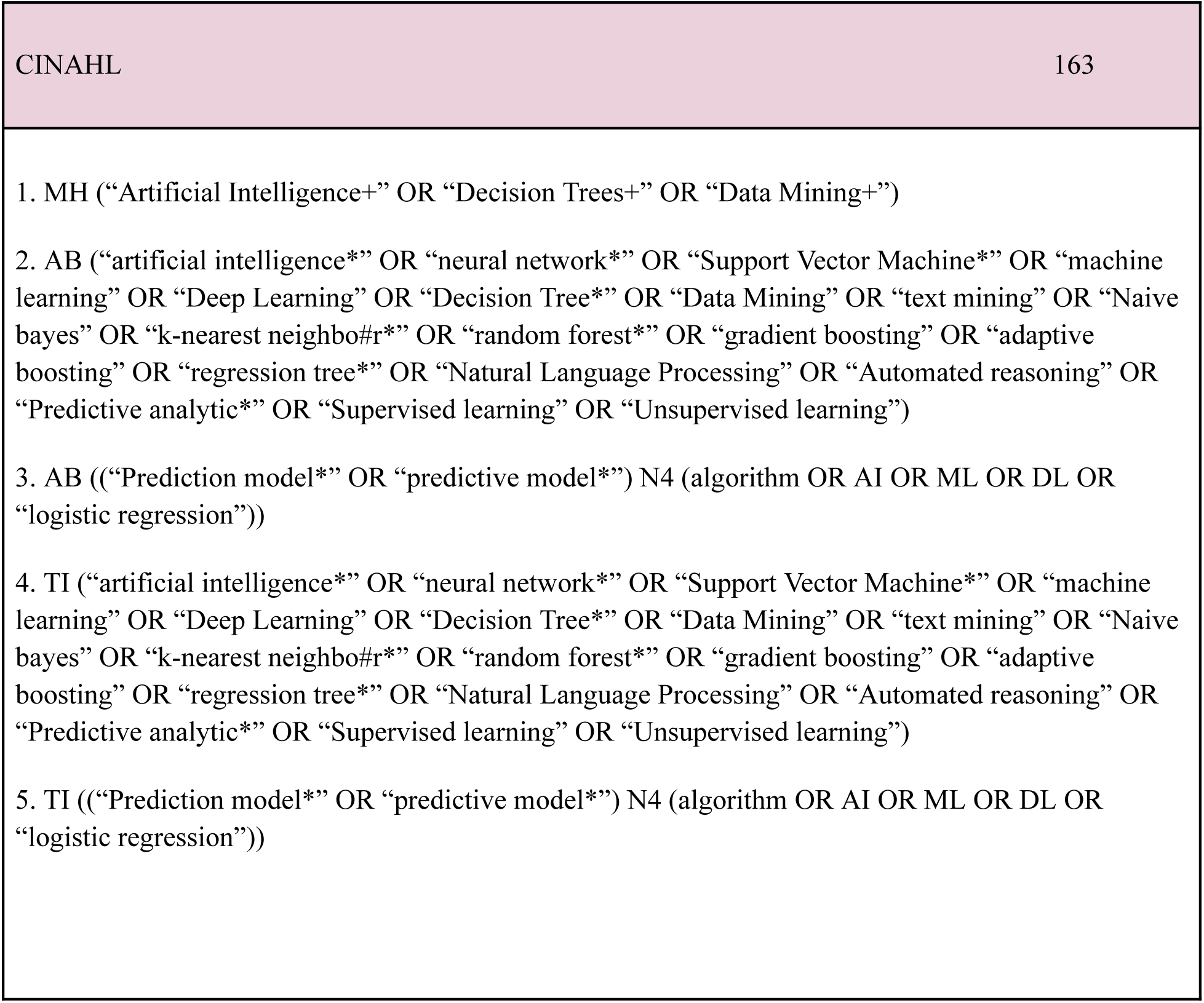
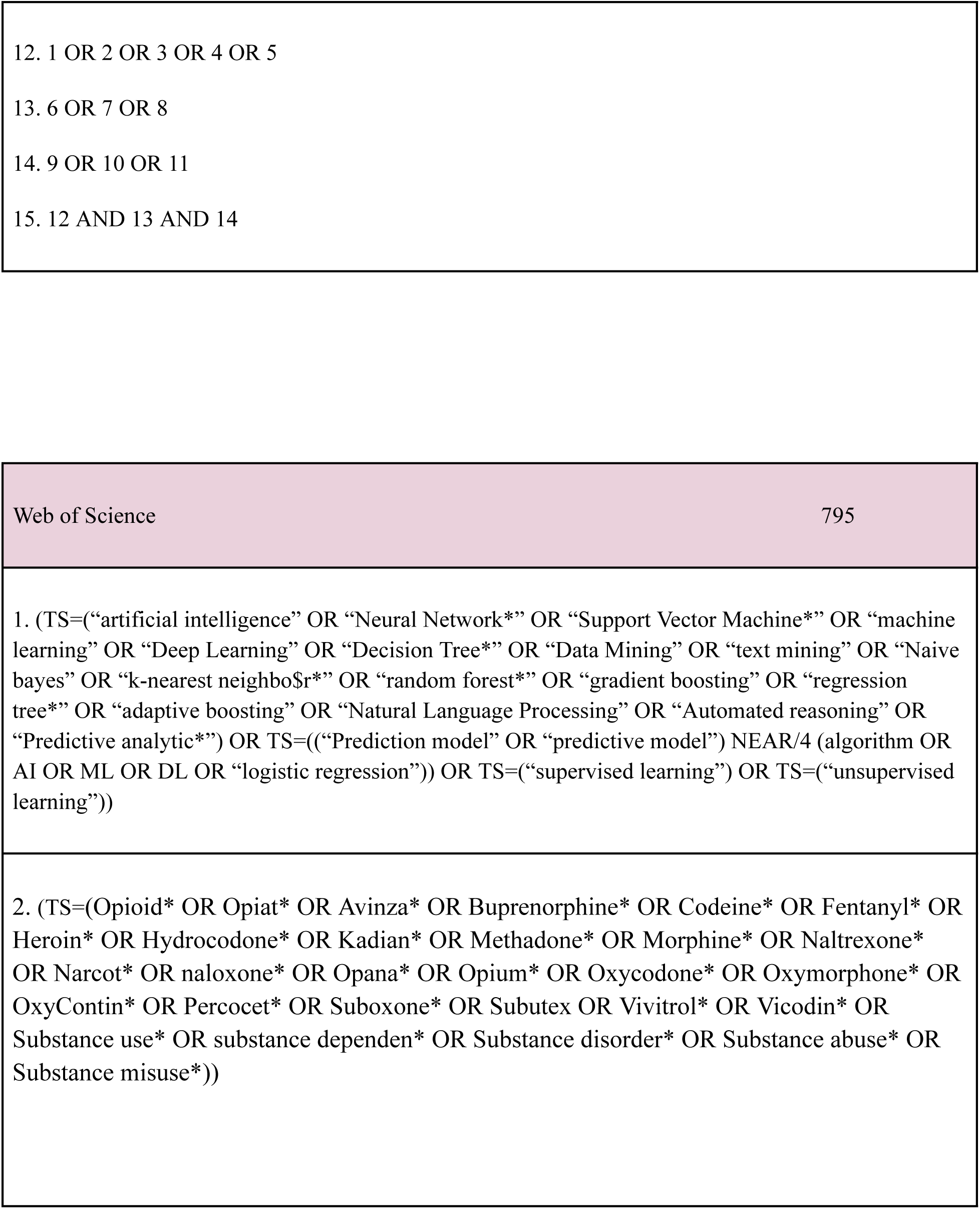
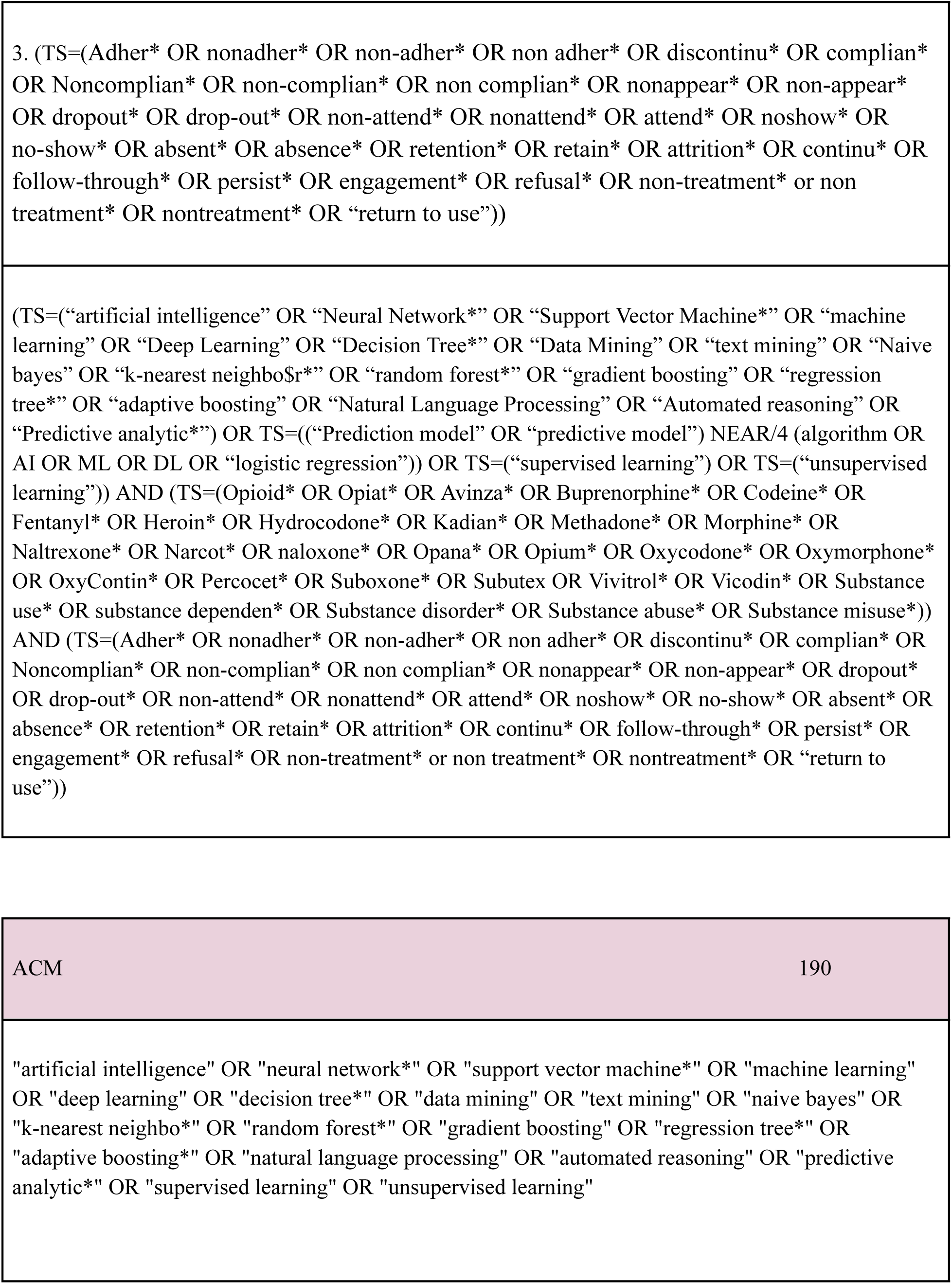
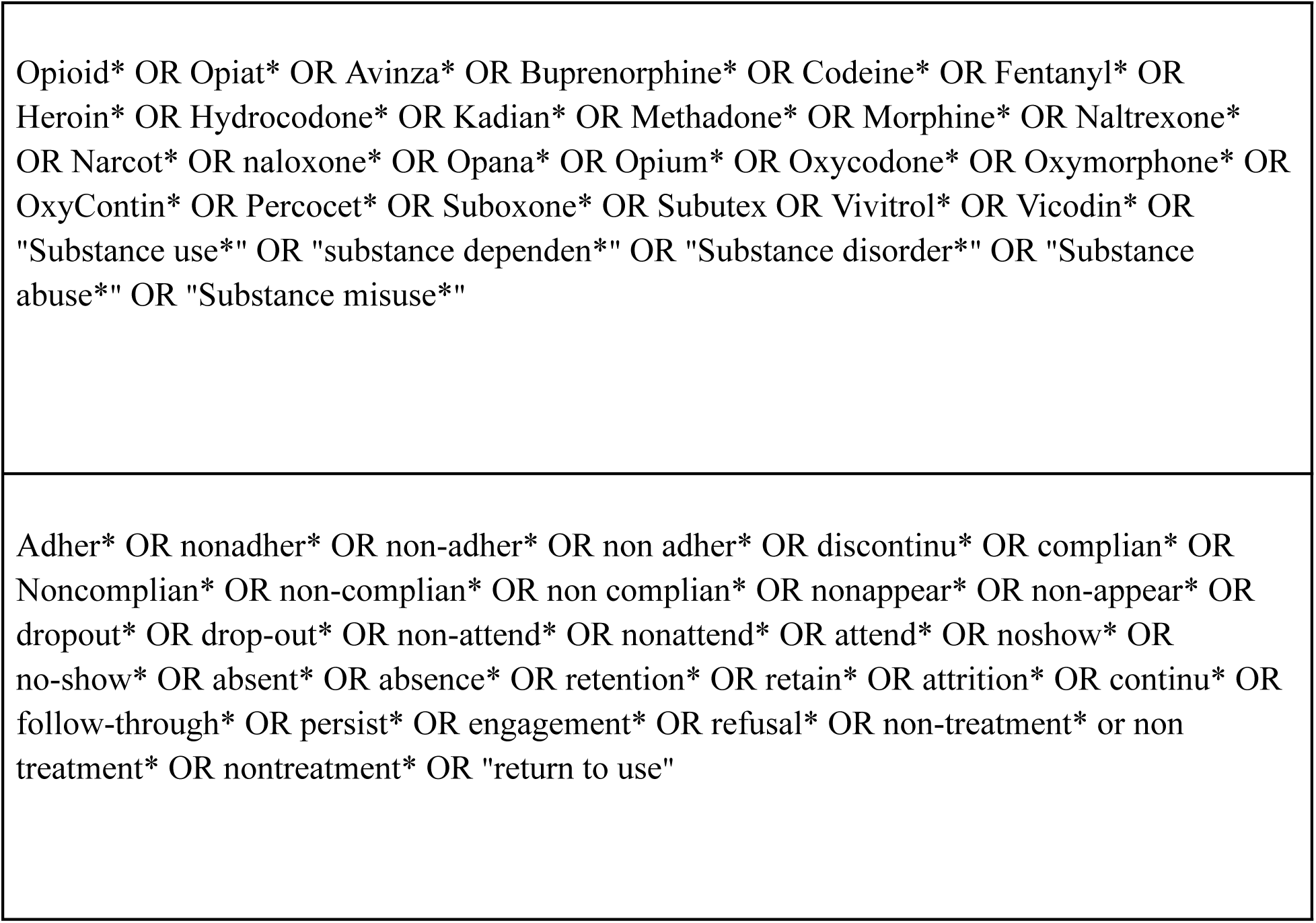
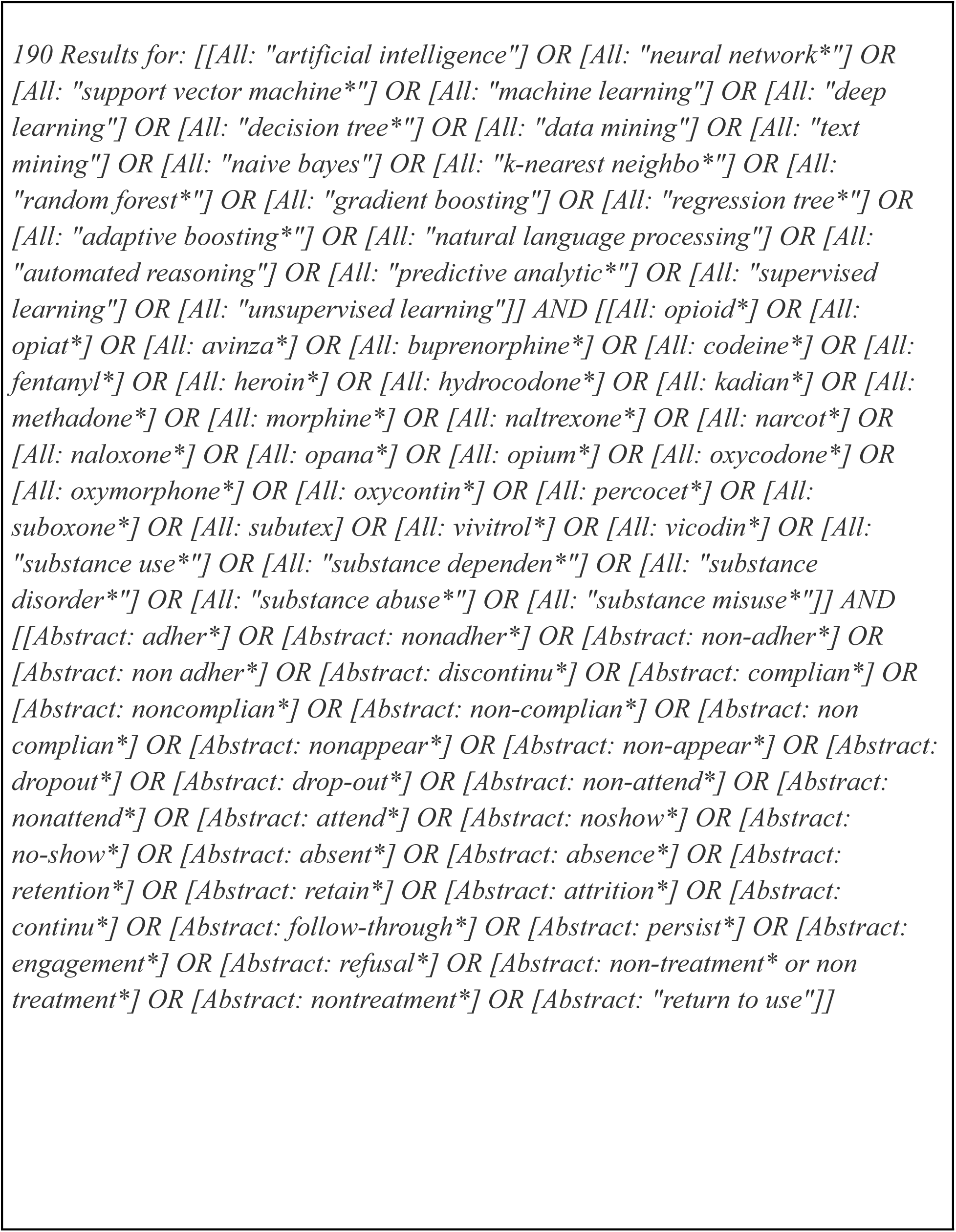
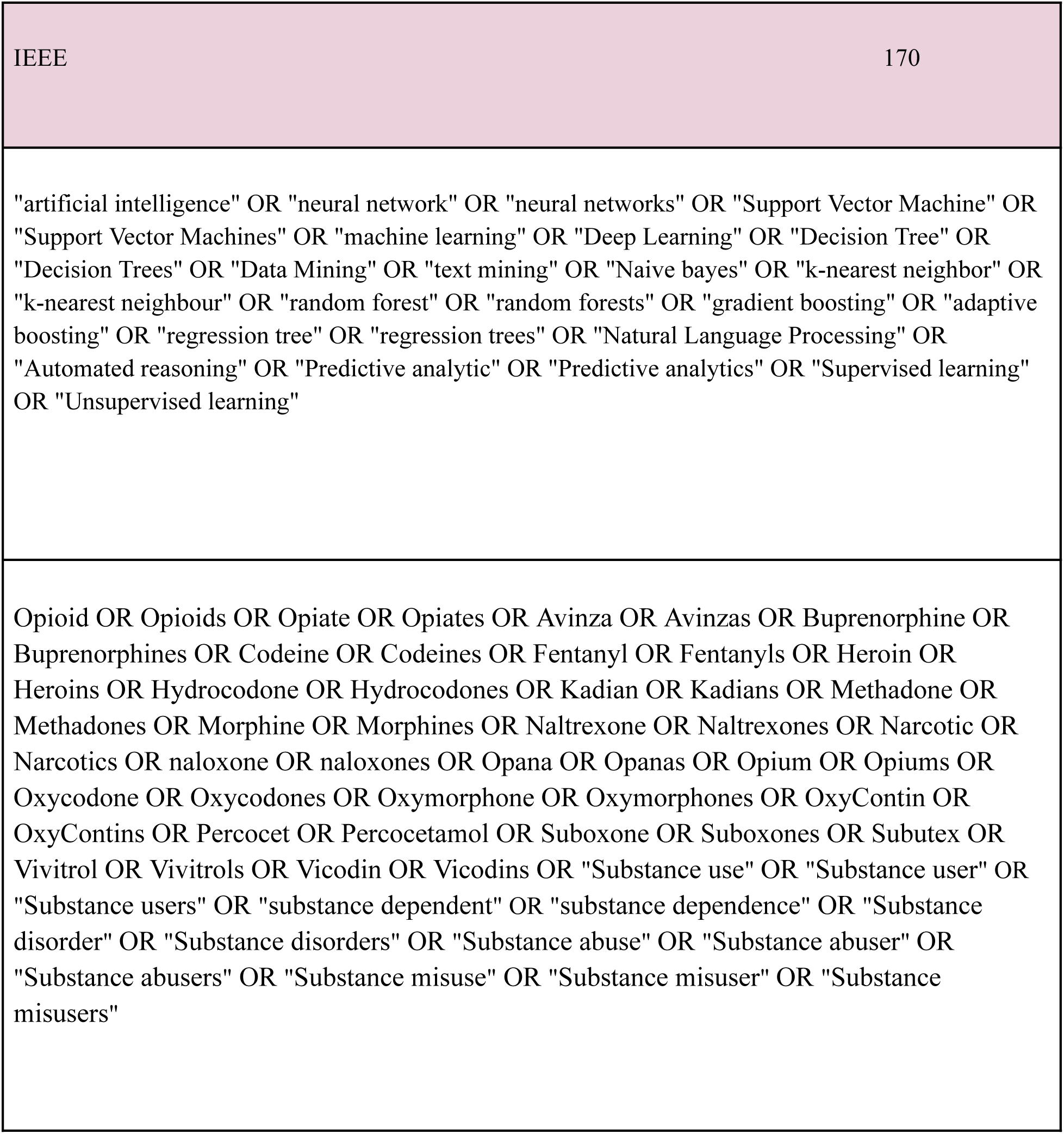
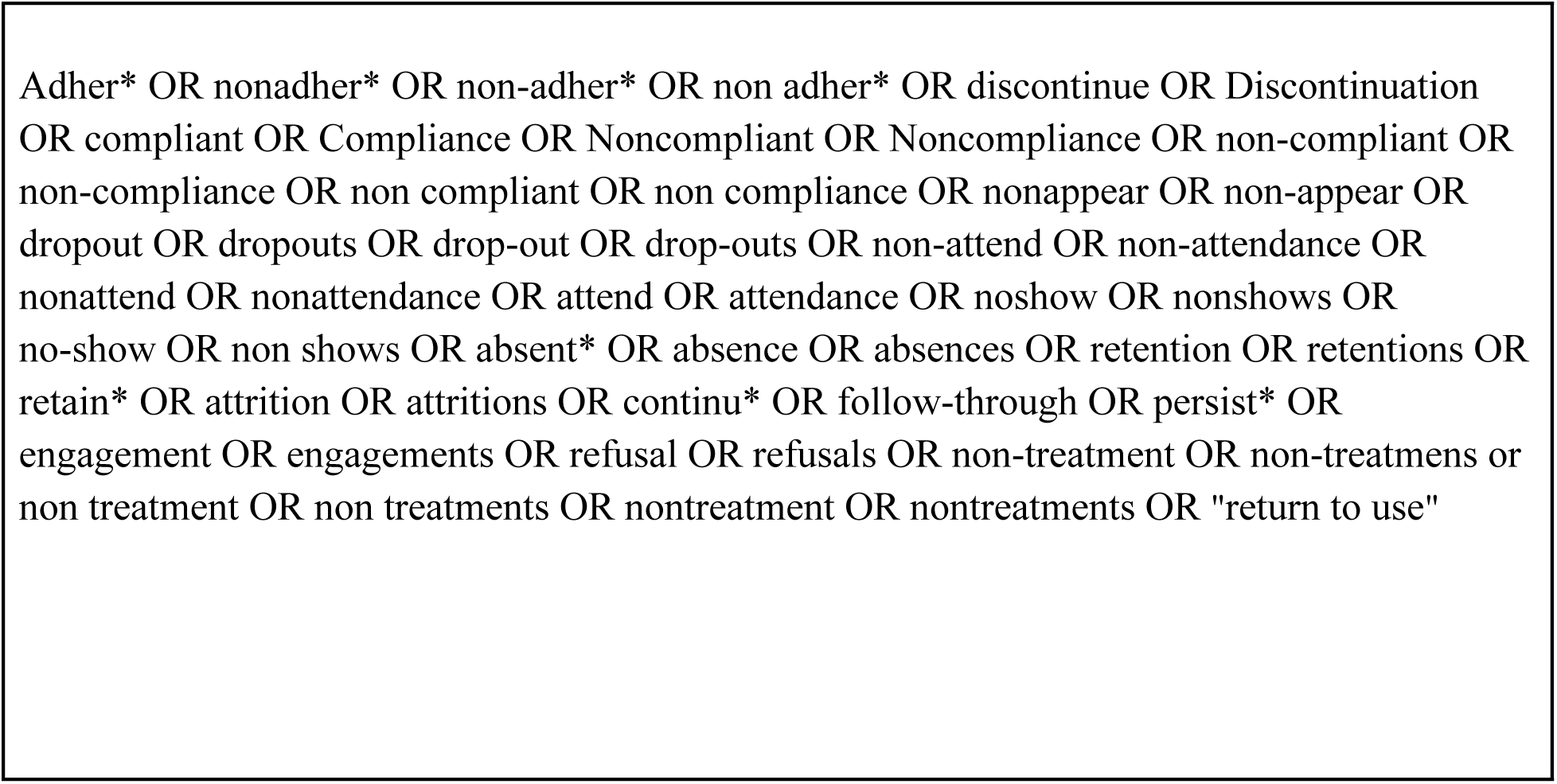
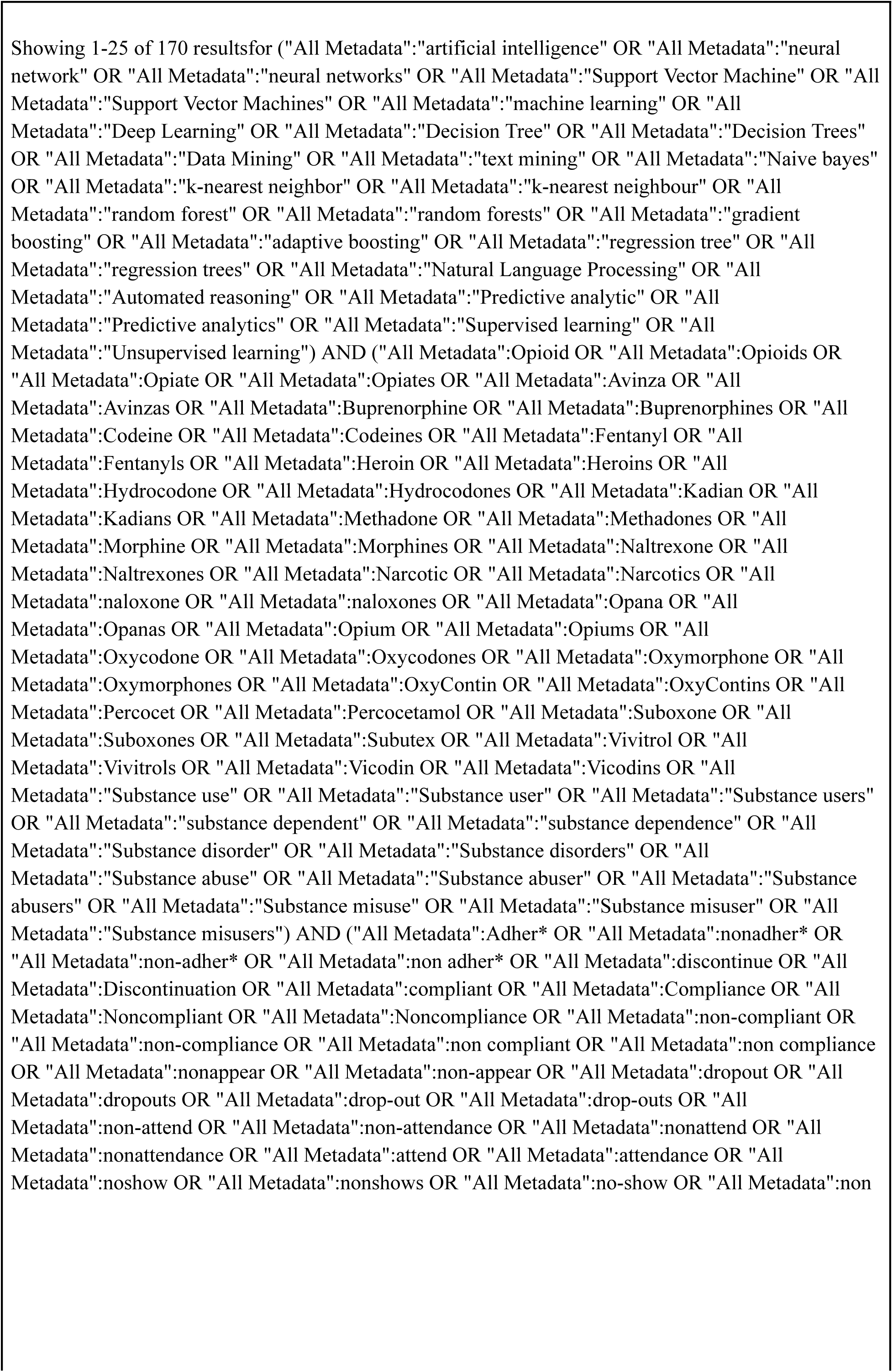
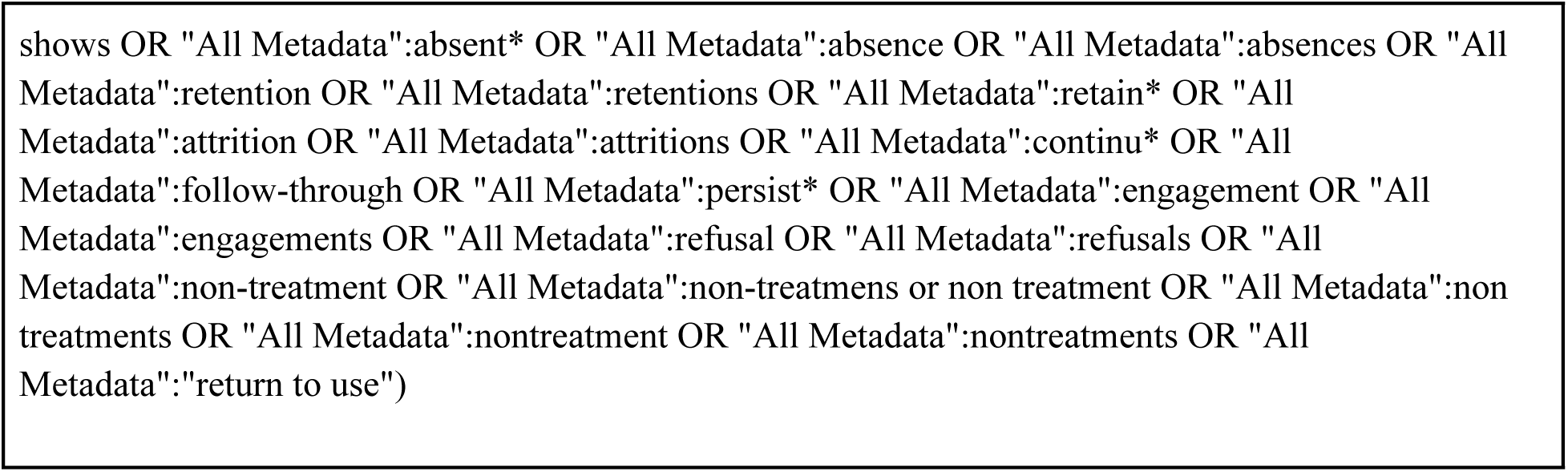
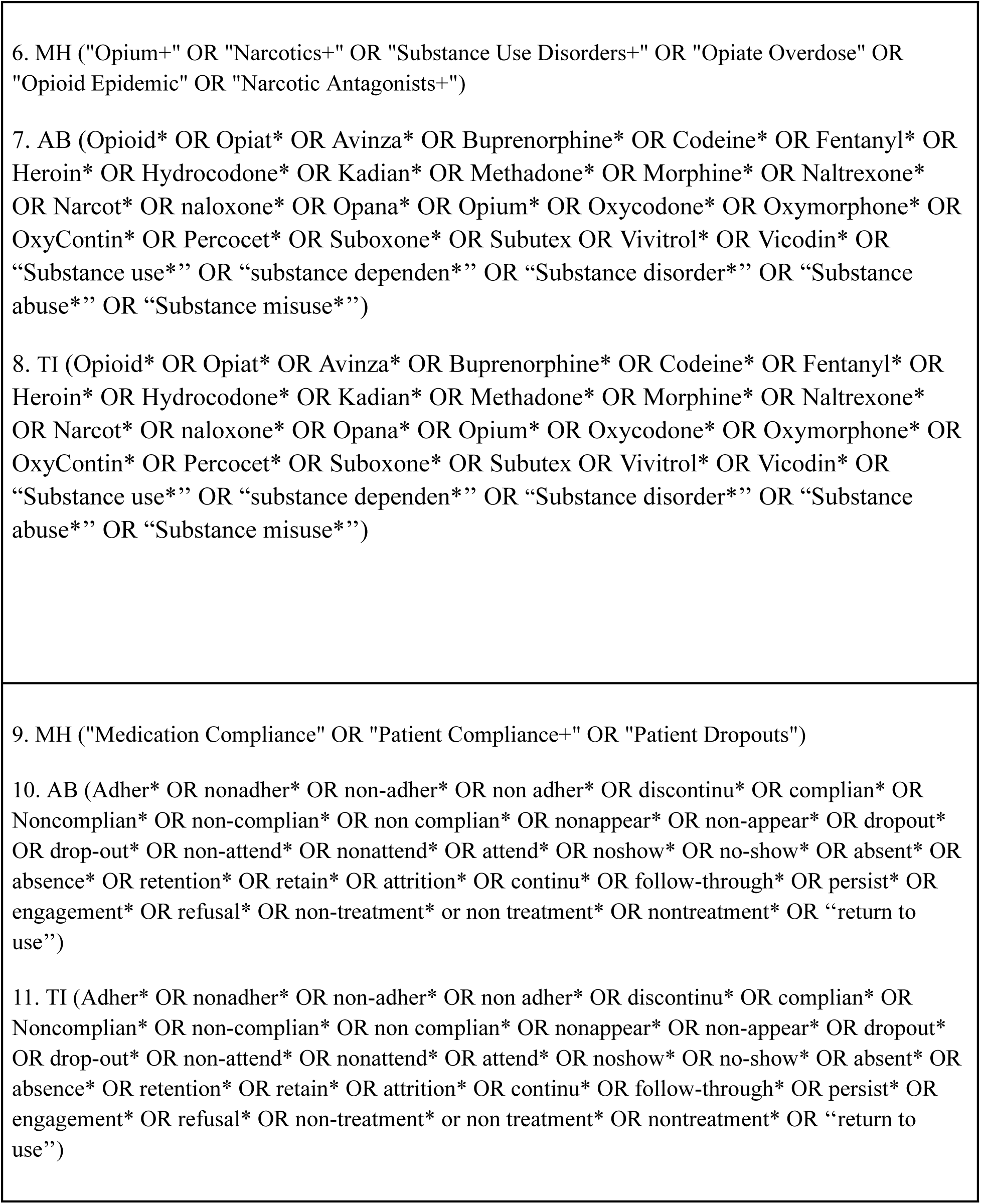
Complete Search Strategy.

## Preferred Reporting Items for Systematic reviews and Meta-Analyses extension for Scoping Reviews (PRISMA-ScR) Checklist

| SECTION | ITEM | PRISMA-ScR CHECKLIST ITEM | REPORTED ON PAGE # |
| --- | --- | --- | --- |
| **TITLE** |  |  |  |
| Title | 1 | Identify the report as a scoping review. | 1 |
| **ABSTRACT** |  |  |  |
| Structured summary | 2 | Provide a structured summary that includes (as applicable): background, objectives, eligibility criteria, sources of evidence, charting methods, results, and conclusions that relate to the review questions and objectives. | 2 |
| **INTRODUCTION** |  |  |  |
| Rationale | 3 | Describe the rationale for the review in the context of what is already known. Explain why the review questions/objectives lend themselves to a scoping review approach. | 4 |
| Objectives | 4 | Provide an explicit statement of the questions and objectives being addressed with reference to their key elements (e.g., population or participants, concepts, and context) or other relevant key elements used to conceptualize the review questions and/or objectives. | 4 |
| **METHODS** |  |  |  |
| Protocol and registration | 5 | Indicate whether a review protocol exists; state if and where it can be accessed (e.g., a Web address); and if available, provide registration information, including the registration number. | 4<br>(https://osf.io/uzbhc/) |
| Eligibility criteria | 6 | Specify characteristics of the sources of evidence used as eligibility criteria (e.g., years considered, language, and publication status), and provide a rationale. | 5 |
| Information sources* | 7 | Describe all information sources in the search (e.g., databases with dates of coverage and contact with authors to identify additional sources), as well as the date the most recent search was executed. | 4 |
| Search | 8 | Present the full electronic search strategy for at least 1 database, including any limits used, such that it could be repeated. | Suppl 1 |
| Selection of sources of evidence† | 9 | State the process for selecting sources of evidence (i.e., screening and eligibility) included in the scoping review. | 4 |
| Data charting process‡ | 10 | Describe the methods of charting data from the included sources of evidence (e.g., calibrated forms or forms that have been tested by the team before their use, and whether data charting was done independently or in duplicate) and any processes for obtaining and confirming data from investigators. | 5 |
| Data items | 11 | List and define all variables for which data were sought and any assumptions and simplifications made. | 5 |
| Critical appraisal of individual sources of evidence§ | 12 | If done, provide a rationale for conducting a critical appraisal of included sources of evidence; describe the methods used and how this information was used in any data synthesis (if appropriate). | N/A |
| Synthesis of results | 13 | Describe the methods of handling and summarizing the data that were charted. | 5 |
| **RESULTS** |  |  |  |
| Selection of sources of evidence | 14 | Give numbers of sources of evidence screened, assessed for eligibility, and included in the review, with reasons for exclusions at each stage, ideally using a flow diagram. | 5 |
| Characteristics of sources of evidence | 15 | For each source of evidence, present characteristics for which data were charted and provide the citations. | Table 1/2 |
| Critical appraisal within sources of evidence | 16 | If done, present data on critical appraisal of included sources of evidence (see item 12). | N/A |
| Results of individual sources of evidence | 17 | For each included source of evidence, present the relevant data that were charted that relate to the review questions and objectives. | Table 1/2 |
| Synthesis of results | 18 | Summarize and/or present the charting results as they relate to the review questions and objectives. | 6-7 |
| **DISCUSSION** |  |  |  |
| Summary of evidence | 19 | Summarize the main results (including an overview of concepts, themes, and types of evidence available), link to the review questions and objectives, and consider the relevance to key groups. | 7-8 |
| Limitations | 20 | Discuss the limitations of the scoping review process. | 9 |
| Conclusions | 21 | Provide a general interpretation of the results with respect to the review questions and objectives, as well as potential implications and/or next steps. | 9 |
| **FUNDING** |  |  |  |
| Funding | 22 | Describe sources of funding for the included sources of evidence, as well as sources of funding for the scoping review. Describe the role of the funders of the scoping review. | 10 |

JB I = Joanna Briggs Institute; PRISMA-ScR = Preferred Reporting Items for Systematic reviews and Meta-Analyses extension for Scoping Reviews.
\* Where *sources of evidence* (see second footnote) are compiled from, such as bibliographic databases, social media platforms, and Web sites.
† A more inclusive/heterogeneous term used to account for the different types of evidence or data sources (e.g., quantitative and/or qualitative research, expert opinion, and policy documents) that may be eligible in a scoping review as opposed to only studies. This is not to be confused with *information sources* (see first footnote).
‡ The frameworks by Arksey and O'Malley (6) and Levac and colleagues (7) and the JB I guidance (4, 5) refer to the process of data extraction in a scoping review as data charting.
§ The process of systematically examining research evidence to assess its validity, results, and relevance before using it to inform a decision. This term is used for items 12 and 19 instead of "risk of bias" (which is more applicable to systematic reviews of interventions) to include and acknowledge the various sources of evidence that may be used in a scoping review (e.g., quantitative and/or qualitative research, expert opinion, and policy document).
From: Tricco AC, Lillie E, Zarin W, O'Brien KK, Colquhoun H, Levac D, et al. PRISMA Extension for Scoping Reviews (PRISMA-ScR): Checklist and Explanation. *Ann Intern Med.* ;169:467–473. doi: 10.7326/M18-0850

## Notes

### Competing Interest Statement

The authors have declared no competing interest.

### Clinical Protocols

https://doi.org/10.17605/OSF.IO/2YT7A

### Funding Statement

This study did not receive any funding.

